# Age-dependent topic modelling of comorbidities in UK Biobank identifies disease subtypes with differential genetic risk

**DOI:** 10.1101/2022.10.23.22281420

**Authors:** Xilin Jiang, Martin Jinye Zhang, Yidong Zhang, Arun Durvasula, Michael Inouye, Chris Holmes, Alkes L. Price, Gil McVean

## Abstract

The analysis of longitudinal data from electronic health records (EHR) has potential to improve clinical diagnoses and enable personalised medicine, motivating efforts to identify disease subtypes from age-dependent patient comorbidity information. Here, we introduce an age-dependent topic modelling (ATM) method that provides a low-rank representation of longitudinal records of hundreds of distinct diseases in large EHR data sets. The model learns, and assigns to each individual, topic weights for several disease topics, each of which reflects a set of diseases that tend to co-occur within individuals as a function of age. Simulations show that ATM attains high accuracy in distinguishing distinct age-dependent comorbidity profiles. We applied ATM to 282,957 UK Biobank samples, analysing 1,726,144 disease diagnoses spanning all 348 diseases with ≥1,000 independent occurrences in the Hospital Episode Statistics (HES) data, identifying 10 disease topics under the optimal model fit. Analysis of an independent cohort, All of Us, with 211,908 samples and 3,098,771 disease diagnoses spanning 233 of the 348 UK Biobank diseases produced highly concordant findings. In UK Biobank we identified 52 diseases with heterogeneous comorbidity profiles (≥500 occurrences assigned to each of ≥2 topics), including breast cancer, type 2 diabetes (T2D), hypertension, and hypercholesterolemia. For most of these diseases, topic assignments were highly age-dependent, suggesting differences in disease aetiology for early-onset vs. late-onset disease. We defined subtypes of the 52 heterogeneous diseases based on the topic assignments, and compared genetic risk across subtypes using polygenic risk scores (PRS). We identified 18 disease subtypes whose PRS differed significantly from other subtypes of the same disease, including a subtype of T2D characterised by cardiovascular comorbidities and a subtype of asthma characterised by dermatological comorbidities. We further identified specific variants underlying these differences such as a T2D-associated SNP in the *HMGA2* locus that has a higher odds ratio in the top quartile of cardiovascular topic weight (1.18±0.02) compared to the bottom quartile (1.00±0.02) (P=3 × 10^-7^ for difference, FDR = 0.0002 < 0.1). In conclusion, ATM identifies disease subtypes with differential genome-wide and locus-specific genetic risk profiles.

## Introduction

Longitudinal electronic health record (EHR) data, encompassing diagnoses across hundreds of distinct diseases, offers immense potential to improve clinical diagnoses and enable personalised medicine^1^. Despite intense interest in both the genetic relationships between distinct diseases^2–11^ and the genetic relationships between biological subtypes of disease^12–15^, there has been limited progress on classifying disease phenotypes into groups of diseases with frequent co-occurrences (comorbidities) and leveraging comorbidities to identify disease subtypes. Low-rank modelling has appealing theoretical properties^16, 17^ and has produced promising applications^18–24^ to infer meaningful representations of high-dimensional data. In particular, low-rank representation is an appealing way to summarise data across hundreds of distinct diseases^25–27^, providing the potential to identify patient-level comorbidity patterns and distinguish disease subtypes. The biological differentiation of disease subtypes inferred from EHR data could be validated by comparing genetic profiles across subtypes, which is possible with emerging data sets that link genetic data with EHR data^28–31^.

Previous studies have used low-rank representation to identify shared genetic components^25–27^ across multiple distinct diseases, identifying relationships between diseases and generating valuable biological insights. However, age at diagnosis information in longitudinal EHR data has the potential to improve such efforts. For example, a recent study used longitudinal disease trajectories to identify disease pairs with statistically significant directionality^32^, suggesting that age information could be leveraged to infer comorbidity profiles that capture temporal information. In addition, patient-level comorbidity information could potentially be leveraged to identify biological subtypes of disease, complementing its application to increase power for identifying genetic associations^12^ and to cluster disease-associated variants into biological pathways^8^; disease subtypes are fundamental to disease aetiology^14, 33–36^.

Here, we propose an age-dependent topic modelling (ATM) method to provide a low-rank representation of longitudinal disease records. ATM learns, and assigns to each individual, topic weights for several disease topics, each of which reflects a set of diseases that tend to co-occur within individuals as a function of age. We applied ATM to 1.7 million disease diagnoses spanning 348 diseases in UK Biobank, inferring 10 disease topics; we validated ATM in All of Us. We identified 52 diseases with heterogeneous comorbidity profiles that enabled us to define disease subtypes. We used genetic data to validate the disease subtypes, showing that they exhibit differential genome-wide and locus-specific genetic risk profiles.

## Results

### Overview of methods

We propose an age-dependent topic modelling (ATM) model, which provides a low-rank representation of longitudinal records of hundreds of distinct diseases in large EHR data sets (Figure 1, Methods). The model assigns to each individual *topic weights* for several *disease topics*; each disease topic reflects a set of diseases that tend to co-occur as a function of age, quantified by age-dependent *topic loadings* for each disease. The model assumes that for each disease diagnosis, a topic is sampled based on the individual’s topic weights (which sum to 1 across topics, for a given individual), and a disease is sampled based on the individual’s age and the age-dependent topic loadings (which sum to 1 across diseases, for a given topic at a given age). The model generalises the latent dirichlet allocation (LDA) model^37, 38^ by allowing topic loadings for each topic to vary with age (Supplementary Note, Supplementary Figure 1).

**Figure 1:**
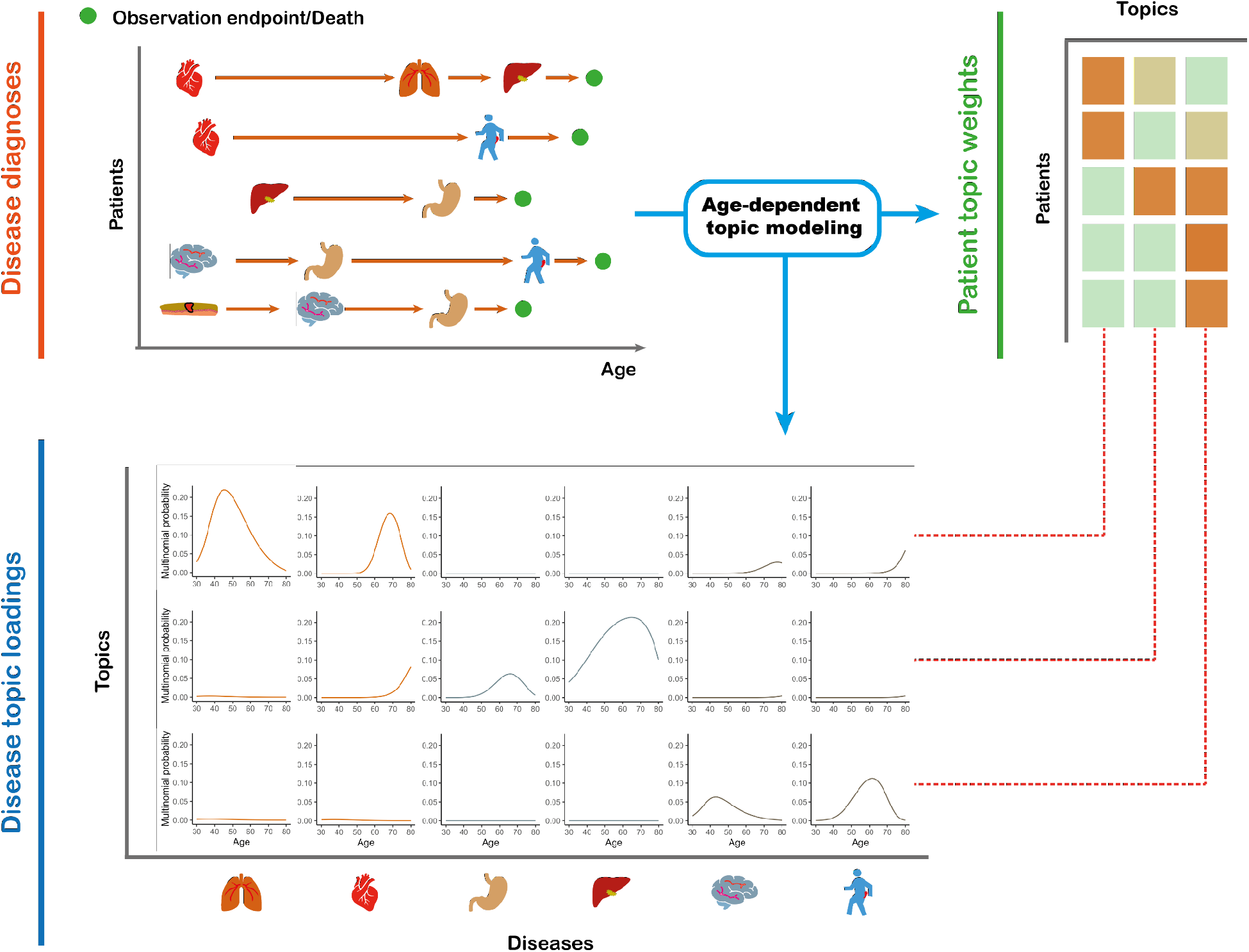
ATM provides an efficient way to represent longitudinal comorbidity data. Top left: input consists of disease diagnoses as a function of age. Top right: ATM assigns a topic weight to each patient. Bottom: ATM infers age-dependent topic loadings.

We developed a method to fit this model that addresses several challenges inherent to large EHR data sets; the method estimates topic weights for each individual, topic loadings for each disease, and posterior diagnosis-specific topic probabilities for each disease diagnosis. First, we derived a scalable deterministic method that uses numerical approximation approaches to fit the parameters of the model, addressing the challenge of computational cost. Second, we used the prediction odds ratio^39^ to compare model structures (e.g. number of topics and parametric form of topic loadings as a function of age), addressing the challenge of appropriate model selection; roughly, the prediction odds ratio quantifies the accuracy of correctly predicting disease diagnoses in held-out individuals using comorbidity information, compared to a predictor based only on prevalence (see Methods and Supplementary Table 1). Third, we employed collapsed variational inference^40^, addressing the challenge of sparsity in the data (e.g. in UK Biobank data, the average patient has diagnoses for 6 of 348 diseases analysed); collapsed variational inference outperformed mean-field variational inference^37^ in empirical data. Further details are provided in the Methods section and Supplementary Note; we have publicly released open-source software implementing the method (see Code Availability).

We applied ATM to longitudinal records of UK Biobank^29^ (282,957 individuals with 1,726,144 disease diagnoses spanning 348 diseases; the targeted individuals are those diagnosed with at least two of the 348 diseases studied) and All of Us^30^ (211,908 individuals with 3,098,771 disease diagnoses spanning 233 of the 348 diseases). Each disease diagnosis has an associated age-at-diagnosis, defined as the earliest age of reported diagnosis of the disease in that individual; we caution that age at diagnosis may differ from age at disease onset (see Discussion). ATM does not use genetic data, but we used genetic data to validate the inferred topics (Methods).

### Simulations

We performed simulations to compare ATM with latent dirichlet allocation (LDA)^37, 38^, a simpler topic modelling approach that does not model age. We simulated 61,000 disease diagnoses spanning 20 diseases in 10,000 individuals, using the ATM generative model; we aimed to choose simulation parameters that resemble real data. In detail, the average number of disease diagnoses per individual (6.1), ratio of #individuals/#diseases (500), topic loadings, and standard deviation in age at diagnosis (8.5 years for each disease) were chosen to match empirical UK Biobank data; we varied the number of topics, number of individuals, and number of diseases in secondary analyses (see below). We assigned each disease diagnosis to one of two subtypes for the target disease based on age and other subtype differences, considering high, medium, or low age-dependent effects by specifying an average difference of 20, 10, or 5 years respectively in age at diagnosis for the two subtypes. For each level of age-dependent effects, we varied the proportion of diagnoses belonging to the first subtype (i.e. the subtype that has an earlier average age-at-diagnosis) from 10-50%. Further details of the simulation framework are provided in the Methods section. Our primary metric for evaluating the LDA and ATM methods is the area under the precision-recall curve (AUPRC)^41^ metric, where precision is defined as the proportion of disease diagnoses that a given method assigned to the first subtype that were assigned correctly and recall is defined as the proportion of disease diagnoses truly belonging to the first subtype that were assigned correctly. We discretized the subtype assigned to each disease diagnosis by a given method by assigning the subtype with higher inferred probability. We note that AUPRC is larger when classifying the minority subtype; results using the second subtype as the classification target are also provided. We used AUPRC (instead of prediction odds ratio) in our simulations because the underlying truth is known. Further details and justifications of metrics used in this study are provided in the Methods Section and Supplementary Table 1.

In simulations with high age-dependent effects, ATM attained much higher AUPRC than LDA across all values of subtype sample size proportion (AUPRC difference: 24%-42%), with both methods performing better at more balanced ratios (Figure 2, Supplementary Table 2). Accordingly, ATM attained both higher precision and higher recall than LDA (Supplementary Figure 2). Results were qualitatively similar when using the second subtype as the classification target (Supplementary Figure 3). In simulations with medium or low age-dependent effects, ATM continued to outperform LDA but with smaller differences between the methods. In simulations without age-dependent effects, ATM slightly underperformed LDA (Supplementary Figure 4A).

**Figure 2:**
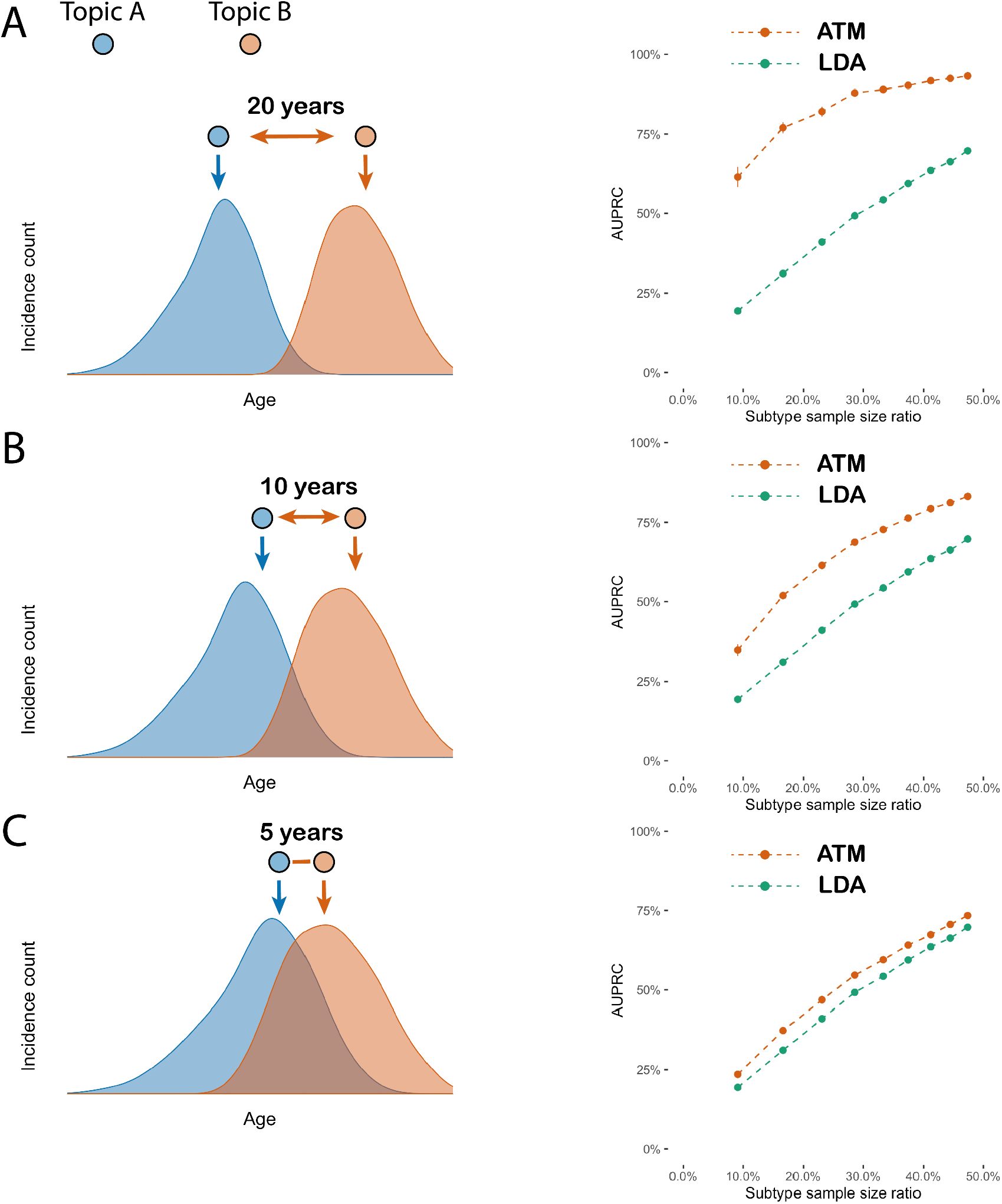
ATM outperforms LDA in simulations with age-dependent effects. In simulations at different levels of age-dependent effects (left panels), we report the area under the precision and recall curve (AUPRC) for ATM vs. LDA as a function of subtype sample size proportion (the proportion of diagnoses belonging to the smaller subtype) (right panels). Each dot represents the mean of 100 simulations of 10,000 individuals. Error bars denote 95% confidence intervals. (A) 20-year difference in age at diagnosis for the two subtypes. (B) 10-year difference in age at diagnosis for the two subtypes. (C) 5-year difference in age at diagnosis for the two subtypes. Numerical results are reported in Supplementary Table 2.

**Figure 3.**
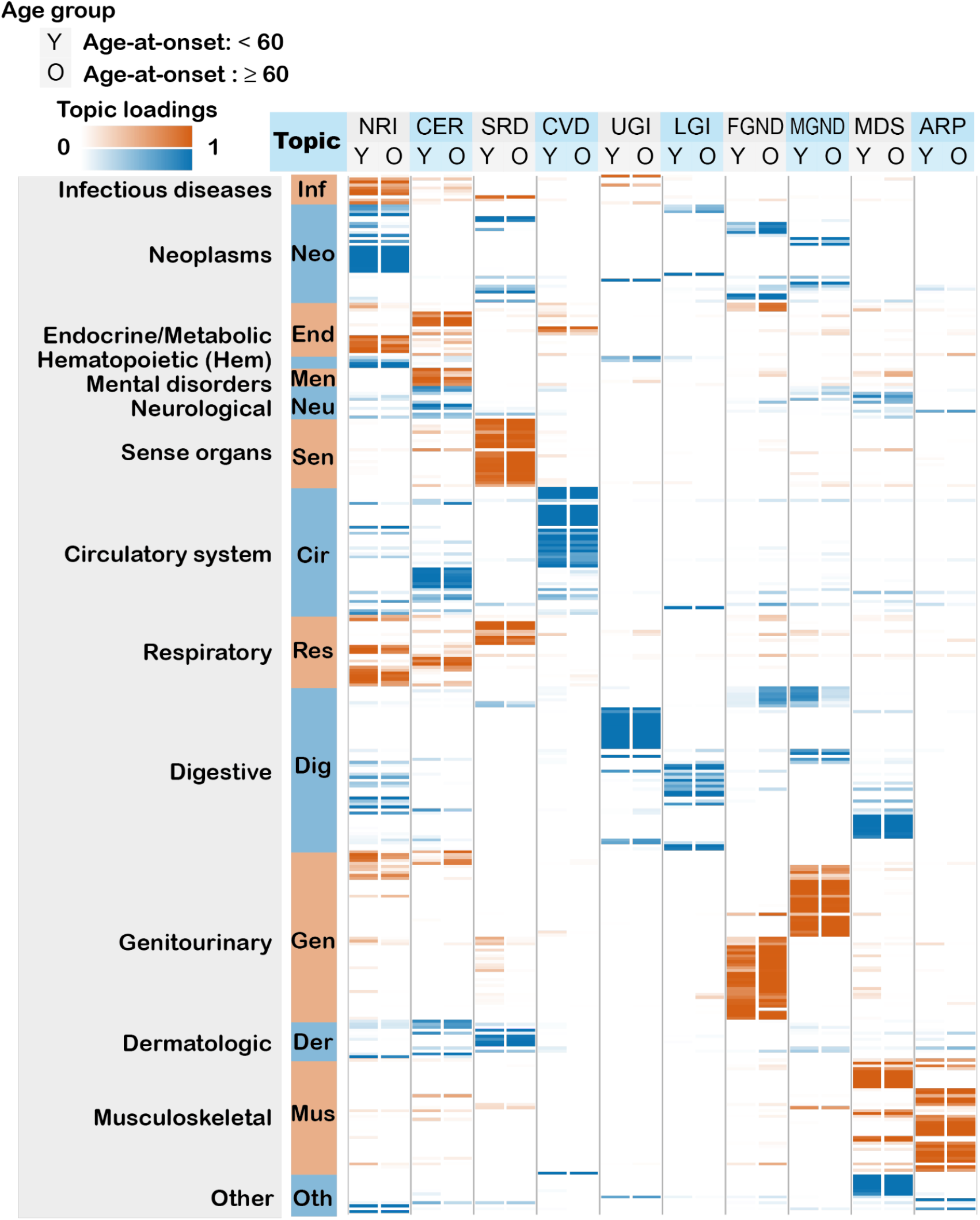
Age-dependent topic loadings of 10 inferred disease topics across 348 diseases in the UK Biobank. We report topic loadings averaged across younger ages (age at diagnosis < 60) and older ages (age at diagnosis > 60). Row labels denote disease categories ordered by Phecode systems, with alternating blue and red color for visualisation purposes; “Other” is a merge of five Phecode systems: “congenital anomalies”, “symptoms”, “injuries & poisoning”, “other tests”, and “death” (which is treated as an additional disease, see Methods). Topics are ordered by the corresponding Phecode system. Further details on the 10 topics are provided in Table 1. Further details on the diseases discussed in the text (type 2 diabetes and breast cancer) are provided in Supplementary Figure 6. Numerical results are reported in Supplementary Table 4.

**Figure 4.**
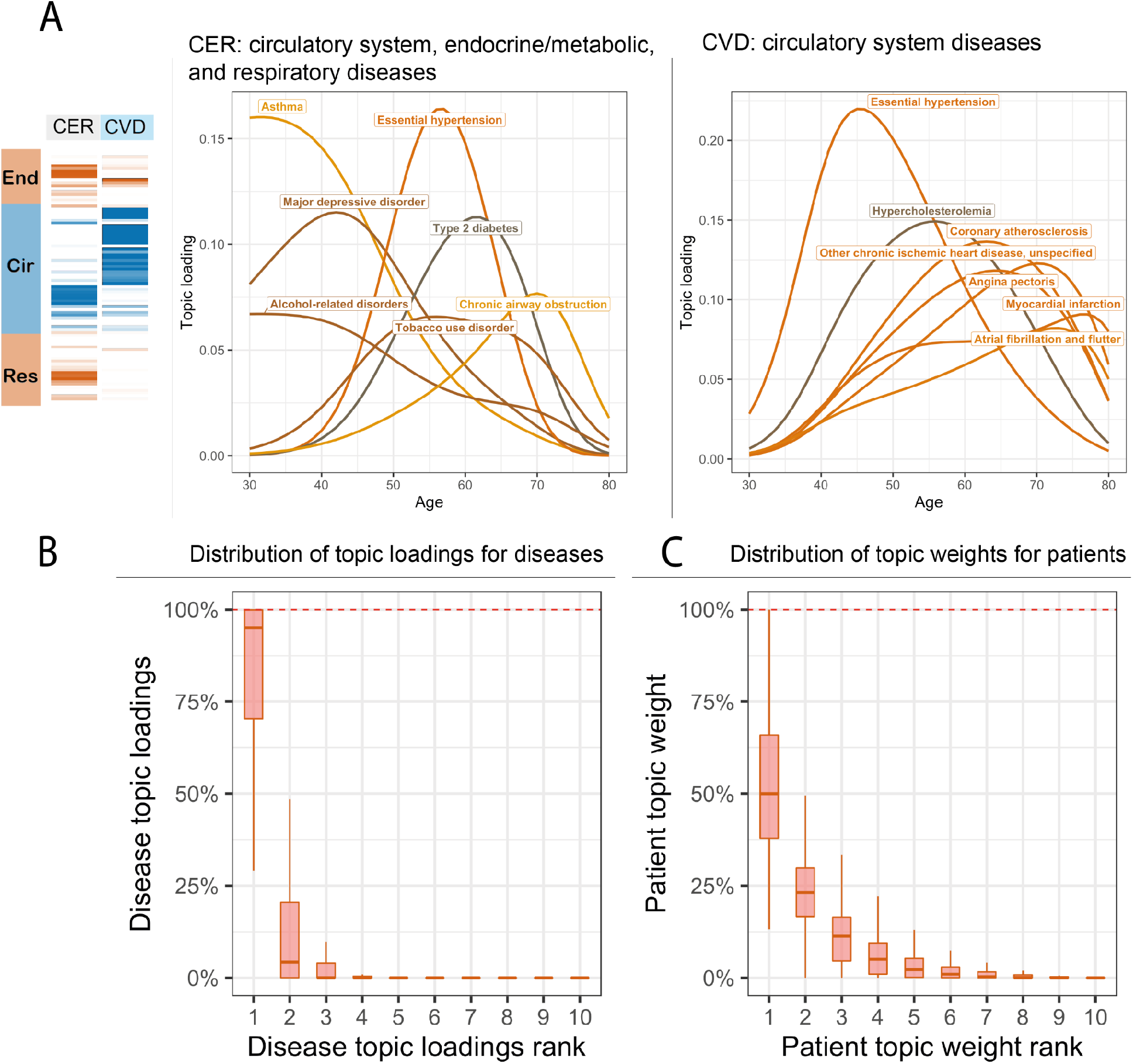
Topic loadings in UK Biobank capture age-dependent comorbidities. (A) Age-dependent topic loadings for two representative topics, CER and CVD; for each topic, we include the top seven diseases with highest topic loadings. Results for all 10 topics are reported in Supplementary Figure 13. (B) Box plot of disease topic loading as a function of rank; disease topic loadings are computed as a weighted average across all values of age at diagnosis. (C) Box plot of patient topic weight as a function of rank. Numerical results are reported in Supplementary Table 5.

**Table 1.**
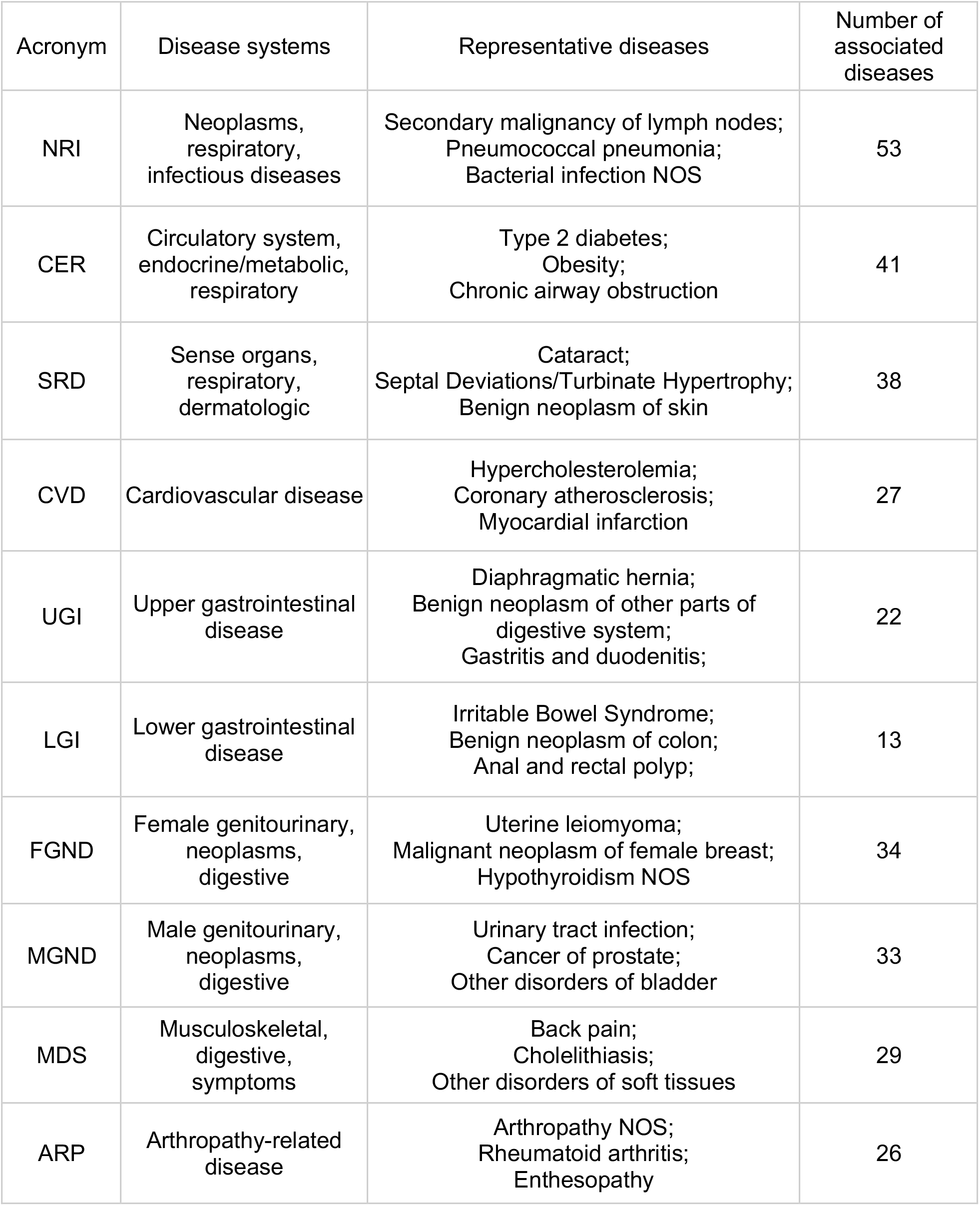
Summary of 10 inferred disease topics in the UK Biobank. For each topic, we list its 3-letter acronym, disease systems, representative diseases, and number of associated diseases (defined as diseases with average diagnosis-specific topic probability >50% for that topic). Topics are ordered by the Phecode system (see Figure 3). 316 of 348 diseases analysed are associated with a topic; the remaining 32 diseases do not have a topic with average diagnosis-specific topic probability >50%.

| Acronym | Disease systems | Representative diseases | Number of associated diseases |
| --- | --- | --- | --- |
| NRI | Neoplasms, respiratory, infectious diseases | Secondary malignancy of lymph nodes;<br>Pneumococcal pneumonia;<br>Bacterial infection NOS | 53 |
| CER | Circulatory system, endocrine/metabolic, respiratory | Type 2 diabetes;<br>Obesity;<br>Chronic airway obstruction | 41 |
| SRD | Sense organs, respiratory, dermatologic | Cataract;<br>Septal Deviations/Turbinate Hypertrophy;<br>Benign neoplasm of skin | 38 |
| CVD | Cardiovascular disease | Hypercholesterolemia;<br>Coronary atherosclerosis;<br>Myocardial infarction | 27 |
| UGI | Upper gastrointestinal disease | Diaphragmatic hernia;<br>Benign neoplasm of other parts of digestive system;<br>Gastritis and duodenitis; | 22 |
| LGI | Lower gastrointestinal disease | Irritable Bowel Syndrome;<br>Benign neoplasm of colon;<br>Anal and rectal polyp; | 13 |
| FGND | Female genitourinary, neoplasms, digestive | Uterine leiomyoma;<br>Malignant neoplasm of female breast;<br>Hypothyroidism NOS | 34 |
| MGND | Male genitourinary, neoplasms, digestive | Urinary tract infection;<br>Cancer of prostate;<br>Other disorders of bladder | 33 |
| MDS | Musculoskeletal, digestive, symptoms | Back pain;<br>Cholelithiasis;<br>Other disorders of soft tissues | 29 |
| ARP | Arthropathy-related disease | Arthropathy NOS;<br>Rheumatoid arthritis;<br>Enthesopathy | 26 |

We performed three secondary analyses. First, we varied the number of individuals, number of diseases, or number of disease diagnoses per individual. ATM continued to outperform LDA in each case, although increasing the number of individuals or the number of disease diagnoses per individual did not always increase AUPRC (Supplementary Figure 4B). Second, we performed simulations in which we increased the number of subtypes from two to five and changed the number of diseases to 50, and compared ATM models trained using different numbers of topics (in 80% training data) by computing the prediction odds ratio; we used the prediction odds ratio (instead of AUPRC) in this analysis both because it is a better metric to evaluate the overall model fit to the data, and because it is unclear how to compare AUPRC across scenarios of varying topic numbers (see Supplementary Table 1). We confirmed that the prediction odds ratio was maximised using five topics, validating the use of the prediction odds ratio for model selection (Supplementary Figure 5A). Third, we computed the accuracy of inferred topic loadings, topic weights, and grouping accuracy (defined as proportion of pairs of diseases truly belonging to the same topic that ATM correctly assigned to the same topic), varying the number of individuals and number of diseases diagnoses per individual. We determined that ATM also performed well under these metrics (Supplementary Figure 5B-E).

**Figure 5.**
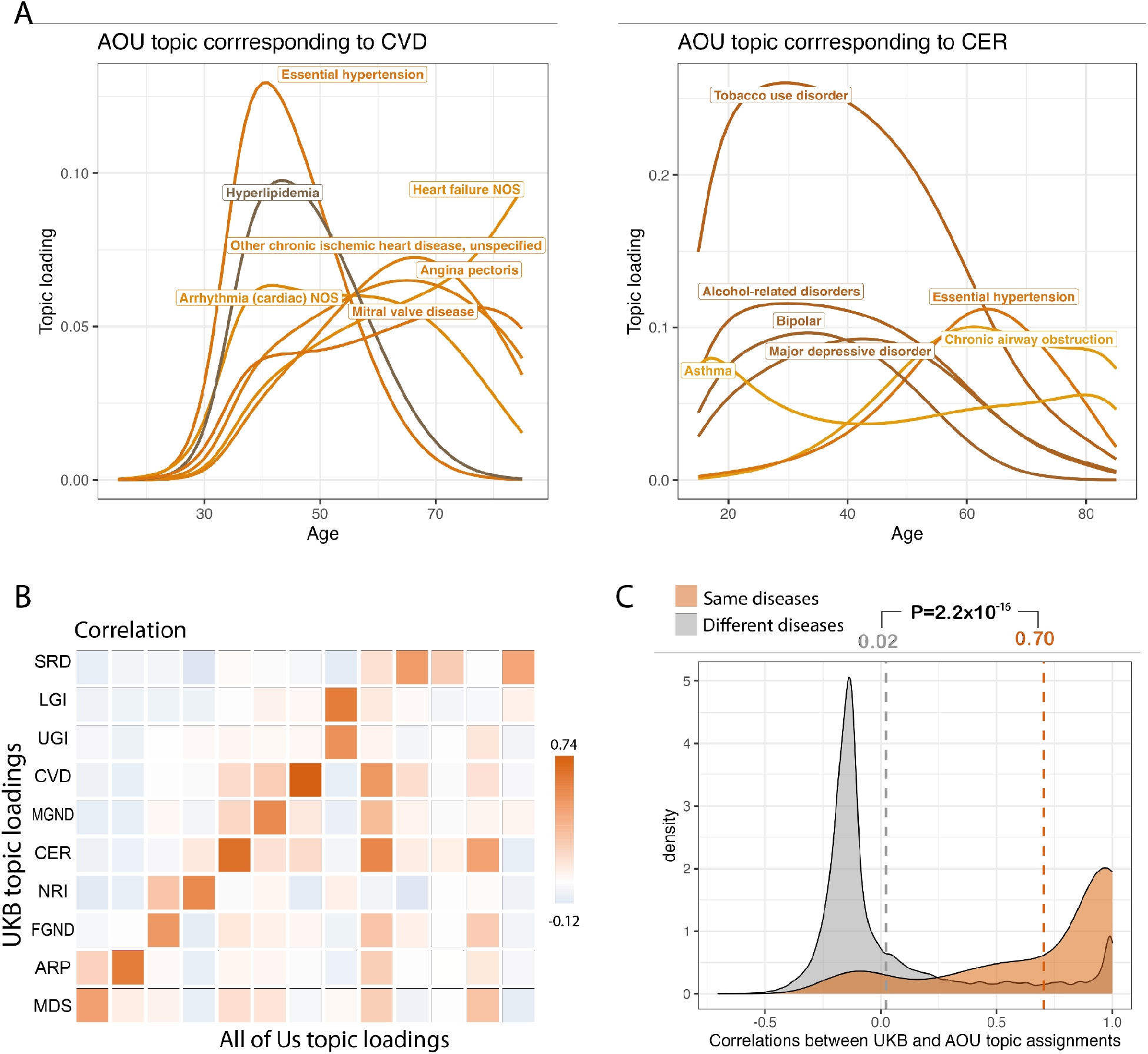
Topic loadings in All of Us capture age-dependent comorbidities that are concordant with UK Biobank. (A) Age-dependent topic loadings for two All of Us topics corresponding to CVD and CER (Figure 4A); for each topic, we include the top seven diseases with highest topic loadings. Correlations of topic loadings between UK Biobank and All of Us topics were 0.74 for CVD and 0.65 for CER. Numerical results for all 13 topics are reported in Supplementary Table 6. (B) Topic loading correlations between UK Biobank (UKB) and All of Us (AOU). The y-axis reflects the 10 topics from the optimal UK Biobank model; the x-axis reflects the 13 topics from the optimal All of Us model. Numerical results are reported in Supplementary Table 7. (C) Correlations between UKB and AOU topic assignments were higher for same diseases (red shading, average = 0.70) than for different diseases (grey shading, average = 0.02)[PA1] . Numerical results are reported in Supplementary Table 9.

We conclude that ATM (which models age) assigns disease diagnoses to subtypes with higher accuracy than LDA (which does not model age) in simulations with age-dependent effects. We caution that our simulations largely represent a best-case scenario for ATM given that the generative model and inference model are very similar (although there are some differences, e.g. topic loadings were generated using a model different from the inference model), thus it is important to analyse empirical data to validate the method.

### Age-dependent disease topic loadings capture comorbidity profiles in the UK Biobank

We applied ATM to longitudinal hospital records of 282,957 individuals from the UK Biobank with an average record span of 28.6 years^29^. We used Phecode^42^ to define 1,726,144 disease diagnoses spanning 348 diseases with at least 1,000 diagnoses each; the average individual had 6.1 disease diagnoses, and the average disease had a standard deviation of 8.5 years in age at diagnosis. The optimal inferred ATM model structure has10 topics and models age-dependent topic loadings for each disease as a spline function with one knot, based on optimizing prediction odds ratio (see below). We assigned names (and corresponding acronyms) to each of the 10 inferred topics based on the Phecode systems^42^ assigned to diseases with high topic loadings (aggregated across ages) for that topic (Table 1, Supplementary Table 3).

Age-dependent topic loadings across all 10 topics and 348 diseases (stratified into Phecode systems), summarised as averages across age<60 and age≥60, are reported in Figure 3, Supplementary Figure 6, and Supplementary Table 4. Some topics such as NRI span diseases across the majority of Phecode systems, while other topics such as ARP are concentrated in a single Phecode system. Conversely, a single Phecode system may be split across multiple topics, e.g. diseases of the digestive system are split across UGI, LGI, and MDS. We note that topic loadings in diseases that span multiple topics are heavily age-dependent. For example, type 2 diabetes patients assigned to the CVD topic are associated with early onset of type 2 diabetes whereas type 2 diabetes patients assigned to the MGND topic are associated with late onset of type 2 diabetes.

**Figure 6.**
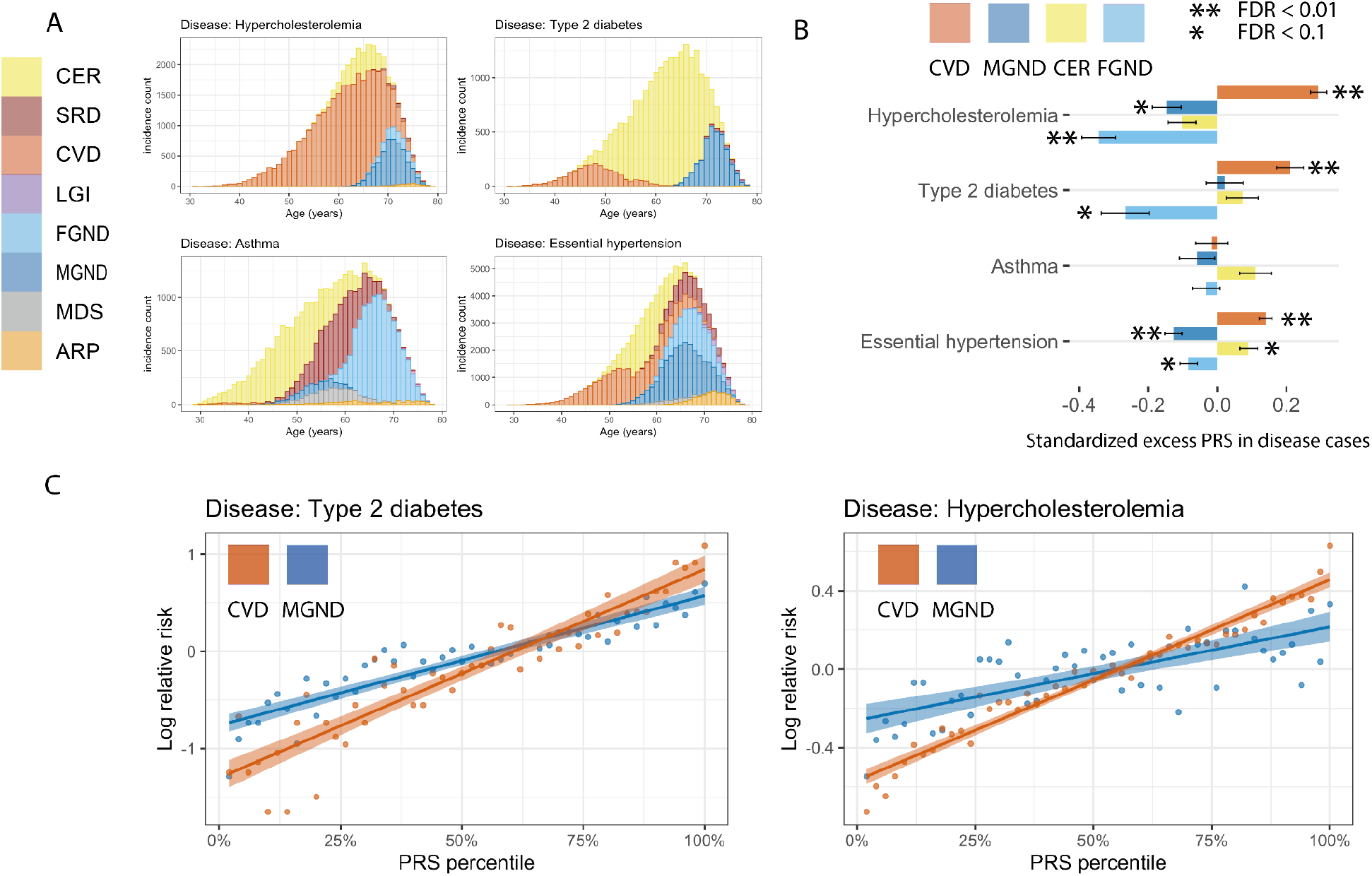
Polygenic risk scores vary across disease subtypes defined by distinct topics. (A) Stacked barplots of age-dependent subtypes (defined by topics) for 4 representative diseases (type 2 diabetes, asthma, hypercholesterolemia, and essential hypertension); for each disease, we include all subtypes with at least one diagnosis. Results for all 52 diseases are reported in Supplementary Figure 21. (B) Standardised excess PRS values in disease cases (s.d. increase in PRS per unit increase in patient topic weight) for 4 representative diseases and 4 corresponding topics. (C) Relative risk for cases of type 2 diabetes and hypercholesterolemia of CVD and MGND subtypes (vs. controls) across PRS percentiles. Each point spans 2 PRS percentiles. Lines denote regression on log scale. Error bars denote 95% confidence intervals. Numerical results are reported in Supplementary Table 10-12.

We performed seven secondary analyses to validate the integrity and reproducibility of inferred comorbidity topics. First, we fit ATM models with different model structures using 80% training data, and computed their prediction odds ratios using 20% testing data. The ATM model structure with 10 topics and age-dependent topic loadings modelled as a spline function performed optimally (Supplementary Figure 7; see Methods). Second, we confirmed that ATM (which models age) attained higher prediction odds ratios than LDA (which does not model age) across different values of the number of topics (Supplementary Figure 8); for the optimal model with 10 topics, ATM attained an average prediction odds ratio of 1.71, compared to a prediction odds ratio of 1.58 for LDA. Third, we reached similar conclusions using evidence lower bounds^39^ (ELBO; see Supplementary Table 1) (Supplementary Figure 9). Fourth, we confirmed that collapsed variational inference ^40^ outperformed mean-field variational inference ^37^ (Supplementary Figure 10). Fifth, we computed a co-occurrence odds ratio evaluating whether diseases grouped into the same topic by ATM in the training data have higher than random probability of co-occurring in the testing data (Supplementary Table 1). The co-occurrence odds ratio is consistently above one and increases with the number of comorbid diseases, for each inferred topic (Supplementary Figure 11). Sixth, we compared the topic loadings by repeating the inference on female-only or male-only populations and observed no major discrepancies, except for genitourinary topics MGND and FGND (topic loading *R*^2^ (female vs. all) = 0.788, topic loading *R*^2^ (male vs. all) = 0.773, Supplementary Figure 12). Lastly, we verified that BMI, sex, Townsend deprivation index, and birth year explained very little of the information in the inferred topics (Supplementary Table 3).

**Figure 7.**
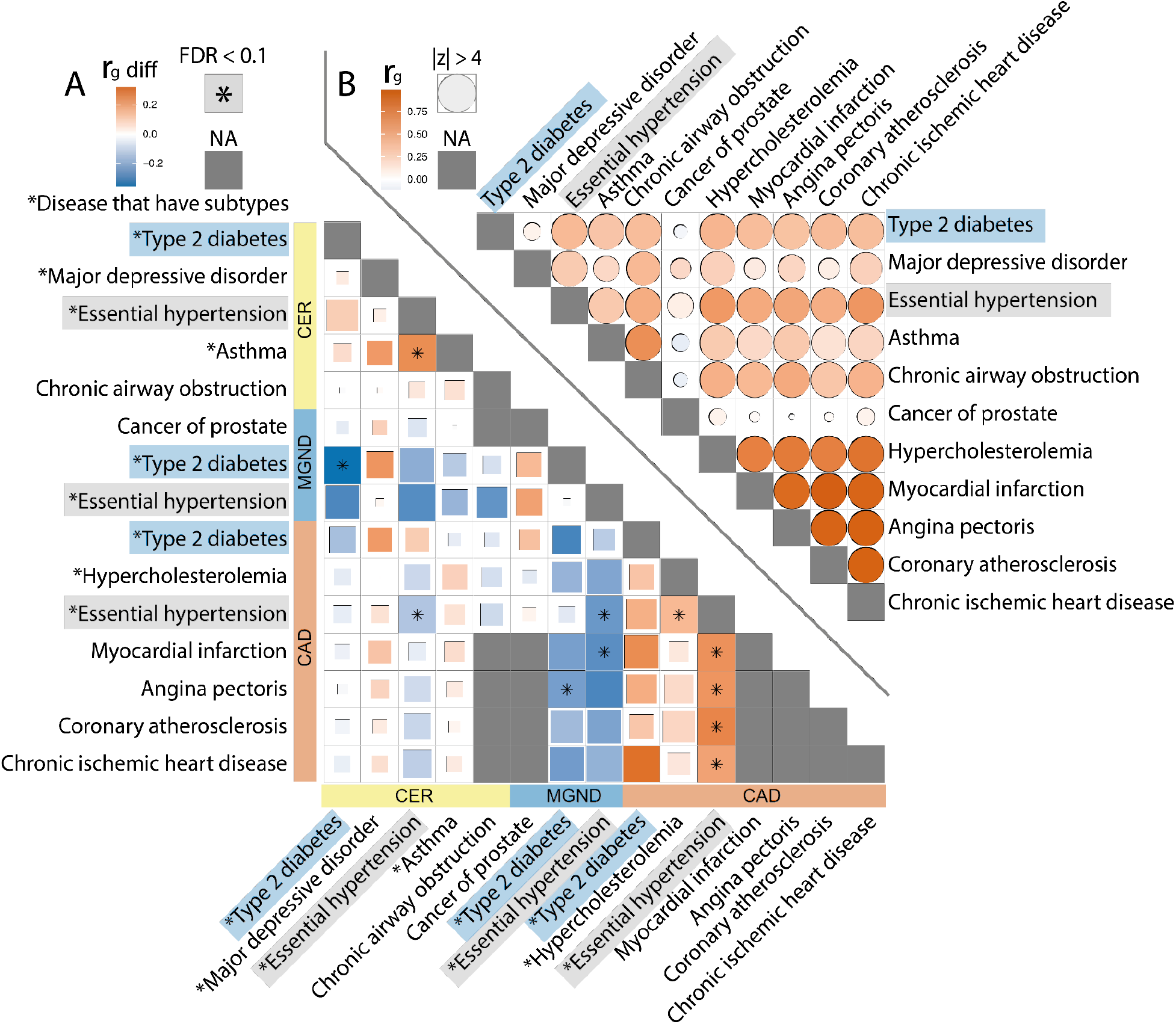
Genetic correlations vary across disease subtypes defined by distinct topics. (a) Excess genetic correlations for pairs of 15 disease subtypes or diseases (9 disease subtypes (denoted with asterisks) + 6 diseases without subtypes), relative to genetic correlations between the underlying diseases. Full square with asterisk denotes FDR < 0.1; less than full squares have area proportional to z-scores for difference. Grey squares denote NA (pair of diseases without subtypes or pair of same disease subtype or disease). (b) Genetic correlations between the underlying diseases. Full circle denotes |z-score| > 4 for nonzero genetic correlation; less than full circles have area proportional to |z-score|. Numerical results are reported in Supplementary Table 13.

**Figure 8.**
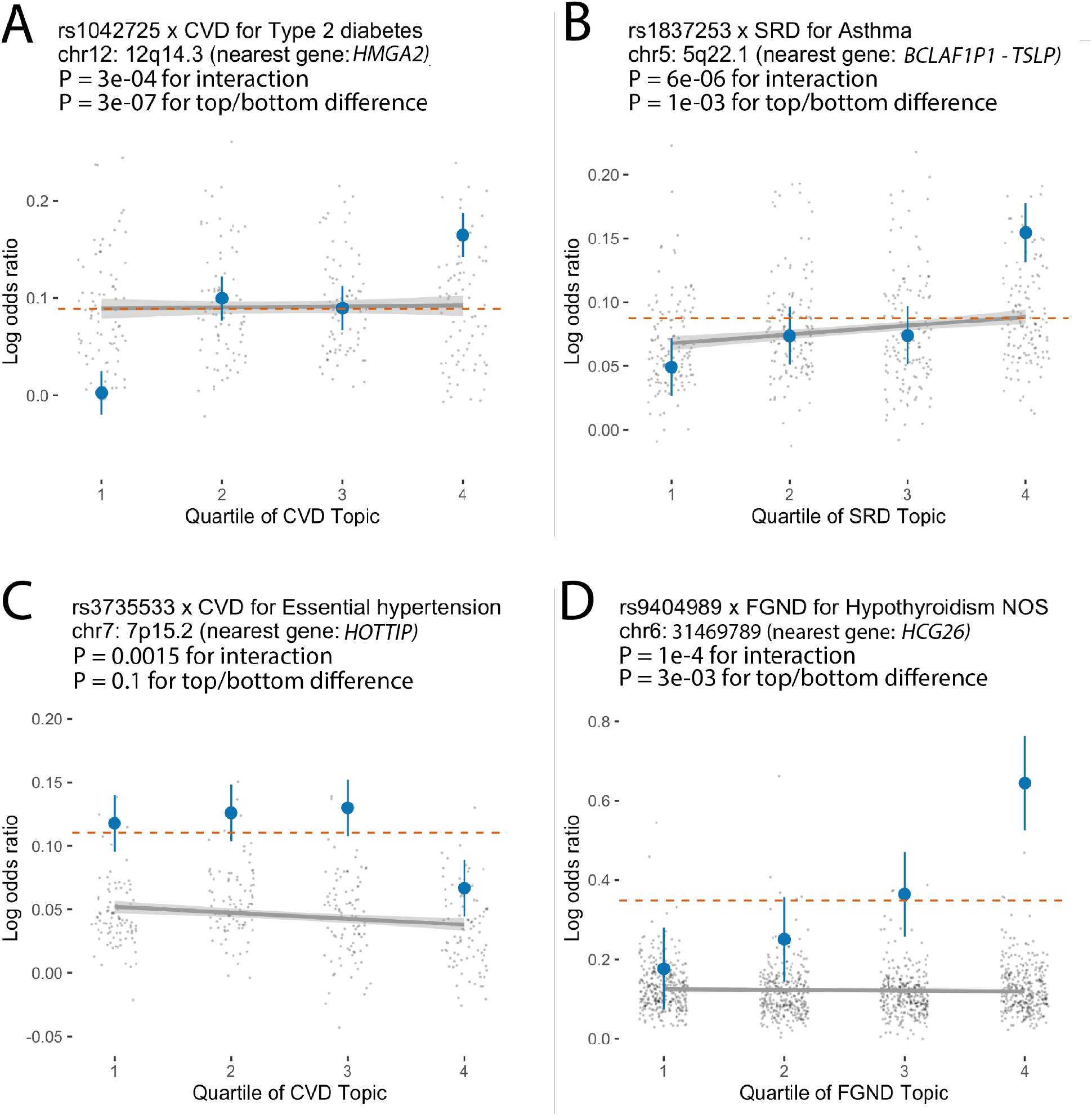
Examples of SNP x topic interaction effects on disease phenotypes. For each example, we report main SNP effects (log odds ratios) specific to each quartile of topic weights across individuals, for both the focal SNP (blue dots) and background SNPs for that disease and topic (genome-wide significant main effect (*P* < 5 × 10^!U^) but non-significant SNP x topic interaction effect (*P* > 0.05); grey dots). Dashed red lines denote aggregate main SNP effects for each focal SNP. Error bars denote 95% confidence intervals. Grey lines denote linear regression of grey dots, with grey shading denoting corresponding 95% confidence intervals. Numerical results are reported in Supplementary Table 18.

Disease topics capture known biology as well as the age-dependency of comorbidities for the same diseases. For example, early onset of essential hypertension is associated with the CVD topic ^43^, which captures the established connection between lipid dysfunction (“hypercholesterolemia”) and cardiovascular diseases^44^, while later onset of essential hypertension is associated with the CER topic, which pertains to type 2 diabetes, obesity and COPD (Figure 4A). Continuously varying age-dependent topic loadings for all 10 topics, restricted to diseases with high topic loadings, are reported in Supplementary Figure 13 and Supplementary Table 5. We note that most diseases have their topic loadings concentrated into a single topic (Figure 4B, Supplementary Figure 14A and Supplementary Table 4), and that most individuals have their topic weights concentrated into 1-2 topics (Figure 4C and Supplementary Figure 14B). For diseases spanning multiple topics (Supplementary Figure 6 and Supplementary Table 4), the assignment of type 2 diabetes patients to the CVD topic is consistent with known pathophysiology and epidemiology ^45, 46^ and has been shown in other comorbidity clustering studies, e.g. with the Beta Cell and Lipodystrophy subtypes described in ref. ^35^ and the severe insulin-deficient diabetes (SIDD) subtype described in ref.^14^, which are characterised by early onset of type 2 diabetes and have multiple morbidities including hypercholesterolemia, hyperlipidemia, and cardiovascular diseases ^47^. In addition, early-onset breast cancer and late-onset breast cancer are associated with different topics, e.g. NRI and FGND, consistent with known treatment effects for breast cancer patients which increase susceptibility to infections, especially bacterial pneumonias^48^ and hypothyroidism ^49^. We conclude that ATM identifies latent disease topics that robustly compress age-dependent comorbidity profiles and capture disease comorbidities both within and across Phecode systems.

### Age-dependent disease topic loadings capture concordant comorbidity profiles in All of Us

To assess the transferability of inferred topics between cohorts, we applied ATM to longitudinal data from 211,908 All of Us samples^30^. We analysed 3,098,771 diagnoses spanning 233 of the 348 diseases analysed in UK Biobank for which data were available; the average individual had 14.6 disease diagnoses, and an average disease had a standard deviation of 14.0 years in age at diagnosis. The optimal model for All of Us included 13 topics (Supplementary Figure 15, Supplementary Figure 16A-B, and Supplementary Table 6); most diseases have their topic loadings concentrated into a single topic and most individuals have their topic weights concentrated into 1-4 topics (Supplementary Figure 17).

We assessed the concordance between each UK Biobank topic and each All of Us topic by computing the correlation between the respective topic loadings across the 233 diseases analysed in both data sets. Results are reported in Figure 5A-B, Supplementary Figure 18, and Supplementary Table 7. The median correlation between the ten UK Biobank topics and the most similar All of Us topic was 0.54, confirming qualitative alignment of topic loadings between All of Us and UK Biobank. For example, the topic loadings of CVD and CER topics were qualitatively similar to the most similar All of Us topics (Figure 5A vs. Figure 4A), even though disease prevalences differ between the two cohorts (Supplementary Table 8). When using the optimal All of Us model (13 topics) to predict diagnoses in UK Biobank, we obtained a prediction odds ratio that was significantly larger than 1 (mean = 1.32; jackknife s.e. = 0.0027, Supplementary Figure 16C). We note 3 key differences between All of Us and UK Biobank data: (i) All of Us contains primary care and hospital data encoded using SNOMED clinical terms, whereas UK Biobank uses hospitalization episode statistics (HES; encoded using ICD-10 clinical terms); (ii) All of Us is based on the U.S. population and U.S. health care system whereas UK Biobank is based on the UK population and UK health care system, which impacts diagnostic criteria and age at diagnosis; and (iii) All of Us individuals have different ancestries and socioeconomic backgrounds (including 26% African and 17% Latino; 78% of All of Us represents groups historically underrepresented in biomedical research based on race, ethnicity, age, gender identity, disability status, medical care access, income, and educational attainment) than UK Biobank individuals (94% European ancestry with higher than average income and educational attainment). We consider the cross-cohort prediction odds ratio of 1.32 to be an encouraging result given these key differences.

For each of the 233 diseases, we assessed the concordance between UK Biobank and All of Us topic assignments for that disease by computing the correlation between UK Biobank topic assignments and All of Us topic assignments that were mapped to UK Biobank topics (by weighting by correlations between topics; Methods). The average correlation between UK Biobank and All of Us topic assignments for the same disease was 0.70 (vs. average correlation of 0.02 for different diseases) (Figure 5C, Supplementary Figure 19, and Supplementary Table 9). We conclude that ATM identifies latent disease topics from the All of Us cohort that align with topics from UK Biobank.

### Disease subtypes defined by distinct topics are genetically heterogeneous

We sought to define disease subtypes in UK Biobank data based on the topic weights of each patient and diagnosis-specific topic probabilities of each disease diagnosis. In some analyses, we used continuous-valued topic weights to model disease subtypes. In analyses that require discrete subtypes, we assigned a discrete topic assignment to each disease diagnosis based on its maximum diagnosis-specific topic probability, and inferred the *comorbidity-derived subtype* of each disease diagnosis based on the discrete topic assignment; we note that discretizing continuous data loses information (see Discussion).We restricted our disease subtype analyses to 52 diseases with at least 500 diagnoses assigned to each of two discrete subtypes; the average correlation between UK Biobank and All of Us disease subtypes (see above; same metric as Figure 5C) was 0.64 for the 41 (of 52) diseases that were shared between the two cohorts (Table 2, Methods, Supplementary Figure 6, Supplementary Figure 20, and Supplementary Table 10).

**Table 2.**
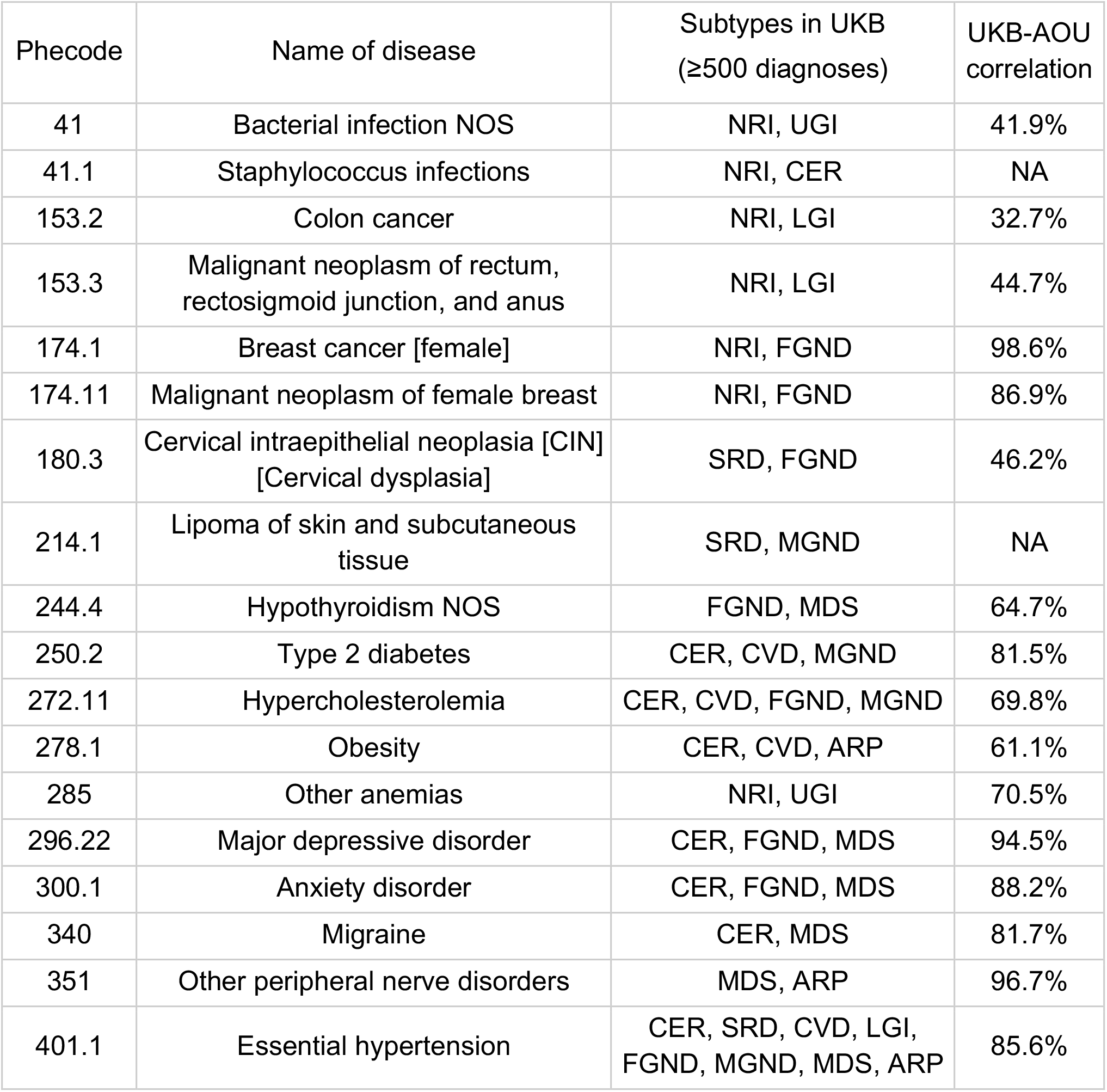
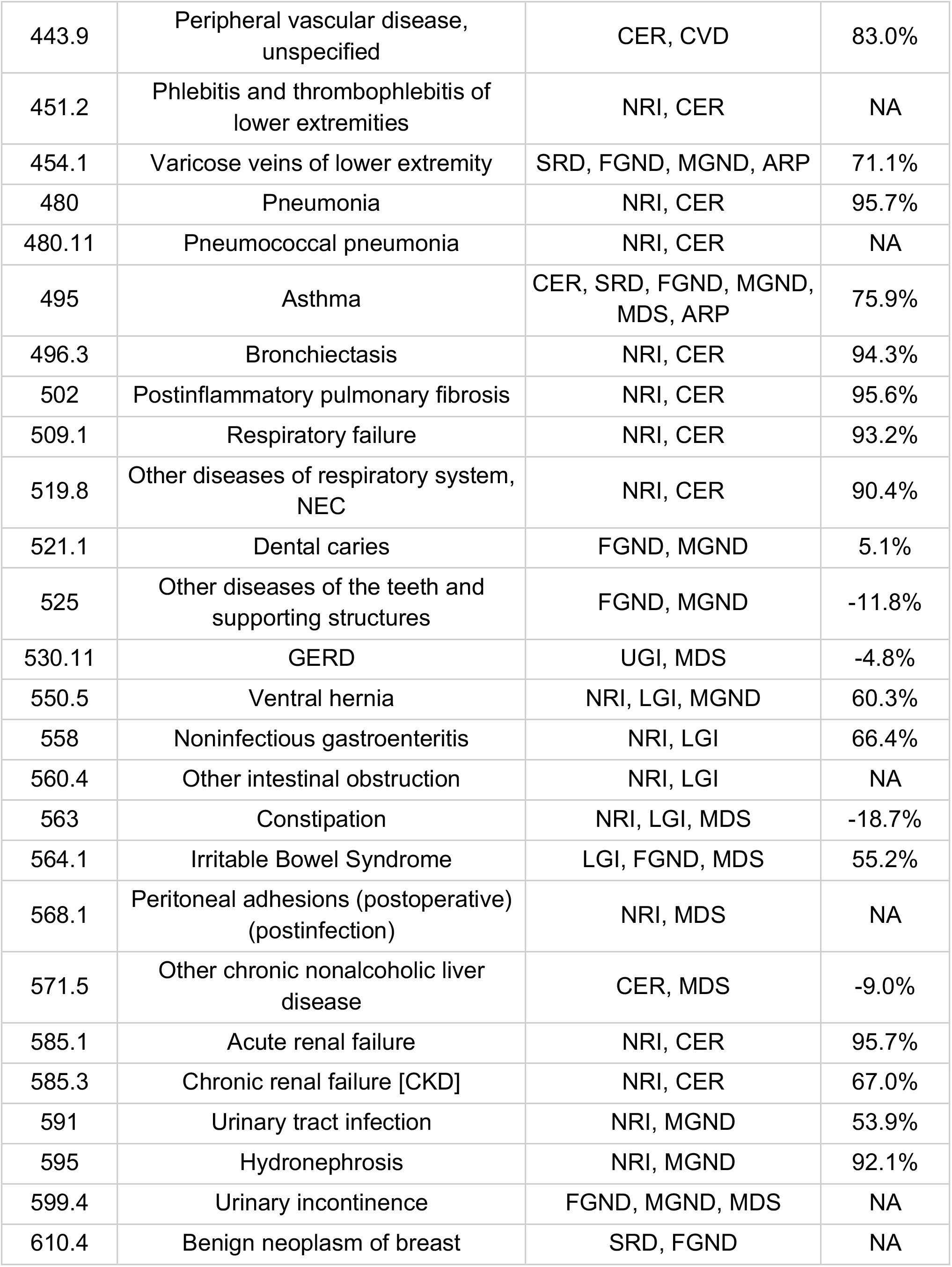
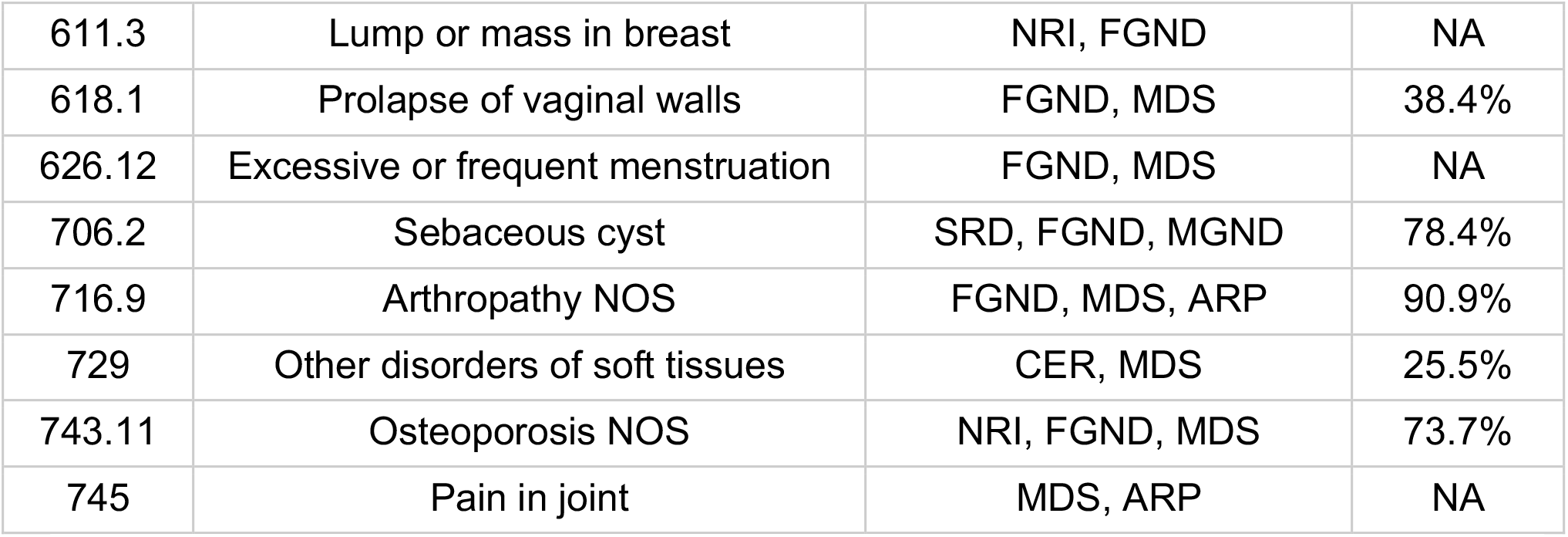
List of 52 diseases with comorbidity subtypes in UK Biobank. For each disease with at least 500 diagnoses assigned to each of two discrete subtypes, we list its Phecode, disease description, subtypes with at least 500 diagnoses, and correlations between UK Biobank topic assignments and All of Us topic assignments that were mapped to UK Biobank topics (Methods).

| Phecode | Name of disease | Subtypes in UKB<br>(≥500 diagnoses) | UKB-AOU<br>correlation |
| --- | --- | --- | --- |
| 41 | Bacterial infection NOS | NRI, UGI | 41.9% |
| 41.1 | Staphylococcus infections | NRI, CER | NA |
| 153.2 | Colon cancer | NRI, LGI | 32.7% |
| 153.3 | Malignant neoplasm of rectum, rectosigmoid junction, and anus | NRI, LGI | 44.7% |
| 174.1 | Breast cancer [female] | NRI, FGND | 98.6% |
| 174.11 | Malignant neoplasm of female breast | NRI, FGND | 86.9% |
| 180.3 | Cervical intraepithelial neoplasia [CIN]<br>[Cervical dysplasia] | SRD, FGND | 46.2% |
| 214.1 | Lipoma of skin and subcutaneous tissue | SRD, MGND | NA |
| 244.4 | Hypothyroidism NOS | FGND, MDS | 64.7% |
| 250.2 | Type 2 diabetes | CER, CVD, MGND | 81.5% |
| 272.11 | Hypercholesterolemia | CER, CVD, FGND, MGND | 69.8% |
| 278.1 | Obesity | CER, CVD, ARP | 61.1% |
| 285 | Other anemias | NRI, UGI | 70.5% |
| 296.22 | Major depressive disorder | CER, FGND, MDS | 94.5% |
| 300.1 | Anxiety disorder | CER, FGND, MDS | 88.2% |
| 340 | Migraine | CER, MDS | 81.7% |
| 351 | Other peripheral nerve disorders | MDS, ARP | 96.7% |
| 401.1 | Essential hypertension | CER, SRD, CVD, LGI, FGND, MGND, MDS, ARP | 85.6% |
| 443.9 | Peripheral vascular disease, unspecified | CER, CVD | 83.0% |
| 451.2 | Phlebitis and thrombophlebitis of lower extremities | NRI, CER | NA |
| 454.1 | Varicose veins of lower extremity | SRD, FGND, MGND, ARP | 71.1% |
| 480 | Pneumonia | NRI, CER | 95.7% |
| 480.11 | Pneumococcal pneumonia | NRI, CER | NA |
| 495 | Asthma | CER, SRD, FGND, MGND, MDS, ARP | 75.9% |
| 496.3 | Bronchiectasis | NRI, CER | 94.3% |
| 502 | Postinflammatory pulmonary fibrosis | NRI, CER | 95.6% |
| 509.1 | Respiratory failure | NRI, CER | 93.2% |
| 519.8 | Other diseases of respiratory system, NEC | NRI, CER | 90.4% |
| 521.1 | Dental caries | FGND, MGND | 5.1% |
| 525 | Other diseases of the teeth and supporting structures | FGND, MGND | -11.8% |
| 530.11 | GERD | UGI, MDS | -4.8% |
| 550.5 | Ventral hernia | NRI, LGI, MGND | 60.3% |
| 558 | Noninfectious gastroenteritis | NRI, LGI | 66.4% |
| 560.4 | Other intestinal obstruction | NRI, LGI | NA |
| 563 | Constipation | NRI, LGI, MDS | -18.7% |
| 564.1 | Irritable Bowel Syndrome | LGI, FGND, MDS | 55.2% |
| 568.1 | Peritoneal adhesions (postoperative) (postinfection) | NRI, MDS | NA |
| 571.5 | Other chronic nonalcoholic liver disease | CER, MDS | -9.0% |
| 585.1 | Acute renal failure | NRI, CER | 95.7% |
| 585.3 | Chronic renal failure [CKD] | NRI, CER | 67.0% |
| 591 | Urinary tract infection | NRI, MGND | 53.9% |
| 595 | Hydronephrosis | NRI, MGND | 92.1% |
| 599.4 | Urinary incontinence | FGND, MGND, MDS | NA |
| 610.4 | Benign neoplasm of breast | SRD, FGND | NA |
| 611.3 | Lump or mass in breast | NRI, FGND | NA |
| 618.1 | Prolapse of vaginal walls | FGND, MDS | 38.4% |
| 626.12 | Excessive or frequent menstruation | FGND, MDS | NA |
| 706.2 | Sebaceous cyst | SRD, FGND, MGND | 78.4% |
| 716.9 | Arthropathy NOS | FGND, MDS, ARP | 90.9% |
| 729 | Other disorders of soft tissues | CER, MDS | 25.5% |
| 743.11 | Osteoporosis NOS | NRI, FGND, MDS | 73.7% |
| 745 | Pain in joint | MDS, ARP | NA |

Age-dependent distributions of *comorbidity-derived* subtypes for four diseases (type 2 diabetes, asthma, hypercholesterolemia, and essential hypertension) are reported in Figure 6A and Supplementary Table 11; results for all 52 diseases are reported in Supplementary Figure 21 and Supplementary Table 11, and age-dependent distributions for the same four diseases in All of Us are reported in Supplementary Figure 22. The number of subtypes can be large, e.g. six subtypes for essential hypertension. Subtypes are often age-dependent, e.g. for the CVD and MGND subtypes of type 2 diabetes^14, 35^ (discussed above).

ATM and the resulting subtype assignments do not make use of genetic data. However, we used genetic data to assess genetic heterogeneity across inferred subtypes of each disease. We used continuous-valued topic weights in this analysis. We first assessed whether PRS for overall disease risk varied with continuous-valued topic weights for each disease; PRS were computed using BOLT-LMM with five-fold cross validation ^50, 51^ (see Methods and Code Availability). Results for four diseases (from Figure 6A) are reported in Figure 6B and Supplementary Table 12; results for all 10 well-powered diseases (10 of 52 diseases with highest z-scores for nonzero SNP-heritability) are reported in Supplementary Figure 23 and Supplementary Table 12. We identified 18 disease-topic pairs (of 10*10=100 disease-topic pairs analysed) for which PRS values in disease cases vary with patient topic weight. For example, for essential hypertension, hypercholesterolemia, and type 2 diabetes, patients assigned to the CVD subtype had significantly higher PRS values than patients assigned to other subtypes. For essential hypertension, patients assigned to the CER subtype had significantly higher PRS values; for type 2 diabetes, patients assigned to the CER subtype had lower PRS values than the CVD subtype, even though the majority of type 2 diabetes diagnoses are assigned to the CER subtype. We further verified that most of the variation in PRS values with disease subtype could not be explained by age^52^ or differences in subtype sample size (Supplementary Figure 24). These associations between subtypes (defined using comorbidity data) and PRS (defined using genetic data) imply that disease subtypes identified through comorbidity are genetically heterogeneous, consistent with phenomenological differences in disease aetiology.

We further investigated whether subtype assignments (defined using comorbidity data) revealed subtype-specific excess genetic correlations. We used discrete subtypes in this analysis. We estimated excess genetic correlations between disease-subtype and subtype-subtype pairs (relative to genetic correlations between the underlying diseases). Excess pairwise genetic correlations for 15 diseases and disease subtypes (spanning 11 diseases and 3 topics: CER, MGND and CVD) are reported in Figure 7A and Supplementary Table 13 (relative to genetic correlations between the underlying diseases; Figure 7B), and excess pairwise genetic correlations for all 89 well-powered diseases and disease subtypes (89 of 378 diseases and disease subtypes with z-score > 4 for nonzero SNP-heritability; 378 = 348 diseases + 30 disease subtypes) are reported in Supplementary Figure 25 and Supplementary Table 13. Genetic correlations between pairs of subtypes involving the same disease were significantly less than 1 (FDR<0.1) for hypertension (CER vs. CVD: *ρ* = 0.86 ± 0.04, P=0.0004; MGND vs. CVD: *ρ* = 0.74 ± 0.05, P=3× 10^-8^) and type 2 diabetes (CER vs. MGND: *ρ* = 0.64 ± 0.09, P=8 × 10^-5^) (Figure 7A; Supplementary Table 13). In addition, we observed significant excess genetic correlations (FDR<0.1) for 8 disease-subtype and subtype-subtype pairs involving different diseases (Figure 7A; Supplementary Table 13).We sought to verify that genetic differences between subtypes were not due to partitions of the cohort that are unrelated to disease (e.g. we expect a nonzero genetic correlation between tall vs. short type 2 diabetes cases, even if height is not genetically correlated to type 2 diabetes). Thus, we assessed whether the excess genetic correlation could be explained by non-disease-specific differences in the underlying topics (which are weakly heritable; Supplementary Table 3) by repeating the analysis using disease cases and controls with matched topic weights (i.e. case and controls have matched topic weights distributions within each disease or disease subtypes) (Methods). We determined that the excess genetic correlations could not be explained by non-disease-specific differences (Supplementary Figure 26). We also estimated subtype-specific SNP-heritability and identified some instances of differences between subtypes, albeit with limited power (Supplementary Table 14).

Finally, we used the population genetic parameter *F*_ST_^53, 54^ to quantify genome-wide differences in allele frequency between two subtypes of the same disease. We used discrete subtypes in this analysis. We wished to avoid inferring genetic differences between subtypes that were due to partitions of the cohort that are unrelated to disease (e.g. we expect a nonzero *F*_ST_ between tall vs. short type 2 diabetes cases; see above). Thus, we assessed the statistical significance of nonzero *F*_ST_ estimates by comparing the observed *F*_ST_ estimates (between two subtypes of the same disease) to the expected *F*_ST_ based on matched topic weights (i.e. *F*_ST_ estimates between two sets of healthy controls with topic weight distributions matched to the respective disease subtypes) (excess *F*_ST_; Methods). We determined that 63 of 104 pairs of disease subtypes involving the same disease (spanning 29 of 49 diseases, excluding 3 diseases that did not have enough controls with matched topic weights) had significant excess *F*_ST_ estimates (FDR < 0.1) (Supplementary Figure 27, Supplementary Table 15). For example, the CVD, CER, and MGND subtypes of type 2 diabetes had significant excess *F*_ST_ estimates (*F*-statistic=0.0003, *P*=0.001 based on 1,000 matched control sets). This provides further evidence that disease subtypes as determined by comorbidity have different molecular and physiological aetiologies. We conclude that disease subtypes defined by distinct topics are genetically heterogeneous.

### Disease-associated SNPs have subtype-dependent effects

We hypothesised that disease genes and pathways might differentially impact the disease subtypes identified by ATM. We investigated the genetic heterogeneity between disease subtypes at the level of individual disease-associated variants. We used continuous-valued topic weights in this analysis. We employed a statistical test that tests for SNP x topic interaction effects on disease phenotype in the presence of separate SNP and topic effects (Methods). We verified via simulations that this statistical test is well-calibrated under a broad range of scenarios with no true interaction, including direct effect of topic on disease, direct effect of disease on topic, pleiotropic SNP effects on disease and topic, and nonlinear effects (Supplementary Figure 28). We also assessed the power to detect true interactions (Supplementary Figure 29). To limit the number of hypotheses tested, we applied this test to independent SNPs with genome-wide significant main effects on disease (Methods). We thus performed 2,530 statistical tests spanning 888 disease-associated SNPs, 14 diseases, and 35 disease subtypes (Supplementary Table 16). We assessed statistical significance using global FDR<0.1 across the 2,530 statistical tests. We also computed main SNP effects specific to each quartile of topic weights across individuals and tested for different odds ratios in top vs. bottom quartiles, as an alternative way to represent SNP x topic interactions; the top/bottom quartile test is more intuitive, but less powerful in most cases.

We identified 43 SNP x topic interactions at FDR<0.1 (Figure 8, Supplementary Figure 30, Supplementary Table 17 and Supplementary Table 18). Here, we highlight a series of examples. First, the type 2 diabetes-associated SNP rs1042725 in the *HMGA2* locus has a higher odds ratio in the top quartile of CVD topic weight (1.18±0.02) than in the bottom quartile (1.00±0.02) (P=3 × 10^-4^ for interaction test (FDR = 0.04 < 0.1); P =3 × 10^-7^ for top/bottom quartile test (FDR = 0.0002 < 0.1)). *HMGA2* is associated with type 2 diabetes^55^ and is reported to have functions in cardiac remodelling^56^, suggesting that shared pathways underlie the observed SNP x topic interaction. Second, the asthma-associated SNP rs1837253 in the *TSLP* locus has a higher odds ratio in the top quartile of SRD topic weight (1.17±0.02) than in the bottom quartile (1.05±0.02) (P = 6 × 10^-6^ for interaction test (FDR = 0.004 < 0.1); P=1 × 10^-3^ for top/bottom quartile test (FDR = 0.08 < 0.1)). *TSLP* plays an important role in promoting Th2 cellular responses and is considered a potential therapeutic target, which is consistent with assignment of asthma and atopic/contact dermatitis^57^ to the SRD topic (Supplementary Table 4). Third, the hypertension-associated SNP rs3735533 within the *HOTTIP* long non-coding RNA has a lower odds ratio in the top quartile of CVD topic weight (1.07±0.02) than in the bottom quartile (1.13±0.02) (P = 0.0015 for interaction test (FDR = 0.09 < 0.1); P=0.1 for top/bottom quartile test (FDR = 0.55)). *HOTTIP* is associated with blood pressure^27, 58^ and conotruncal heart malformations^59^. Fourth, the hypothyroidism-associated SNP rs9404989 in the *HCG26* long non-coding RNA has a higher odds ratio in the top quartile of FGND topic weight (1.90±0.24) than in the bottom quartile (1.19±0.13) (P = 1 × 10^-4^ for interaction test (FDR = 0.02 < 0.1);

P=3 × 10^-3^ for top/bottom quartile test (FDR = 0.15)). Hypothyroidism associations have been reported in the HLA region^27^, but not to our knowledge in relation to the *HCG26*. To verify correct calibration, we performed control SNP x topic interaction tests using the same 888 disease-associated SNPs together with random topics that did not correspond to disease subtypes, and confirmed that these control tests were well-calibrated (Supplementary Figure 31B). We conclude that genetic heterogeneity between disease subtypes can be detected at the level of individual disease-associated variants.

## Discussion

We have introduced an age-dependent topic modelling (ATM) method to provide a low-rank representation of longitudinal disease records, leveraging age-dependent comorbidity profiles to identify and validate biological subtypes of disease. Our study builds on previous studies on topic modelling^37, 38, 40, 60^, genetic subtype identification^13–15^, and low-rank modelling of multiple diseases to identify shared genetic components^25–27^. We highlight three specific contributions of our study. First, we incorporated age at diagnosis information into our low-rank representation, complementing the use of age information in other contexts^32, 52, 61^; we showed that age information is highly informative for our inferred comorbidity profiles in both simulated and empirical data, emphasising the importance of accounting for age in efforts to classify disease diagnoses. Second, we identified 52 diseases with heterogeneous comorbidity profiles that we used to define disease subtypes, many of which had not previously been identified (Supplementary Table 19); comorbidity-derived disease subtypes were consistent between UK Biobank and All of Us, despite key differences between these cohorts. Third, we used genetic data (including PRS, genetic correlation and *F*_ST_ analyses) to validate these disease subtypes, confirming that the inferred subtypes reflect true differences in disease aetiology.

We emphasise three downstream implications of our findings. First, it is of interest to perform disease subtype-specific GWAS on the disease subtypes that we have identified here, analogous to GWAS of previously identified disease subtypes^13–15^. Second, our findings motivate efforts to understand the functional biology underlying the disease subtypes that we identified; the recent availability of functional data that is linked to EHR is likely to aid this endeavor^29, 62^. Third, the efficient inference of ATM permits identifying age-dependent comorbidity profiles and disease subtypes in much larger EHR data sets^63^;though we acknowledge that establishing comprehensive representations of disease topics that are transferable and robust across different healthcare systems and data sources represents a major future challenge.

Our findings reflect a growing understanding of the importance of context, such as age, sex, socioeconomic status and previous medical history, in genetic risk ^52, 64, 65^. To maximise power and ensure accurate calibration, context information needs to be integrated into clinical risk prediction tools that combine genetic information (such as polygenic risk scores ^1,66^) and non-genetic risk factors. Our work focuses on age, but motivates further investigation of other contexts. We note that aspects of context are themselves influenced by genetic risk factors, hence there is an open and important challenge in determining how best to combine medical history and/or causal biomarker measurements with genetic risk to predict future events^67^.

We note several limitations of our work. First, age at diagnosis information in EHR data may be an imperfect proxy for true age at onset, particularly for less severe diseases that may be detected as secondary diagnoses; although perfectly accurate age at onset information would be ideal, our study shows that that imperfect age at diagnosis information is sufficient to draw meaningful conclusions. Second, raw EHR data may be inaccurate and/or difficult to parse^1^; again, although perfectly accurate EHR data would be ideal, our study shows that imperfect EHR data is sufficient to draw meaningful conclusions. Third, our ATM approach incurs substantial computational cost (Supplementary Table 20); however, analyses of biobank-scale data sets are computationally tractable, with our main analysis requiring only 4.7 hours of running time.

Fourth, the genetic correlation and *F*_ST_ analyses were based on discrete subtypes, but discretizing continuous data loses information and may compromise power. However, definitions of disease often discretize continuous variables^68^. In addition, our PRS analysis (Figure 6) and SNP x topic interaction analysis (Figure 8) leveraged continuous-valued topic weights. Fifth, interpretability can be a potential downside of data reduction approaches. The interpretation of a particular disease topic is that it consists of diseases that tend to co-occur with a specified set of diseases as a function of age. Identifying the functional biology underlying these co-occurrences remains a direction for future research, but there is immediate utility in performing disease subtype-specific GWAS and downstream analyses using the subtypes that we have identified. Despite these limitations, ATM is a powerful approach for identifying age-dependent comorbidity profiles and disease subtypes.

## Data Availability

All data produced in the present work are contained in the manuscript

## Acknowledgements

This research has been conducted using the UK Biobank Resource; application number 12788.

Funded by Wellcome (BST00080-H503.01 to XJ, 100956/Z/13/Z to GM, https:// wellcome.org); the Li Ka Shing Foundation (to GM, https://www.lksf.org); NIH grants R01 HG006399, R01 MH101244, and R37 MH107649 (to ALP); The Alan Turing Institute (https://www.turing.ac.uk), Health Data Research UK (https://www.hdruk.ac.uk), the Medical Research Council UK (https://mrc.ukri.org), the Engineering and Physical Sciences Research Council (EPSRChttps://epsrc.ukri.org) through the Bayes4Health programme Grant EP/R018561/1, and AI for Science and Government UK Research and Innovation (UKRI, https://www.turing.ac.uk/ research/asg) (to CH); BHF Chair award CH/12/2/29428 (to XJ). This work was supported by core funding from the: British Heart Foundation (RG/13/13/30194; RG/18/13/33946), BHF Cambridge Centre of Research Excellence (RE/13/6/30180) and NIHR Cambridge Biomedical Research Centre (BRC-1215-20014). The funders had no role in study design, data collection and analysis, decision to publish, or preparation of the manuscript.

This work uses data provided by patients and collected by the NHS as part of their care and Support. Computation used the Oxford Biomedical Research Computing (BMRC) facility, a joint development between the Wellcome Centre for Human Genetics and the Big Data Institute supported by Health Data Research UK and the NIHR Oxford Biomedical Research Centre. The views expressed are those of the authors and not necessarily those of the NHS, the NIHR or the Department of Health. We thank Kushal Dey, Luke Kelly and Yunlong Jiao for the discussion.

## Data availability

UK Biobank data is publicly available at https://www.ukbiobank.ac.uk/.

## Code availability

Open-source software implementing the ATM method is available at https://github.com/Xilin-Jiang/ATM. BOLT-LMM is available at https://alkesgroup.broadinstitute.org/BOLT-LMM/. Heritability and genetic correlation analysis were performed using LDSC, which is available at https://github.com/bulik/ldsc. PLINK v1.9, which was used for *F_ST_* and association tests, is available at https://www.cog-genomics.org/plink/.

## Methods

### Age-dependent topic model (ATM)

Our Age-dependent topic model (ATM) is a Bayesian hierarchical model to infer latent risk profiles for common diseases. The model assumes that each individual possesses several age-evolving disease profiles (topic loadings), which summarise the risk over age for multiple diseases that tend to co-occur within an individual’s lifetime, namely the age specific multi-morbidity profiles. At each disease diagnosis, one of the disease profiles is first chosen based on individual weights of profile composition (topic weights), the disease is then sampled from this profile conditional on the age of the incidence.

We constructed a Bayesian hierarchical model to infer *K* latent risk profiles for *D* distinct common diseases. Each latent risk profile (comorbidity topics) is age-evolving and contains risk trajectories for all *D* diseases considered. Each individual might have a different number of diseases, while the disease risk is determined by the weighted combination of latent risk topics. The indices in this note are as follows:

● *s* = *1*, . . ., *M*;
● *n* = *1*, . . ., *N*_*s*_;
● *i* = *1*, . . ., *K*;
● *j* = *1*, . . ., *D*;

where *M* is the number of subjects, *N_s_* is the number of records within *s^th^* subject, *K* is the number of topics, and *D* is the total number of diseases we are interested in. The plate notation of the generative model is summarised in Supplementary Figure 1:

● *θ* ∈ *R* ^*M*^ ^×^ ^*K*^ is the topic weight for all individuals (referred to as patient topic weights), each row of which (∈ *R*^*k*^) is assumed to be sampled from a Dirichlet distribution with parameter *α*. *α* is set as a hyper parameter: *θ*_*s*_ ∼ *Dir*(*α*). We used topic weights to assign continuous values for disease subtypes in PRS and SNP x Topic analyses.
● *z* ∈ {*1*,*2*, . . ., *K*}^∑s *N*s^ (referred to as diagnosis-specific topic probability) is the topic assignment for each diagnosis W ∈ {*1*,*2*, . . ., *D*}^∑s *N*s^ . Note the total number of diagnoses across all patients are ∑_s_ *N*_s_. The topic assignment for each diagnosis is generated from a categorical distribution with parameters equal to *s^th^* individual topic weight: *z*_*sn*_ ∼ *Multi*(*θ*_*s*_) . We used diagnosis-specific topic probability to define discrete disease subtypes in excess genetic correlation and excess *F*_ST_ analyses.
● *β*(*t*) ∈ *F*(*t*)^*K*^ ^×^ ^*D*^ is the topic loading which is *K* × *D* functions of age *t. F*(*t*) is the class of functions of *t*. At each plausible *t*, the following is satisfied: ∑_*j*_ *β*_*ij*_(*t*) = *1* . In practice we ensure above is true and add smoothness by constrain *F*(*t*) to be a softmax of spline or polynomial functions: 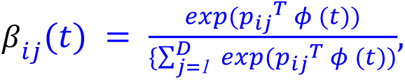, where 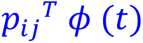 polynomial and spline functions of *t*; *p_ij_* = { *p_ijd_*}; *d* = *1*,*2*, . . ., *P* ; *P* is the degree of freedom that controls the smoothness; *ϕ* (*t*) is polynomial and spline basis for age *t*.
● W ∈ {*1*,*2*, . . ., *D*}^∑s *N*s^ are observed diagnoses. The *n*^*t*^*^ℎ^* diagnosis of *s^th^* individual W_*sn*_ is sampled from the topic *β*_*zsn*_ (*t*) chosen by *z*_*sn*_: W_*sn*_ ∼ *Multi*(*β*_*zsn*_(*t*_*sn*_)), here *t*_*sn*_ is the age of the observed age at diagnosis of the observed diagnosis W_*sn*_.

The values of interest in this model are global topic parameter *β*, individual (patient) level topic weight *θ*, and diagnosis-specific topic probability *z*. Based on the generative process above, we notice that each patient is independent conditional on *α*. Therefore, the inference of *θ* and (discussed below) is performed by looping each individual in turn.

The key element in our model is age-evolving risk profiles, which is achieved by model the comorbidity trajectories *β*(*t*) ∈ *F*(*t*)^*K*^ ^×^ ^*D*^ as functions of age. The functionals *F*(*t*) are parameterized as linear, quadratic, cubic polynomials, and cubic splines with one, two and three knots. We use prediction odds ratio to decide the optimal model structure including the function forms and the number of topics; we use ELBO to choose the optimal inference results (with random parameter initialization) for the same model structure(Supplementary Table 1).

## Inference of ATM

The variables of interest are global topic parameter *β*(*t*), individual (patient) level topic weight *θ*, and diagnosis-specific topic probability *z* of each diagnosis. We could adopt an EM strategy, where in the E-step we first estimate posterior distribution of *θ* and *z*, then in the M-step we estimate *β* which maximises the evidence lower bound (ELBO).

The details of the inference is explained in Supplementary Note. In summary, in a Bayesian setting, We used the evidence function (W|*α*, *β*) to evaluate how well the model fits the data. The best β(*t*) is found by maximise the evidence function, while for *θ* and *z* we aim to find or approximate their posterior distribution *p*(*z*, *θ* | W, *α*, *β*) . Given that the posterior distribution is intractable, we use variational distribution (*z*, *θ*) to approximate them. Now we could write the evidence function as:

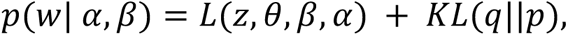

here 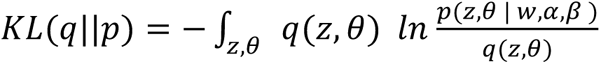 is the KL divergence. Since KL divergence is always positive, (*z*, *θ*, *β*, *α*) is a lower bound of the evidence function:

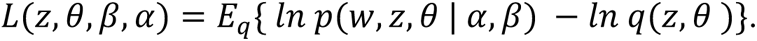

When finding the posterior of *θ* and *z*, we want *ln q*(*z*, *θ*) to be as close to the posterior*p*(*z*, *θ* | *W, α*, *β*) as possible. Since *KL*(*q*||*p*) = *0* when *q*(*z*, *θ*) = *p*(*z*, *θ* | W, *α*, *β*), this is achieved by minimising *KL*(*q*||*p*) or maximise *L*(*z*, *θ*, *β*, *α*). The most commonly used form of (*z*, *θ*) assumes the distribution is factorised, which might cause instability when signal-to-noise ratio is low^69^. Therefore, more accurate inference methods such as collapsed variational inference is considered^40^. Comparison of the evidence lower bound (*z*, *θ*, *β*, *α*) shows collapsed variational inference (CVB) is consistently more accurate than mean-field variational inference (VB) (Supplementary Figure 10). Therefore we chose the collapsed variational inference^40^. The collapsed variational inference is achieved by integrate out *θ* from the likelihood function (W, *z*, *θ* | *α*, *β*) and find the approximated posterior distribution *q*(*z*). For detailed derivation, the comparison between collapsed variational inference and mean-field variational inference, and update algorithms, see Supplementary Note.

When finding the β(*t*) that maximises the evidence function, we again maximise *L*(*z*, *θ*, *β*, *α*). Maximising (*z*, *θ*, *β*, *α*) with respect to *β*(*t*) does not have an analytical solution due to its softmax structure. We use local variational methods and numeric optimisation to find the distribution of β(*t*). In summary, L(z, θ, β, α) is not tractable with respect to *β*(*t*) as it contains a log of softmax function (Section 3.2 of Supplementary Note). We introduced a local variational variable to obtain a tractable lower bound of L(z, θ, β, α) (equation 11 in Supplementary Note) and use gradient descent to approximate the lower bound. Details are provided in Supplementary Note.

We extract topic weights at patient-level and diagnosis-level from the posterior distribution inferred from the data. Our model has the desired property that each patient and patient-diagnosis are assigned to comorbidity topics. The model estimates the posterior distribution (*z*), which is a categorical distribution (equation 8 of Supplementary Note). We listed following definitions in this paper that are derived from the (*z*):

● Each patient-diagnosis (incident disease) has a *diagnosis-specific topic probability*, which is computed as *E_q_*{*z*_*n*_} .
● Each patient has a posterior *topic weights θ*, which is a dirichlet distribution *θ*_*s*_ ∼ 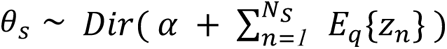. The *topic weights* of each patient is defined as the mode of this Dirichlet distribution 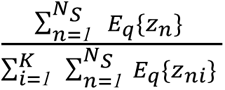 (we used *α* = *1*, which puts an noninformative prior on the topic weights). Topic weight is the low-rank representation of disease history, for analyses including PRS association with comorbidity topics and SNP x Topic interaction analysis.
● The *average topic assignments* of disease *j* is the mean over all incidences 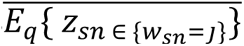. This metric is used to measure which comorbidity topic a disease is associated with (Figure 4B), and it is equivalent to a weighted average of topic loadings (Supplementary Note equation 5 shows the link between diagnosis-specific topic probability and topic loading). A disease assigned to multiple topics is considered to have comorbidity subtypes.
● A hard assignment of a patient-diagnosis to a *comorbidity-derived subtype* is based on the max value of the vector *E_q_*{*z*_*n*_} . The incident disease is assigned to topic *argmix_i_* (*E*_*q*_{*z*_*ni*_}) .

### Metrics for evaluating ATM

ATM is evaluated for different purposes, which requires different metrics (Supplementary Table 1). Here we list the details of the four metrics considered: *Prediction odds ratio, Evidence Lower Bound (ELBO), the Area under the Precision-Recall curve (AUPRC)*^70^*, and Co-occurrence odds ratio*.

#### Prediction odds ratio

To compare models of different topic numbers and configuration of age profiles, we compare the prediction odds ratio of each model. Briefly, prediction odds ratio is defined on 20% held-out test data as the odds that the true diseases are within the top 1% diseases predicted by ATM (trained on 80% of the training set and uses earlier diagnoses as input), divided by the odds that the true diseases are within the top 1% of diseases ranked by prevalence.

Specifically, we separate UK Biobank patients into a training set (80%) and a testing set (20%). On the training set, we estimate the comorbidity topic loadings. On the testing set, we fix the topic loadings and infer the patient topic weights to predict the next disease in chronological order. The topic loadings are estimated using the *n* diseases and compute the risk rank of diseases at the age of the *n*+1 disease. The odds ratio is computed by the odds of the *n*+1 disease being in the top 1% of diseases versus being in the top 1% most prevalent diseases. We use the top 1% most prevalent diseases instead of randomly chosen diseases as it represents a naive prediction model that predicts disease based on prevalence. The patient topic weights computation is in section Inference of ATM and the risk is computed as the linear combination of topics using topic weights as coefficients. We also compute the prediction odds ratio using the LDA model. We repeat the procedure for 10 times for each model configuration.

We compared the prediction odds ratio for topic number between 5 to 20, with linear, quadratic polynomial, cubic polynomial, and splines with one, two and three knots. We also compare the ATM model with the LDA model of topic number between 5 to 20.

#### Evidence Lower Bound (ELBO)

ELBO evaluated the accuracy of the variational inference method on a specific data set ^39^. The mathematical expression of ELBO for ATM is presented in equation 9 in the Supplementary Note. To find the best model that fits the entire dataset, we evaluate the ELBO for models with 19 choices of the number of topics: 5-20, 25, 30, and 50; 6 choices of age profiles configuration: linear, quadratic polynomial, cubic polynomial, and splines with one, two and three knots. Each model is run for 10 times with random initialisations. We choose the model that has the highest ELBO after converging.

#### AURPC

To evaluate whether a model could capture the comorbidity subtypes in simulation analysis, we compute the precision, recall, and area under precision-recall curve (AUPRC) to correctly classify disease diagnosis to be from the topic that it is generated from. The topic of each diagnosis is determined by diagnosis-specific topic probability. Note we could only evaluate AUPRC in simulations where the truth is known.

#### Co-occurrence odds ratio

To verify that the comorbidity profiles that the model captured are capturing diseases that are more likely to present within the same individual, we estimate the odds ratio of the disease duo, trio, quartet, and quintet that are captured by the topic versus that of random combinations. We divide the population into an 80% training set and a 20% testing set. We trained the ATM model with five random initialisations and kept the inference with the highest ELBO. Each disease is assigned to a topic by the highest average topic assignments. (section Inference of ATM) We focus on the top 100 diseases ranked by prevalence to avoid the combination being too rare to appear in the population. In the testing set, we computed the odds of individuals who have all diseases in the comorbidities versus the odds implied if all diseases are independent (computed as the product of disease prevalence). The odds ratio is computed for all combinations of duo, trio, quartet, and quintet that are assigned to the same topics. We perform the same analysis using PCA for comparison.

#### Simulations of ATM method

To test whether the algorithm could assign disease diagnosis to correct comorbidity profiles, we simulated disease from two disease topics within a population of 10,000, using following parameters:

● *M* = *10*,*000*;
● 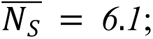
● 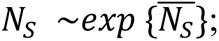
● *D* = *20*;
● *K* = *2*;

Here *M* is the number of individuals in the population, 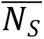 is the average number of diseases for each individual, *D* is the total number of diseases, is the number of comorbidity topics. The distribution of disease number per-individual *N*_s_is sampled from an exponential distribution, which matches those from UK Biobank data (Supplementary Figure 32). According to equation 3.1 in Ghorbani et al.^69^, whether the topic model could capture the true latent structure is determined by the information signal-to-noise ratio and could be evaluated with limits *M* → ∞; 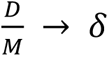, where *δ* is a constant. Therefore we choose *D* and *M* at scales that make 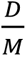 approximately similar to those of the UK Biobank dataset (Samples size = 282,957; distinct disease number = 349).

The simulated topics loadings are constructed as follows:

● All but *K* diseases are simulated to be associated with comorbidity profiles. Each of them has a risk period of 30 years and overlaps for 10 years with the next disease. For example, if disease 1 has a risk period from 30 to 59 years of age, disease 2 will have a risk period between 50 to 79 years of age. When the risk period reaches the maximal age, the truncated part will be carried to the next disease to create diseases with shorter risk period. All risk periods are assigned a value 1. The overlapping structure of topic loadings is chosen so that average standard deviation in age-at-diagnosis (8.5 years) and the age window under consideration (30-80 years of age) matches UK Biobank data.
● *K* diseases that are not associated with comorbidity are simulated to span all topics. The values of these diseases are sampled from 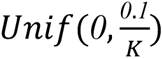 for each topic. Here *K* is the *K* number of topics.
● The age profiles are then normalised at each age point to ensure 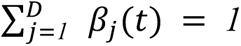 for all *t*. With this constraint we could sample a disease at each age *t* using a multinomial probability with the topic loading as the parameter. The age range of the simulated topics is 30 to 81 years of age, which is the minimal and maximal age at diagnosis of incident disease in the UK Biobank population. An example of a simulated topic is shown in Supplementary Figure 33.

For each individual, we sampled the Dirichlet parameter *α* from a gamma distribution (shape = 50, rate = 50). Topic loadings are sampled from the Dirichlet distribution for each patient as the generative process. For each patient, we first sample the number of diseases *N*_s_. For each incident disease, we sample the disease age from uniform distribution between age 30 to 81 and a topic from the topic loading. We then choose the incident disease based on the age at diagnosis from the chosen topic. The procedure follows the generative process described in Supplementary Note.

Since in real data we only use the first age at diagnosis for diseases that are recorded repeatedly within the same patient, we filter the simulated diseases accordingly. The filtered data are fed into the inference functions to infer the latent topics and disease assignments. The inferred topics resemble the true topics used to simulate diseases as shown in Supplementary Figure 33. For the initialisation of each inference, we first sample *β* and *θ* from the Dirichlet distribution of non-informative hyperparameters, then initialise other variables parameters following the generative process. The variational inference converged where the relative increase of ELBO is below *10^-6^*.

To simulate disease having distinct comorbidity subtypes, we first simulate diseases using the procedure described above. We consider two scenarios: (1) the subtype of diseases have the same age at diagnosis distribution. (2) the subtypes of disease have distinct age at diagnosis distribution.

We create diseases with distinct comorbidity profiles by combining diseases that are sampled from distinct topics and labelling them as a single disease. We first chose one disease (**disease A**) then sampled a proportion of a second disease (**disease B**) to label as **disease A**. The proportion is varied to create a different sample size ratio of the two subtypes. In scenario one, **disease B** is a disease that has the exact same age distribution as **disease A** but from the other topic. In scenario two, **disease B** is from the other topic and has a different age distribution (age at diagnosis moves up for 20 years, 10 years, or 5 years, respectively) than **disease A**. After changing the labels of **disease B** to be the same as **disease A**, we used the inference procedure described as above to get the posterior distribution.

To evaluate whether a model could capture the comorbidity subtypes, we compute the precision, recall, and area under precision-recall curve (AUPRC) to correctly classify incident **disease B** to be from the topic that it is generated from. The topic of each diagnosis is determined by diagnosis-specific topic probability. We use other diseases from the topic of **disease B** to benchmark the topic label. Topic modelling on the simulated data is performed with both ATM and LDA (both implemented using collapsed variational inference for fair comparison) to compare the performances.

We evaluate the subtype classification with varying values for three simulation parameters:

● ratio of sample sizes between the two subtypes. We change the ratio of the two subtypes by a grid between 0 to 0.9 with a step size 0.1. The default value of sample size ratio is set as 0.1 in other simulations to test for other parameters that have impacts on the precision and recall.
● Simulated population size. We simulated population sizes equal to 200, 500, 1000, 2000, 5000, and 10,000. The default population size is 10,000 in other simulations.
● Number of distinct diseases. We simulated datasets with 20, 30, 40, and 50 distinct diseases, with 2, 3, 4 and 5 underlying disease topics respectively. The default number of distinct diseases is 20 in other simulations.
● Difference of age distribution. We considered three scenarios of subtype age distribution, with 0, 10, and 20 years of difference in the average age at diagnosis.

### UK Biobank comorbidity data

We analysed comorbidity data from 282,957 UK Biobank samples with diagnoses for at least two of the 348 focal diseases that we studied (see below). We use the hospital episode statistics (HES) data within the UK Biobank dataset, which records diseases using the ICD-10/ICD-10CM coding system; the average record span of HES data is 28.6 years. Codes started with letters from A to N are kept as they correspond to disease code (opposed to procedure codes). The disease records were mapped from ICD-10/ICD-10CM codes to PheCodes using a three-step procedure: First, we mapped the first four letters of each ICD-10 records to the phecodes, using the map file downloaded from phewascatalog.org; second, we mapped the remaining records using ICD-10CM map file downloaded from phewascatalog.org; last, we mapped remaining records using the same ICD-10CM map system but only use the first four character of each ICD-10CM codes.

We also noticed (ICD-10/ICD-10CM)-Phecode pairs are not always one-to-one; when a single ICD-10/ICD-10CM code is mapped to more than one PheCodes, we chose the Phecode with the largest number of links to ICD-10/ICD-10CM codes to reduce redundancy of the mapping result. Using the procedure above, we mapped 99.7% ICD-10/ICD-10CM code to PheCodes, with 4,637,127 records in total.

The mapped Phecodes are filtered to keep only the first age at diagnosis for the same diseases within a patient. The age at diagnosis for each record is computed as the difference between month of birth to the episode starting date. We then computed the occurrence of each disease in the UK Biobank and kept 348 that have more than 1,000 occurrences (Supplementary Table 4). Starting with all 488,377 UK Biobank patients (including both European and non-European ancestries), we filtered the patients to keep only those who have at least two distinct diseases from the 348 focal diseases, as we are most interested in the comorbidity information. We treated the death as an additional disease (8,666 records) to evaluate if certain comorbidities are more likely to lead to fatal events. After these procedures, there are in total 1,726,144 distinct records across 282,957 patients.

To name the topics inferred from the UK Biobank, we take the sum of *average topic assignments* (section Inference of ATM) over diseases for each Phecode system and extract the three most common Phecode disease systems. Six topics are named using the three most common Phecod disease systems: NRI “neoplasms, respiratory, infectious diseases”, CER “cardiovascular, endocrine/metabolic, respiratory”, SRD “sense organs, respiratory, dermatologic”, FGND “female genitourinary, neoplasms, digestive”, MGND “male genitourinary, digestive, neoplasms”, MDS “musculoskeletal, digestive, symptoms”, and ARP. For four topics that are predominantly associated with one system, we name them based on their top associated Phecode system: LGI “lower gastrointestinal”, UGI “upper gastrointestinal”, CVD “cardiovascular”, and ARP “arthropathy”.

We present focal diseases for each topic in two ways. Firstly, we filter each topic using the profile mean value between age 30 to 81 to keep the top seven diseases. We chose seven for visualisation, as we found more diseases would be harder to read on a plot. Secondly, we also show seven diseases that have the highest average assignment to each topic. This will give a picture of diseases that are not the most prevalent in the population but are predominantly associated with the target topic.

To compare the comorbidity heterogeneity between age groups, we group the incidences for each disease to two age groups: young group (<60 years of age) and old group (≥60 years of age). We compute the average topic assignment of each group as described in section Inference of ATM. Additionally, we inferred topics for male (984,554 records in 156,366 individuals) and female (741,590 records in 126,591 individuals) populations respectively using a model with 10 topics and spline function with one knot. We extract the average topic assignment for each disease, and use Pearson’s correlation to match the topics for both sexes to the topics inferred on the entire population.

Each diagnosis is assigned to a specific topic using max diagnosis-specific topic probability. We focus our disease heterogeneity analysis on 52 diseases that have at least 500 incidences assigned to a secondary topic.

### All of Us comorbidity data

We analysed EHR data collected in the EHR domain of All of Us samples, which includes both primary care and secondary care data. The average distance between first and last diagnoses is 7.9 years (vs. 7.0 years in UK Biobank); the average record span period is unknown, but we hypothesized that it is likely to be considerably larger than 7.9 years (vs. 28.6 years in UK Biobank). Disease codes in the All of Us EHR domain are coded in SNOMED CT. We first mapped All of Us disease codes from SNOMED CT to ICD-10CM code using map version 20220901 downloaded from https://www.nlm.nih.gov/research/umls/mapping_projects/snomedct_to_icd10cm.html. When a single SNOMED CT code was mapped to multiple ICD-10CM codes, we choose the code with the highest UK Biobank prevalence from these ICD-10CM codes. We then mapped ICD-10CM codes to Phecodes, using the same procedure described in the section above. We kept 233 Phecodes that overlap with the 348 diseases analysed in the UK Biobank. We kept the first diagnosis for recurrent diseases in each patient. After mapping, we are left with 3,098,771 diagnoses spanning 211,908 All of Us samples. We run ATM with topic number from 5 to 20 and spline with two knots (degree of freedom = 5) on the All of Us comorbidity data and computed prediction odds ratio (using five-fold cross validation) and ELBO (on all 211,908 samples).

### Comparing disease topics between UK Biobank and All of Us

We compared the optimal models from UK Biobank (10 topics, degree of freedom = 5) and All of Us (13 topics, degree of freedom = 5). We constrained our analyses on 233 of the 348 diseases that are shared between the two data sets. We performed three analyses to compare the comorbidity patterns from the two data sets.

First, we computed the correlation of topic loadings from two data sets. Since the topic loadings are functions of age, we computed their correlations using four different ways to summarise age information: topic loadings averaged across age; topic loadings at age 50, 60, and 70. For each UK Biobank topic, we found its most similar All of Us topic that has max correlation of topic loadings (averaged across age).

Second, we computed the cross-population prediction odds ratio, using the All of Us topics to predict on UK Biobank comorbidity data. We divided the UK Biobank samples into 10 jackknife blocks and computed prediction odds ratios on each leave-one-out sample.

Third, we compared the correlation of comorbidity profiles (measured by average topic assignments; see Methods for definition) for 233 diseases that are shared between the two populations. We define *correlations between topic assignments* as the correlation between UK Biobank average topic assignments and All of Us average topic assignments after mapped to UK Biobank topic space (see below).

Comparing disease topics inferred from different data sets is challenging due to the exchangeability of topics (i.e. distinct topic configurations have the same likelihood for a given data set). To compute *correlations between topic assignments* from ATM inference on different populations, we first mapped the topics to the same topic space. Suppose there are two topic spaces {*T^1^*} and {*T^2^*}. We create a map (.) from {*T^1^*} and {*T^2^*} by computing the normalised *R^2^* between topic loadings:

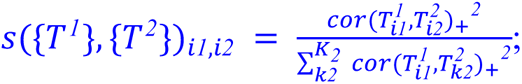

here 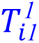 is the *i1*^*t*^*^ℎ^* topic loading from {*T^1^*}, 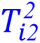 is the *i2*^*t*^*^ℎ^* topic loading from {*T^2^*}; *K_1_*, *K_2_* is the number of topics in {*T^1^*}, {*T^2^*}; *cor*(.)_+_ is the positive part of the correlation, where we set the negative correlations to zero as negative correlations between topic loadings are uninformative consequences of the multinomial distribution in the model. Intuitively, ({*T^1^*}, {*T^2^*})_i_*_1_*_,i*2*_ maps each 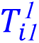 to {*T^2^*} based on the proportion of 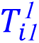 variance explained by the *K_2_* topics in {*T^2^*}. 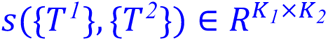.

Suppose we have the comorbidity profile of disease 1 and disease 2, which lies in {*T^1^*} and {*T^2^*} respectively. We could map disease 1 diagnosis-specific topic probability *z*_*sn*,_*_1_* ∈ *R*^*K*^*^1^* (or average topic assignments of disease 1; see these definitions in Methods) to topic space 2: *z′*_*sn*,*1*_ = *s*({*T^1^*}, {*T^2^*})^*T*^ *z*_*sn*,_*_1_*, *z′*_*sn*,*1*_ ∈ *R*^*K*^*^2^* . The correlations between topic assignments in topic space 2 between disease 1 and disease 2 is the correlation 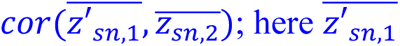 is the average topic assignments for disease 1 after mapped to topic space 2, which is the average of *z′*_*sn*,*1*_ across all diagnoses; 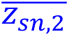 is the average topic assignments for disease 2. For correlations between topic assignments within the same topic, ({*T^1^*}, {*T^1^*}) is an identical matrix.

### UK Biobank genotype data

For genetic correlation analysis, *F*_ST_, and SNP x Topic interaction analyses, we used genetic data from 488,377 UK Biobank participants (prior to restricting to 282,957 samples with at least two of the 348 diseases studied). For PRS and heritability estimation of the 10 topics, we used mixed-effect association model implemented in BOLT-LMM software^50, 51^, where we constrained our analysis to 409,694 British Isle ancestry individuals to adjust for population structure. For *F*_ST_ analysis with PLINK we used 805,426 genotyped SNPs; for BOLT-LMM PRS analysis we used 727,882 genotyped SNP with MAF>0.1%; for genetic correlation analysis using LDSC, we used 157,756 Genotyped SNPs mapped to HapMap3 SNPs; for computing heritability where mixed-effect association are performed using BOLT-LMM ^50, 51^ subsequent heritability estimation was performed using LDSC^2^, we used 1,201,838 imputed SNPs mapped to HapMap3 SNPs SNPs.

### Polygenic risk scores (PRS) analysis

Despite population stratification cannot be excluded^71^, to adjusted for and minimize the impact of population stratification, we applied mixed-effect association model to samples of British Isle ancestry group (N = 409,694) to compute PRS, for 10 heritable diseases that have the highest heritability z-scores. We used a mixed model to estimate effect size implemented by BOLT-LMM and constructed genome-wide PRS ^50^. For four diseases with more than 20,485 case (essential hypertension, arthropathy, asthma, and hypercholesterolemia), we downsampled controls to make the total sample size half of that of British isle ancestry population (N = 204,847) for computation efficiency; for other diseases, we sampled 9 controls for each case to ensure case proportion at or above 10% as recommended by BOLT-LMM (type 2 diabetes, varicose veins of lower extremity, hypothyroidism, other peripheral nerve disorders, major depressive disorder, and GRED). We used PLINK to select genotyped SNPs with MAF > 0.1% as recommended in BOLT-LMM. For each disease, we used 5-fold cross validation to estimate effect sizes using BOLT-LMM and computed the PRS on the held-out testing set. We used continuous-valued topic weights to analyse association between disease subtypes and PRS. The predictive PRS are used to compute the excess PRS over different topic loadings, by a linear regression where PRS is the response variable and topic weights is the predictor.

We compute the relative risk for each percentile of PRS using the following formula:

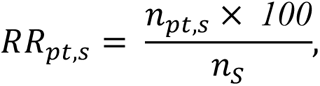

where *RR*_*pt*,*s*_is the relative risk of *s* subtype for the *pt*^*t*^*^ℎ^* PRS percentile (computed for the entire population); *n*_*pt*,*s*_is the number of cases in *s* subtype that has PRS within the *pt*^*t*^*^ℎ^* percentile; *n*_s_ is the number of cases in the *s* subtype.

### Genetic correlation analysis

We used discrete subtypes in genetic correlation analysis (different from the PRS analysis above). For each disease and disease subtype, we use a case-control matching strategy to construct data to estimate coefficients for genetic correlation analysis. For each case in the disease group, we pick four nearest neighbors (without replacement) from the control group, matching sex, BMI, year of birth and 40 genetic principal components. The covariates are available within the UK Biobank data set, over which we computed the principal components.

We then compute the Euclidean distance of the principal components to find the nearest neighbours in the population. All cases are matched with four controls except for 401.1 essential hypertension which has a sample size larger than 20% of the population. We match only one control for each hypertension case.

We perform logistic regression with sex and top 10 principal components as covariates to estimate the main variant effect of the 805,426 variants that are genotyped. We used PLINK 1.9 for association analysis^72^. With the summary statistics from the association analysis, we use LDSC to map the summary statistics to HapMap3 SNPs and match the effect and non-effect alleles^2,73^. Since UK Biobank is mostly of British Isle ancestry, we use the pre-computed LD score from the LDSC website. We estimated the heritability for each disease or disease subtype which has more than 1000 incidences (378 = 30 diseases subtypes + 348 diseases). We use 1000 incidence threshold as LDSC are more accurate with larger sample size. We focus on 71 disease and 18 disease subtypes of the 378 diseases subtypes and diseases that have heritability z-score above 4 for genetic correlation analysis.

The genetic correlation is computed for each pair of disease-disease, disease-subtype, and subtype-subtype using the logistic regression summary statistics and LD score regression. We report the estimate of genetic correlation and z-scores. Additionally, for pairs that involve subtypes (disease-subtype or subtype-subtype), we compute the excess genetic correlation, defined as the difference between the genetic correlation involving subtypes (disease-subtype and subtype-subtype) and the genetic correlation involving all disease diagnoses (disease-disease). For example, the genetic correlation between T2D-CER and hypertension-CVD is compared to the genetic correlation between all T2D and all hypertension. The z-score and p-value of the genetic correlation differences are reported. We note that genetic correlations between subtypes of the same disease are compared to 1. We only reported p-values of excess genetic correlation when both genetic correlation estimation has standard error <0.1 and at least one of the genetic correlation has |z-score|>4.

To avoid potential collider effects where subtypes are defined by topic components that are independent of the diseases, we performed the same genetic correlation analyses but match cases in each subtype with controls with similar topic loadings. We computed PCs from 23 variables (10 topic loadings, 10 PCs, year of birth, sex, and BMI) and used the nearest neighbour procedure (by Euclidean Distance) to find controls for each case. Here controls are chosen from individuals without the targeting disease, i.e. an individual with one subtype of the target disease could not be a control for the other subtypes. We performed the same analysis using this case-control matching procedure and compared the genetic correlation with the case-control procedure described above. We perform the analysis for four diseases that have evidence for genetic subtypes: asthma, type 2 diabetes, hypercholesterolemia, and hypertension. For one subtype (hypertension-CVD), the heritability (0.0313, s.e. = 0.0289) is below threshold after matching the topic, which was excluded in genetic correlation analysis.

### *F*_ST_ analysis

We used discrete subtypes in genetic correlation analysis (same as genetic correlation analysis above; different from the PRS analysis). To evaluate the genetic heterogeneity between disease subtypes, we estimated the *F*_ST_ for 52 diseases that have at least 500 incidences assigned to a secondary topic. To test the statistical significance of Fst, we adopted a permutation strategy and sampled the same number of controls of similar topic weights distribution for each subtype. The topic weights are matched by sampling (without replacement) the same number of controls for each dominant topic weight quartile of the cases (i.e. matching the topic that defines the subtype), which ensures the controls have the same topic weight stratification as the disease subtypes. We then compute the *F*_ST_ across the control groups matched for subtypes. We excluded three diseases, “hypertension”, “hypercholesterolemia”, and “arthropathy”, from *F*_ST_ analysis as we do not have enough controls that match topic weight distribution. The *F*_ST_s are computed using PLINK 1.9’s weighted mean across all genotyped SNPs, which report *F* statistics across all subtypes.

We obtained 1,000 permutation samples and reported the permutation p-value. Under the assumption that causal and non-causal variants have similar allele frequency differences across the subtypes, *F*_ST_ is a measure of causal genetic effect heterogeneity across subtypes.

### SNP x topic interaction test

We used continuous-valued topic weights in the SNP x topic interaction analysis (same as the PRS analysis; different from the genetic correlation and *F*_ST_ analyses). For the diseases that have heritability z-score above 4 in the UK Biobank, we further investigated whether there are interactions between genetic risk factors with the topic loadings. We used a fit a logistic regression model using following model:

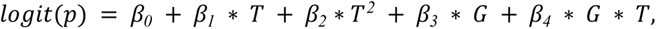

where *T* is individual topic weights for a specified topic, *G* is the genotype, and *p* is the probability of getting the disease. We computed the test statistics under the null that *β_4_* = *0*.

Since the simulation shows the interaction test is underpowered when the variant effects are small, we focus on the set of SNP that reaches genome-wide significance level to increase power to detect interaction effects. We performed LD-clumping using *r^2^* > *0*.*6* to remove variants that are in strong LD with the lead variants. We computed the test statistics using the model above (for testing *β_4_* = *0*) and computed study-wise FDR across 2530 disease-topic pairs.We used QQ plots to check that interaction test statistics computed using all non-subtype topics for each disease (which are expected to be null) were well-calibrated. (Supplementary Figure 31B).

To verify the significant interactions, we divided cases into quartiles based on topic loading for each disease-topic pair, and randomly sampled two controls that match the topic loading for each case. We estimated the main effect sizes for all GWAS-SNP within each quartile of topic loadings to capture effects that are modulated by topic weights. We focus on the SNPs that have significant interaction test statistics computed in the previous step and compare it with background SNPs that have genome-wide significant main effects but no interaction effect (P>0.05).

### Simulations of SNP x topic interaction

We simulate comorbidity with genetics to test interaction between genetic and comorbidity topics. We simulated 100 independent variants with MAF randomly sampled from the MAF of 888 independent disease associated SNPs. We assumed an additive model and simulated genotypes for the population using Hardy-Weinberg equilibrium. We simulated three types of genetic effects on topic and diseases on topic of the simulation framework described in Simulations of ATM method section:

● Genetics-topic effect: each variant is simulated to have an linear effect of 0.04 on the topic loading. We choose this value as after normalising the topic, a regression of causal variant to topic would have an effect size approximately 0.01 which is similar to our observation in the UK Biobank. The number of variants that are causal to the topic varies between 2 to 20. We simulated the effect on one topic by adding additive SNP effects and normalise the topic loadings of each patient. The topic-disease causality is a natural consequence following the generative process of sampling data.
● Genetic-disease-topic effect: we simulated a heritable disease that is causal to the topic. The disease is simulated with 20 causal variants each of effect size 0.15. We vary the disease-to-topic causal effect from 0.05 to 0.5, with a default value of 0.1 in other analyses (similar to the correlation we found in UK Biobank analysis). We simulated the effect on one topic by adding additive causal disease effects and normalise the topic loadings of each patient.
● The genetic effect could interact with the topic when contributing to disease risk. We simulated four additional diseases to represent different structures (Supplementary Figure 28).

○ Genetic effects interact with topic loading on altering disease risk. The interaction term is added to the mean of disease liability, which is sampled from a Gaussian distribution. The disease is then sampled by a threshold on the liability, where the incidence rate is by default 0.5. The interaction effect is varied from 0.4 to 4, with default value equal to 2.
○ Pleiotropy effects are simulated with a variant that have both genetic-disease and genetic-topic-disease effects. Both genetic and topic effects are added to the mean of disease liability. A disease is sampled by a threshold with default incidence rate equal to 0.5. The topic-disease effect is varied from 0.4 to 4, with default value equal to 2.
○ Pleiotropy effect with nonlinear topic-disease effect. A quadratic term of topic-disease effect added to the second model.
○ Pleiotropy effect with nonlinear genetic-disease effect. A quadratic term of genetic-disease effect added to the second model.

For disease-topic or topic-disease causal effects, we simulated 50 repetition at each causal effect size. For interaction analysis, we repeated 10 times at each parameter value, as there are more SNPs for uncertainty estimation. The simulated disease sets are fed into the inference procedure to infer the patient topic weights.

## Supplementary Tables (see Excel file)

**Supplementary Table 1. List of metrics for evaluating ATM performance.** The name, purpose, and implementation details of each metric are listed for comparison. For more details of each metric, see Methods.

**Supplementary Table 2. Simulation results for ATM on identifying disease subtypes.** We show the area under the precision-recall curve (AUPRC) for ATM in simulated data with two subtypes that have 20/10/5 years of age at diagnosis differences. Results for LDA (fifth column) are also shown for comparison. Rows show results for varying proportions of samples that belong to the smaller subtype. The results correspond to Figure 2.

**Supplementary Table 3. Characteristics of disease topics inferred from the UK Biobank.** For each topic, we listed the top 10 representative diseases (by topic loadings), heritability estimates, average topic weights (across all individuals), average age (weighted across all disease diagnosis assigned to the topic), proportional of variance explained by BMI, sex, Townsend deprivation index, and birth year.

**Supplementary Table 4. Topic loadings of 10 inferred disease topics across 348 diseases in the UK Biobank.** For each disease we reported the topic loading across diagnoses before 60 years old and after 60 years old. The Phecode, number of incidences, ICD-10 code, disease name, and Phecode systems are also listed for each disease. The values in this table correspond to Figure 3.

**Supplementary Table 5. Topic loadings as functions of age for 10 inferred disease topics.** For each topic, we listed the topic loading of each disease from age 30 to 80 years old. At each age point, the topic loadings add to one across diseases for each topic. The values correspond to Figure 4A and Supplementary Figure 13.

**Supplementary Table 6. Topic loadings as functions of age for 13 inferred disease topics from the All of Us data.** For each topic, we listed the topic loading of each disease between age 20 to 85. At each age point, the topic loadings add to one across diseases for each topic.

**Supplementary Table 7. Correlation of topic loadings between each pair of All of Us and UK Biobank topics.** Numeric values for Figure 5B.

**Supplementary Table 8. Prevalences in All of Us and UK Biobank for the 233 diseases that are shared between the two data sets.**

**Supplementary Table 9. Correlations between topic assignments for pairs of All of Us disease and UK Biobank disease across 233 diseases that are shared between the two data sets.** Disease associations to topics are measured using average topic assignments (see Methods for definition) for both UK Biobank and All of Us. Average topic assignments in All of Us are mapped to UK Biobank using topic loading correlation; the correlation for each disease pair in the UK Biobank topic space (Methods).

**Supplementary Table 10. Number of diagnoses assigned to each subtypes for 52 diseases.** We listed the number of diagnoses assigned to each disease subtypes by the diagnosis-specific topic probability, for the 52 diseases that have at least two subtypes with >500 diagnosis.

**Supplementary Table 11. Average age at diagnosis for each subtypes of the 52 diseases.** We listed the age at diagnosis across all diagnoses within each disease subtype, for the 52 diseases that have at least two subtypes with >500 diagnosis.

**Supplementary Table 12. Excess PRS in cases for all topics across 10 diseases (selected by heritability z-score).** We report the estimated changes in s.d. of PRS per unit changes in the patient topic weight, which is estimated through regression across disease diagnoses. The PRS was estimated using BOLT-LMM and all the cases of British Isle Ancestry. Numbers correspond to Figure 6B and Supplementary Figure 23.

**Supplementary Table 13. Excess genetic correlations.** Columns are: Phecode of the first disease (trait1), Phecode of the second trait (trait2), subtype of the first trait (topic1), subtype of the second trait (topic2), the z-score of genetic correlation between two disease subtypes (subtype.rho.zscore), estimate of genetic correlation between two disease subtypes (subtype.rho.est), standard error of genetic correlation between two disease subtypes (subtyp.rho.err), the z-score of genetic correlation between two diseases (all.rho.zscore), estimate of genetic correlation between two diseases (all.rho.est), standard error of genetic correlation between two diseases (all.rho.err), z-score of excess genetic correlation (diff.zscore), estimate of excess genetic correlation (diff.rg), absolute value of excess genetic correlation (diff.rg.zscore.abs), p-values for excess genetic correlation (P), FDR for excess genetic correlation (FDR), name of the first disease (phenotype.1), and name of the second disease (phenotype.2). We only reported p-values of excess genetic correlation when both genetic correlation estimation has standard error <0.1 and at least one of the genetic correlation has |z-score|>4. Numbers correspond to Figure 7 and Supplementary Figure 25.

**Supplementary Table 14. Heritability estimation for disease subtypes.** For each disease we list heritability estimates for two subtypes using LDSC. Topic.x refer to the subtype with the highest heritability, topic.y refer to subtype with the lowest heritability. We report the point estimate, standard error, z-scores of both disease subtypes. The z-score of heritability differences between the two subtypes are also reported. Note we used a different sample threshold 1000 (due to the power of LDSC), which includes 26 of the 52 diseases that have subtypes.

**Supplementary Table 15. Excess *F_ST_* estimation across disease subtypes.** We report the estimate of excess *F_ST_* (computed as the *F_ST_* across subtypes subtracted by the *F_ST_* from controls with matched topic weights). The p-values are for excess *F_ST_*>0, which is computed from 1000 randomly sampled control sets.

**Supplementary Table 16. GxTopic interaction tests across independent GWAS SNPs.** Each row represents one SNP x topic weight paris (disease subtype). OR, SE, STAT, and P represent the odds ratio, standard error, test statistics, and p-value of the main effects. SNPxTopic OR, SNPxTopic STAT, SNPxTopic P, SNPxTopic FDR represent the odds ratio, test statistics, p-value, and genome-wide FDR of testing the interaction effect in model 2 of Supplementary Figure 28. We use Studywise FDR which adjusts for multiple testing across GWAS SNPs of all disease subtypes.

**Supplementary Table 17. Significant SNP x topic interactions.** Same table as supplementary Table 16, but filtered to SNP-topic pairs with interaction effect passing FDR<0.1. Reported SNP-topic pairs were selected for topic weights specific effect estimation in Figure 8 and Supplementary Figure 30.

**Supplementary Table 18. Effect size estimation across topic weight quartiles for significant SNP x topic interactions.** Quartile, mean_effect, se_effect, refer to the quartile of topic weight, estimate of effect sizes of the SNP using case-controls from this quartile, and standard error of the effect size estimation. We also reported the nearest genes reported by GWAS Catalog. The last two columns report the P-value of effect size being different between the top and bottom quartiles and the FDR (across 2530 tests).

**Supplementary Table 19. Literature search of disease subtypes identified by ATM.** We searched on Pubmed using the description (ignoring conjunctions) AND “subtype” in title/abstract and manually screened the top 10 relevant results between 2012 and 2022. The studies that mentioned subtypes of the searched diseases are included in the “Published references” column. If there is no reference of target disease subtypes among the top 10 search results, we use “NA”. We note our search is not exhaustive but nevertheless provides information on whether subtypes of the target disease are described in studies involving target disease and subtypes.

**Supplementary Table 20. ATM running time.** We tested the running time on the UK Biobank data using ATM of varying topic number and parametric form of topic loadings. Degrees of freedom from 2 to 7 represent linear, quadratic polynomial, cubic polynomial, spline with one knot, spline with two knots, and spline with three knots. Note a few models with 50 topics did not converge.

## Supplementary Figures

**Supplementary Figure 1.**
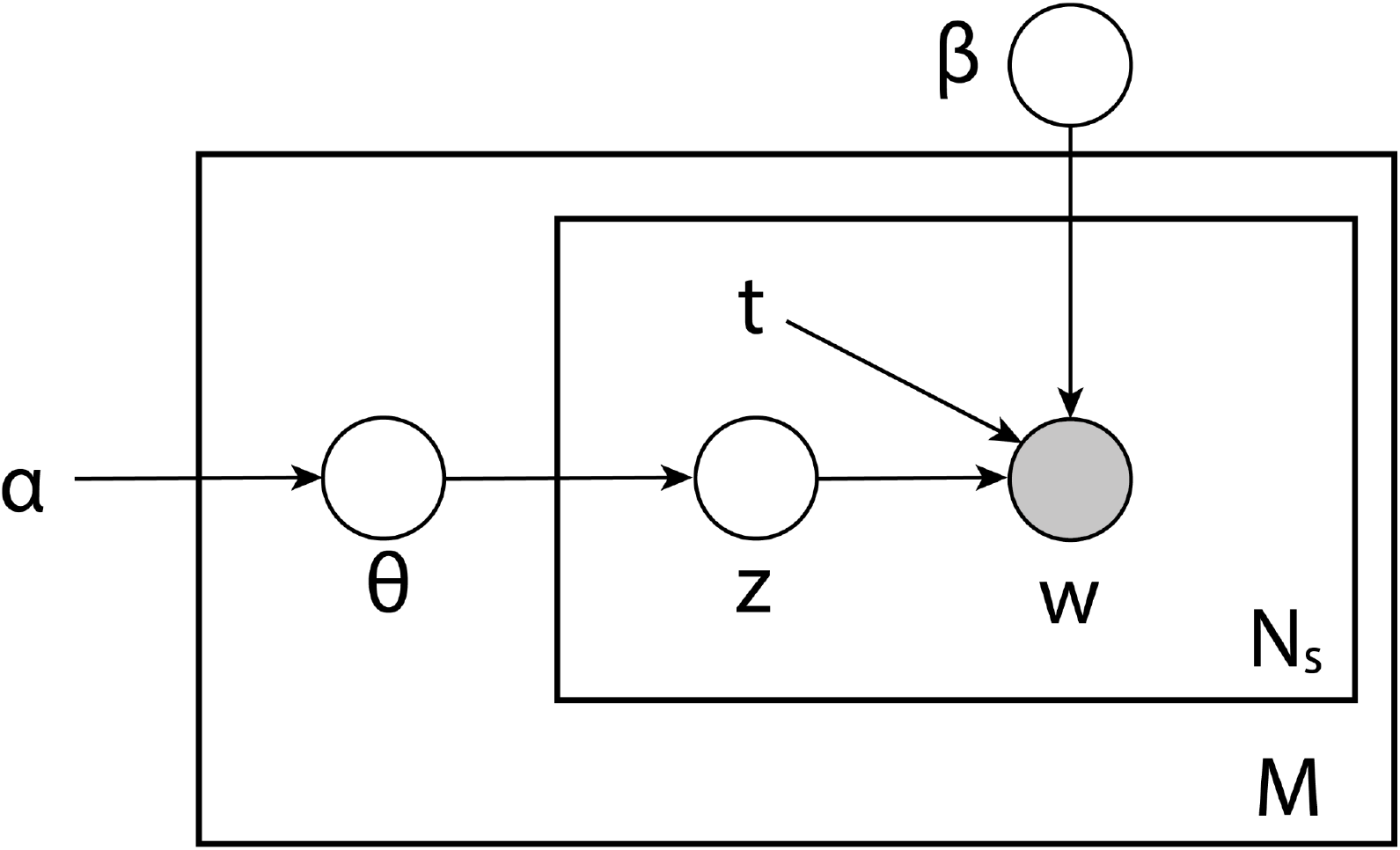
Plate notation of ATM generative model. *M* is the number of subjects; *N_s_* is the number of records within *s^th^* subject. All plates (circles) are variables in the generative process, where the plates with shade W is the observed variable and plates without shade are unobserved variables to be inferred; *θ* is the topic weight for all individuals; *z* is diagnosis-specific topic probability; *t* is the age at onset for each diagnosis; *β* is the topic loadings which are functions of age *t*; *α* is the (non-informative) hyperparameter of the prior distribution of *θ*. The generative process is described in the Methods and Supplementary Note.

**Supplementary Figure 2.**
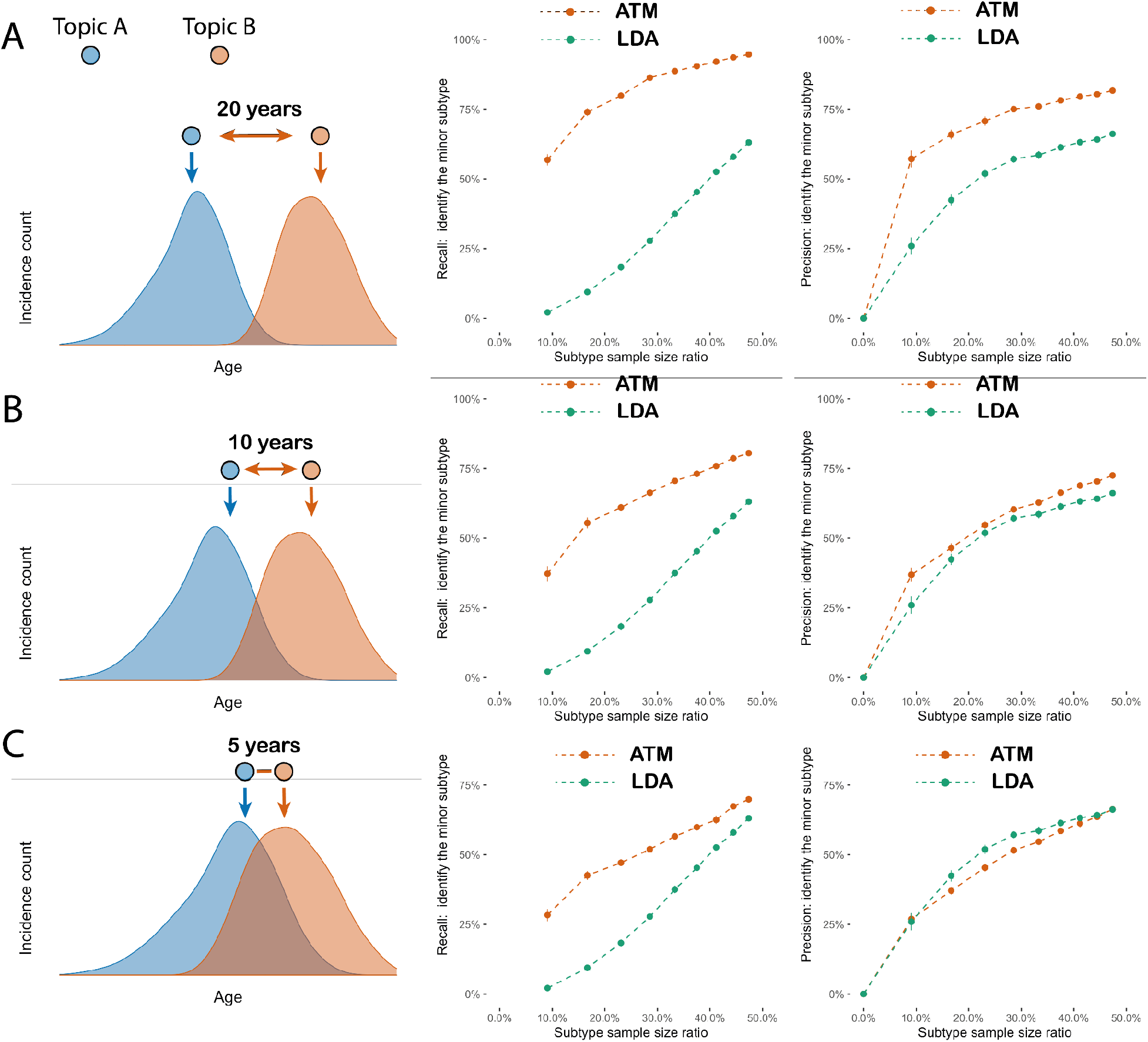
Additional simulation studies established the power of the method to identify comorbidity. The precision and recall rate to correctly assign incident disease to correct comorbidity profiles using Latent Dirichlet Allocation (LDA) and our method (ATM). X-axis refers to the proportion of cases that belong to the small subgroup; precision and recall are computed for the label incidences in the small subgroup. Each dot represents the mean of 100 simulations of 10,000 people, the bar shows the 95% confidence intervals. Red refers to the ATM and green refers to the LDA model.(a) Scenario where two subtypes are simulated with 20 years of difference in age at diagnosis. (b) Scenario where two subtypes are simulated with 10 years of difference in age at diagnosis. (c) Scenario where two subtypes are simulated with 5 years of age difference.

**Supplementary Figure 3.**
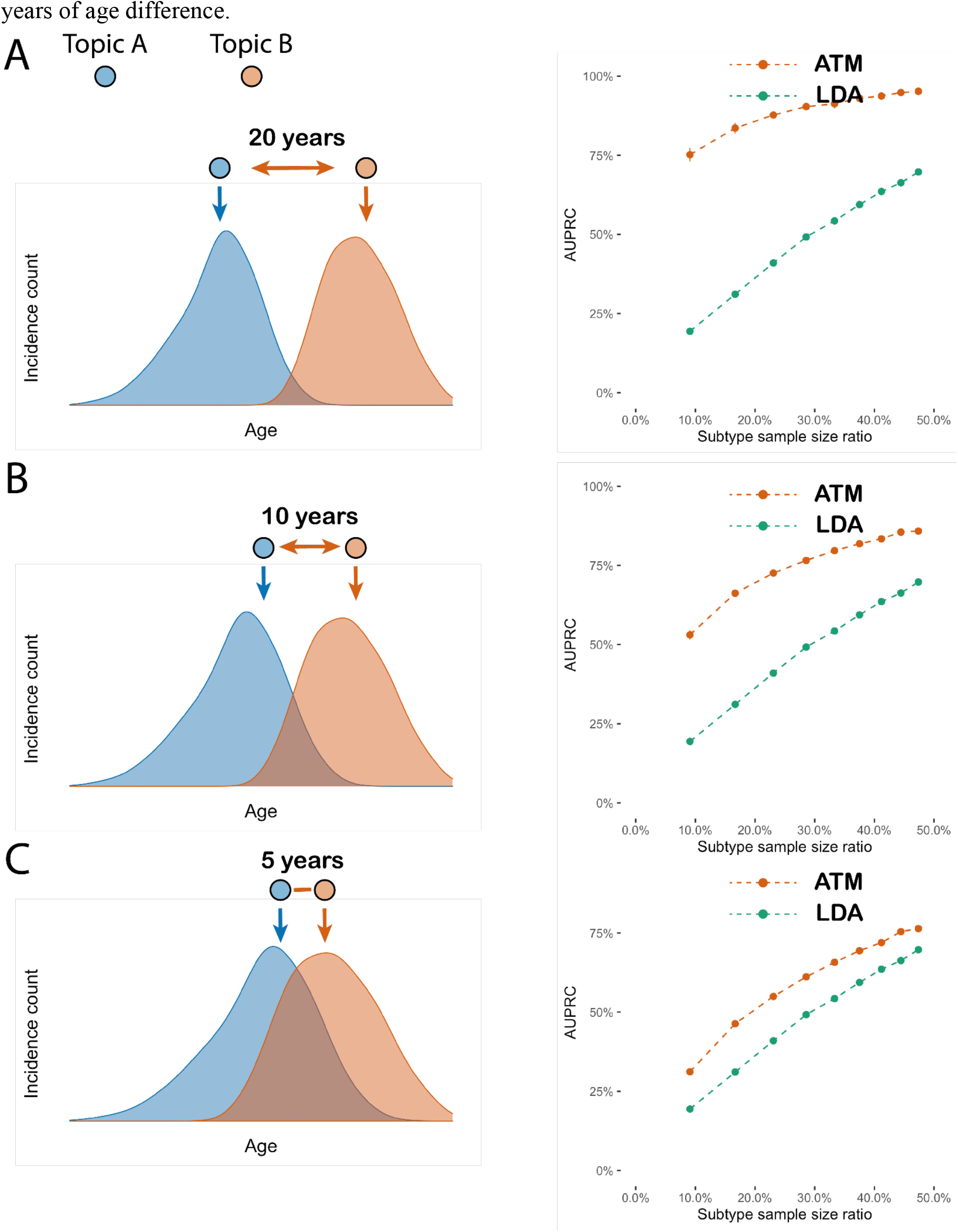
Same analysis as in Figure 2 but simulating the smaller subtype to have older age at diagnosis. The area under precision and recall curve (AUPRC) to correctly assign incident disease to correct comorbidity profiles using Latent Dirichlet Allocation (LDA) and ATM. X-axis refers to the proportion of cases that belong to the older subtype (the orange subtype); precision and recall are computed for classifying the incidences in the older subgroup. Each dot represents the mean of 100 simulations of 10,000 people, the bar shows the 95% confidence intervals. In the right column red refers to the ATM and green refers to the LDA model. Note AUPRC is only meaningful when precision and recall pertains to classifying the smaller subtype, therefore we simulate with the smaller subtype taking up to 50% of cases. (a) Scenario where two subtypes are simulated with 20 years of difference in age at diagnosis. (b) Scenario where two subtypes are simulated with 10 years of difference in age at diagnosis. (c) Scenario where two subtypes are simulated with 5 years of age difference.

**Supplementary Figure 4.**
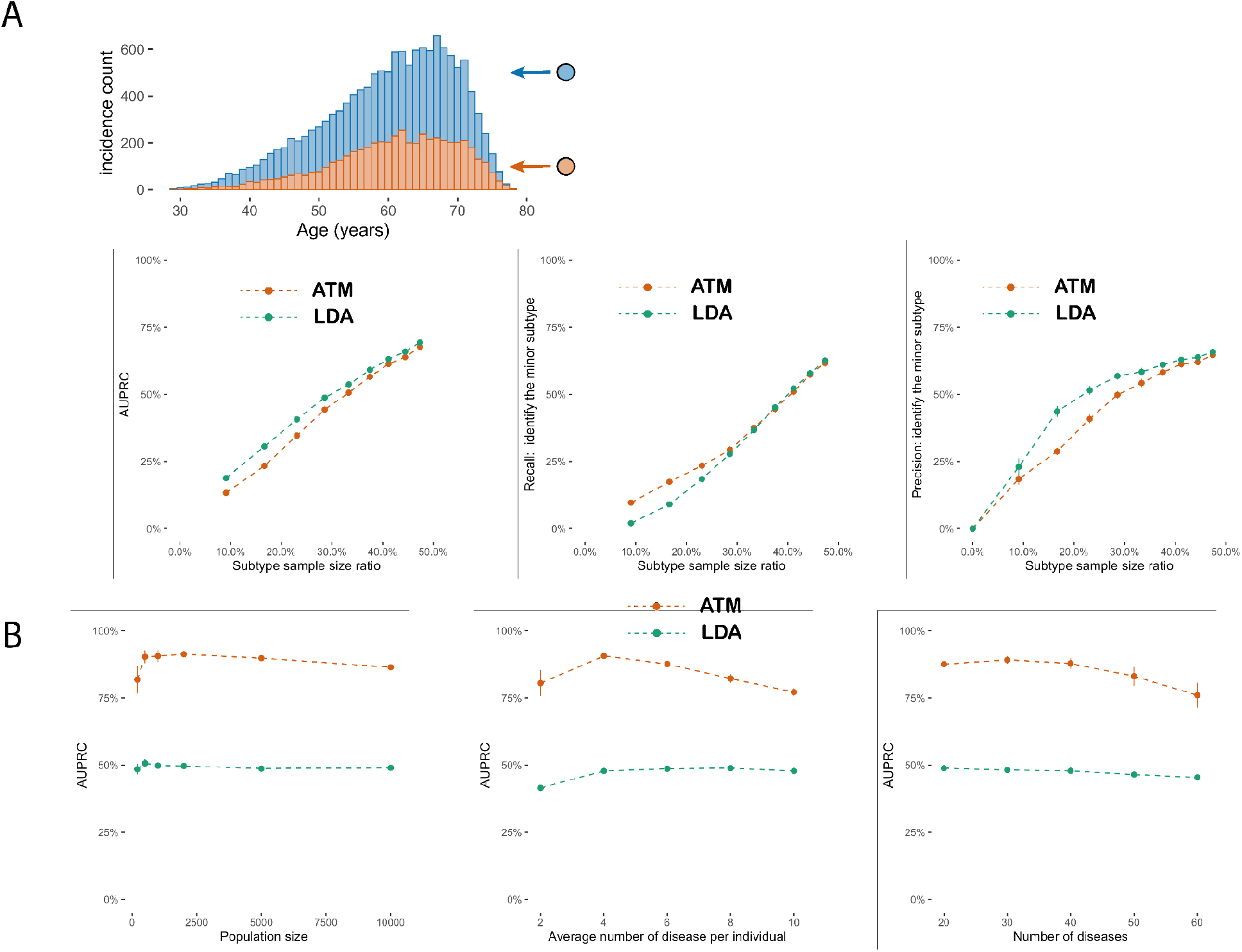
Additional simulation studies established the power of the method to identify comorbidity. (a) Same analysis as Figure 2 but simulated subtypes with same age at diagnosis distribution. LDA outperforms ATM slightly as we have additional regularisation when modelling topic loading as functions of age, while for LDA age is not modelled. (b) AUPRC computed as in Figure 2A with varying population size, average number of diseases per individual, and number of distinct diseases. Each dot shows the mean of 20 simulations and the bar shows 95% confidence interval.

**Supplementary Figure 5.**
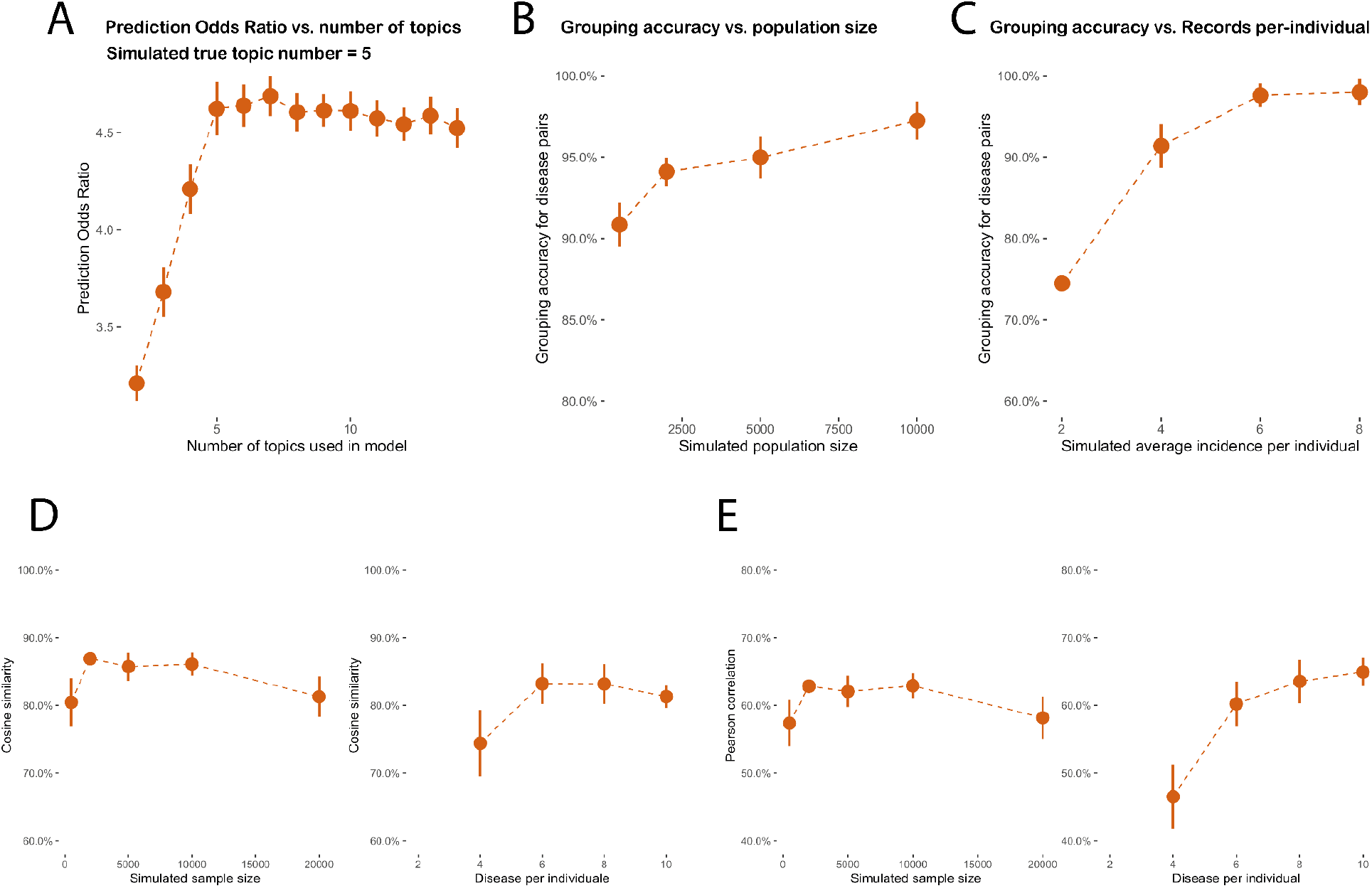
Simulations confirming that ATM could accurately recover topic loadings and topic weights. (a) We simulated data using 5 topics while fitting models of varying topic numbers. To compute prediction odds ratios (see Methods), we used 80% of data as training data to fit ATM and computed prediction odds ratio in the held out data, where we use the topic loading computed from the training data and prior diseases to infer the topic weights to predict the target diseases. The simulation was performed for 20 replications for each topic number in the inference. (b-c) We assign each disease to a single topic based on topic loading and compute the grouping accuracy as the proportion of disease pairs that are correctly grouped to the same topic. The grouping accuracy remains high for varying simulated population size and average disease per individual. (d) Recovery of topic loadings. We evaluate the accuracy of topic loading inference by computing the cosine similarity between inferred topic loading with the underlying truth. We match the inferred topics with the true topics using correlation of topic weights, using a greedy procedure (matching the first inferred topic from all true topics and then matching the next topic from the remaining not-matched true topics) to ensure the matching is bijectively. (e) Recovery of topic weights. We evaluate the accuracy of topic weight inference by computing the correlation of inferred topic weights and with the underlying truth. The ordering of topics uses the same strategy as in panel d.

**Supplementary Figure 6.**
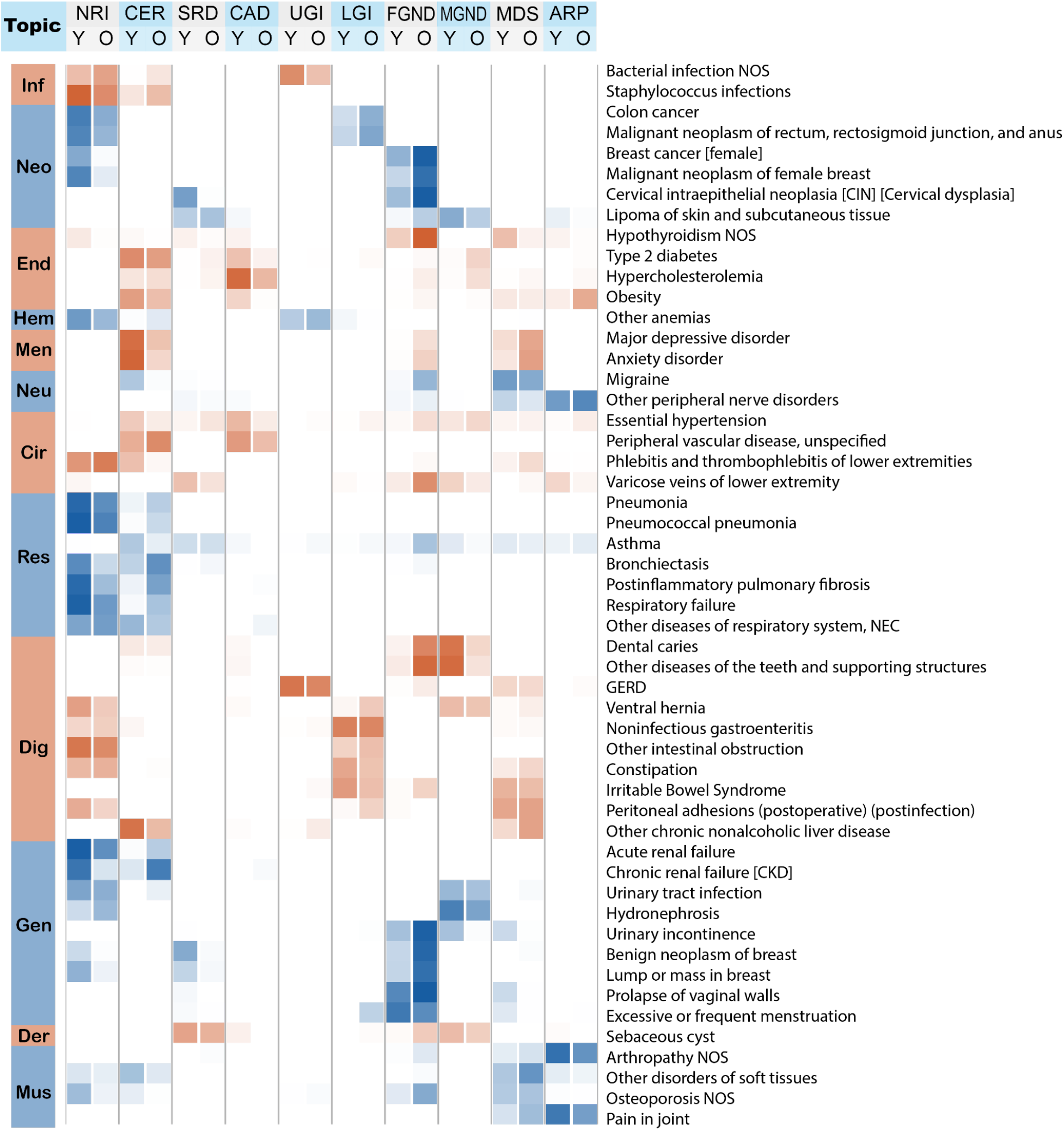
Posterior topic distributions are different between age groups for diseases that have subtypes. The figure has the same legends as Figure 3A but focusing on 52 diseases that have a subtype with at least 500 incidences.

**Supplementary Figure 7.**
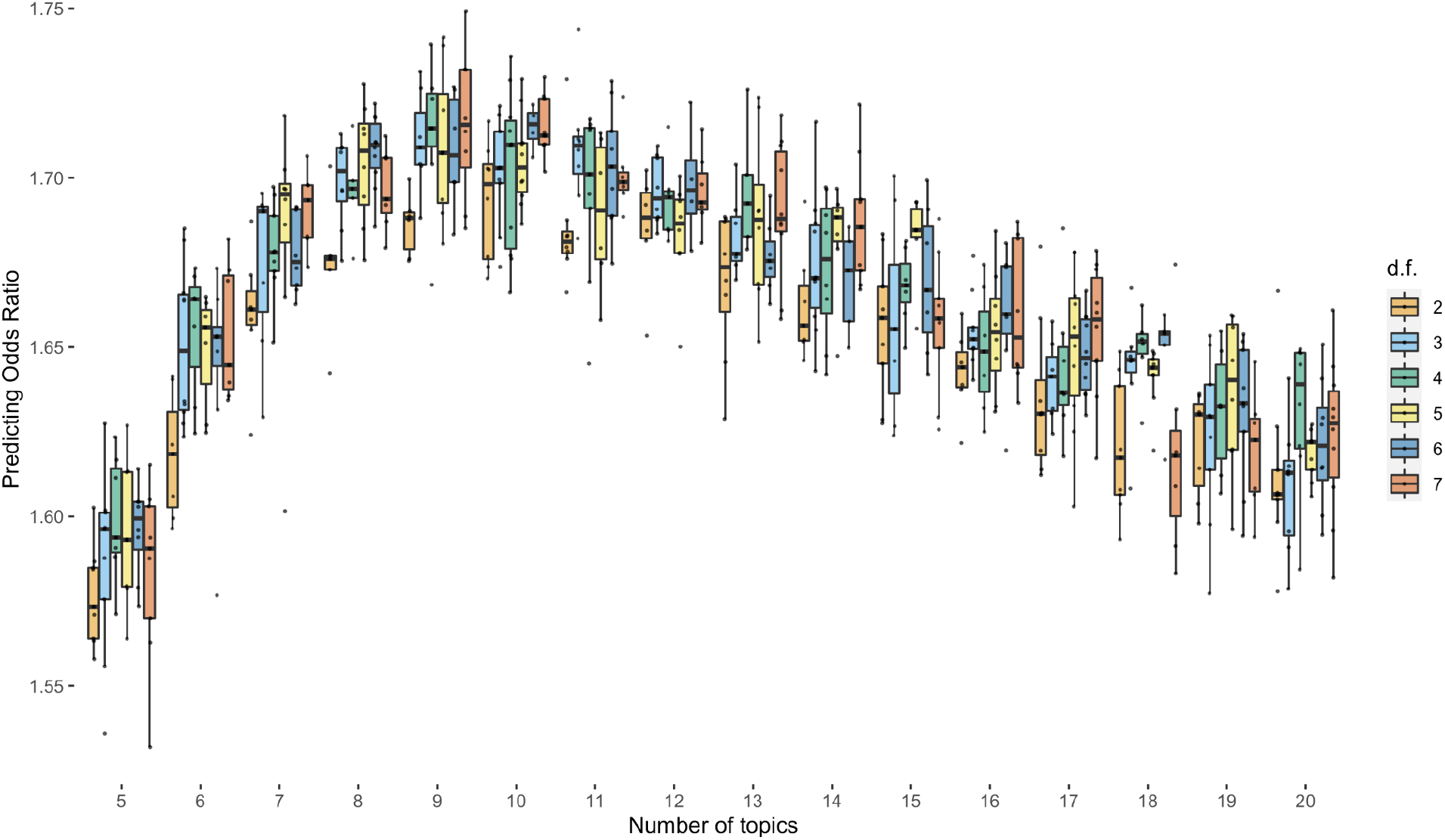
Prediction odds ratio across different model configurations. Each dot represents one inference on a random training and testing split of the UK Biobank individuals. The models are run with different topic numbers and parametric configurations of topic loadings. Degrees of freedom (d.f.) from 2 to 7 represent linear, quadratic polynomial, cubic polynomial, spline with one knot, spline with two knots, and spline with three knots.The prediction odds ratios are computed on the testing data using topic loadings inferred from the training data and topic weights inferred using previous diseases of testing individuals. The odds ratios are between the odds that target diseases are within model-predicted top percentile disease set versus the odds that target diseases are within the prevalence-ordered top percentile disease set.

**Supplementary Figure 8.**
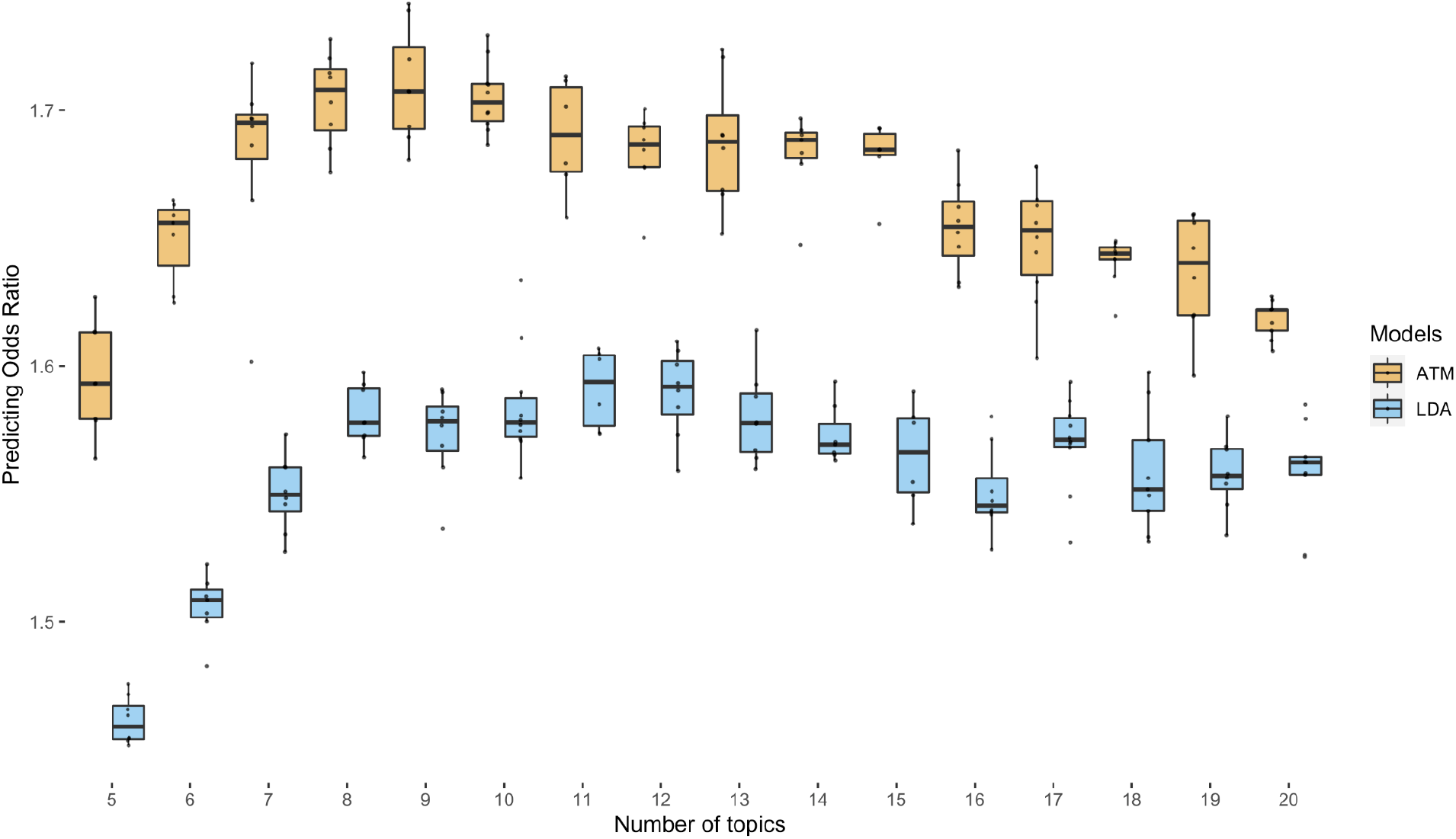
Comparison of prediction odds ratio between LDA and ATM. Each dot represents results from running either ATM or LDA on the same random training and testing split. The models were run with different topic numbers and we chose a cubic spline with one knot for configuring ATM topic loadings. The prediction odds ratios are computed on the testing data using topic loadings inferred from the training data and topic weights inferred using previous diseases of testing individuals. The odds ratios are between the odds that target diseases are within model-predicted top percentile disease set versus the odds that target diseases are within the prevalence-ordered top percentile disease set. For the optimal model with 10 topics, ATM has an average prediction odds ratio 1.71 (across 10 random training-testing splits); LDA has an average prediction odds ratio 1.58 (across 10 random training-testing splits).

**Supplementary Figure 9.**
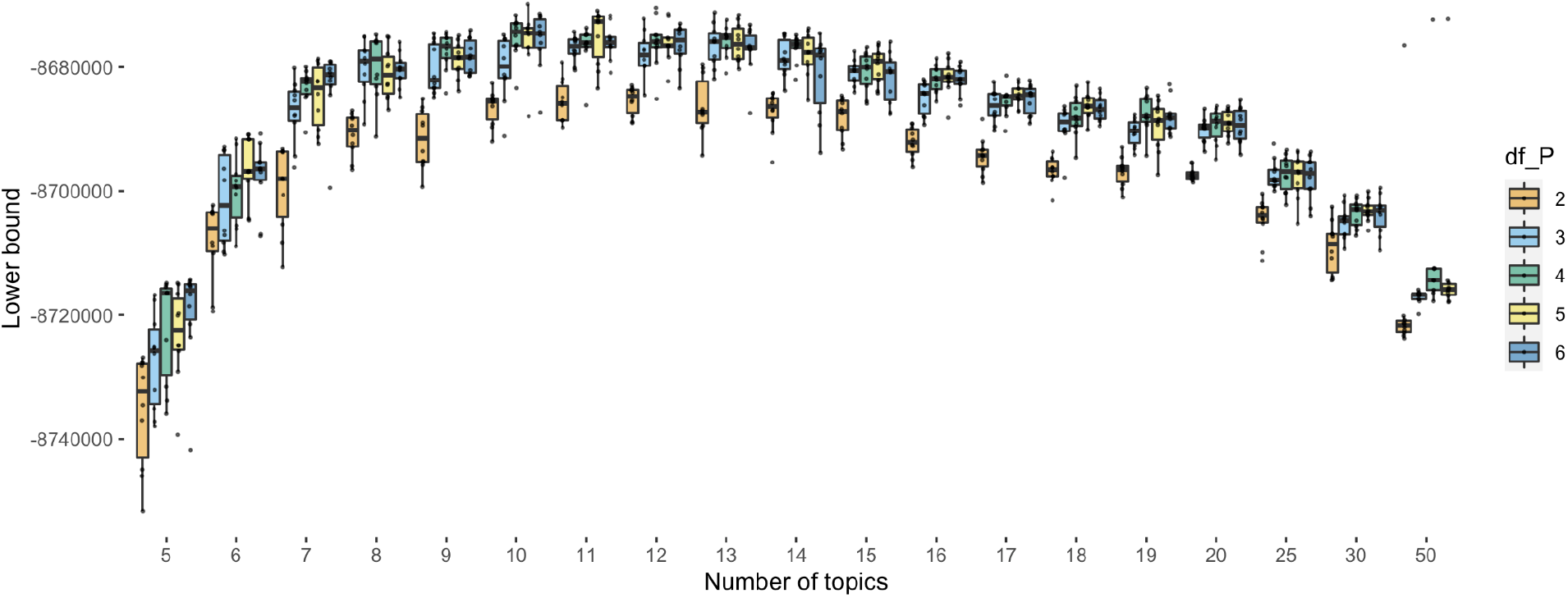
Evidence lower bound (ELBO) of different model configurations on the entire dataset. Each dot represents one inference on a random training and testing split of the UK Biobank individuals. The models are run with different topic numbers and parametric configurations of topic loadings. Degrees of freedom (d.f.) from 2 to 7 represent linear, quadratic polynomial, cubic polynomial, spline with one knot, spline with two knots, and spline with three knots.

**Supplementary Figure 10.**
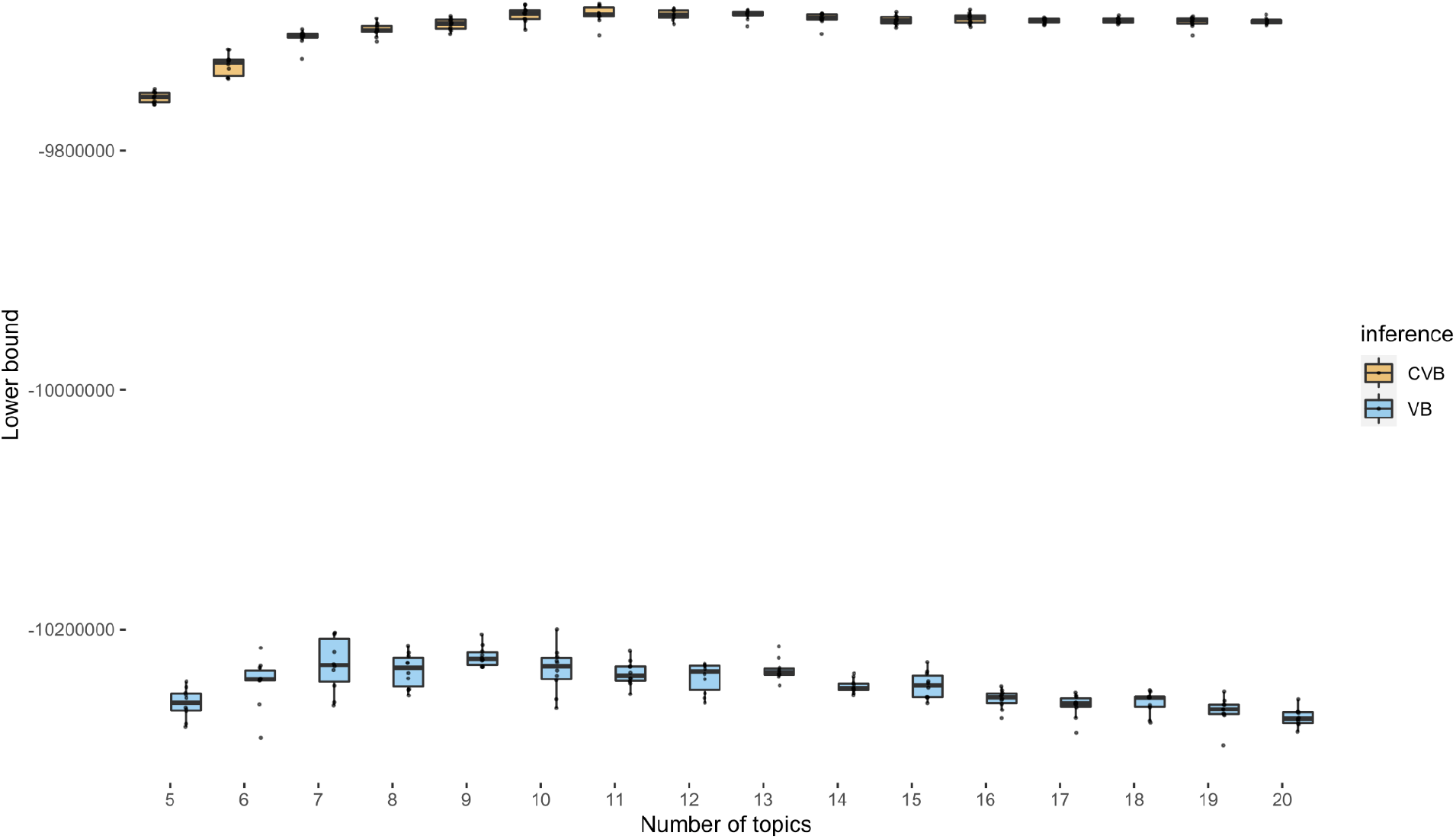
Comparison of ELBO for collapsed variational inference and mean-field variational inference. ELBO is computed by fitting the ATM using two inference methods on the entire UK Biobank dataset, where the topic loadings are configured as cubic polynomials. Models of different numbers of topics are fitted with 10 random initialisations for both CVB and the VB (mean-field variational inference, which is a more commonly used inference method for Bayesian models). The ELBO of an inference methods is a lower bound that approximates the evidence function, which depends on the number of topics and parametric form of topic loading, but not the inference methods; higher ELBO means better inference accuracy.

**Supplementary Figure 11.**
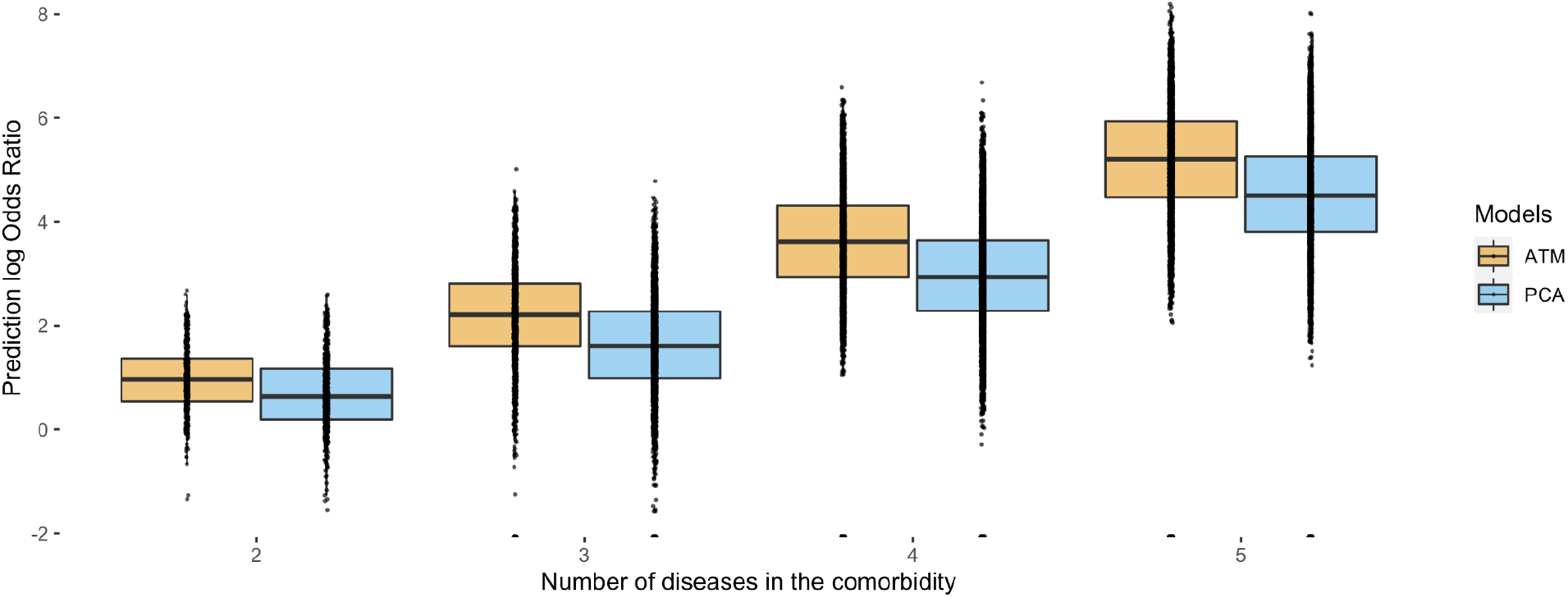
Prediction log odds ratio of comorbidities. Diseases combinations (comorbidities) are extracted from topics loadings that are trained on a training set, using max value of average topic assignments (Methods). Roughly, the odds ratio of each disease combination is computed by selecting all disease sets containing combinations of 2, 3, 4, and 5 diseases assigned to the same topic by max topic loading, and dividing incidences where the disease sets appeared in one patient by the expected number in an independent testing set. We show the comparison of ATM and PCA for all combinations of 2, 3, 4, and 5 diseases; here we use PCA as we wish to show the superiority of topic modelling in identifying clusters of disease compared to other low-rank methods that are not based on multinomial distribution.

**Supplementary Figure 12:**
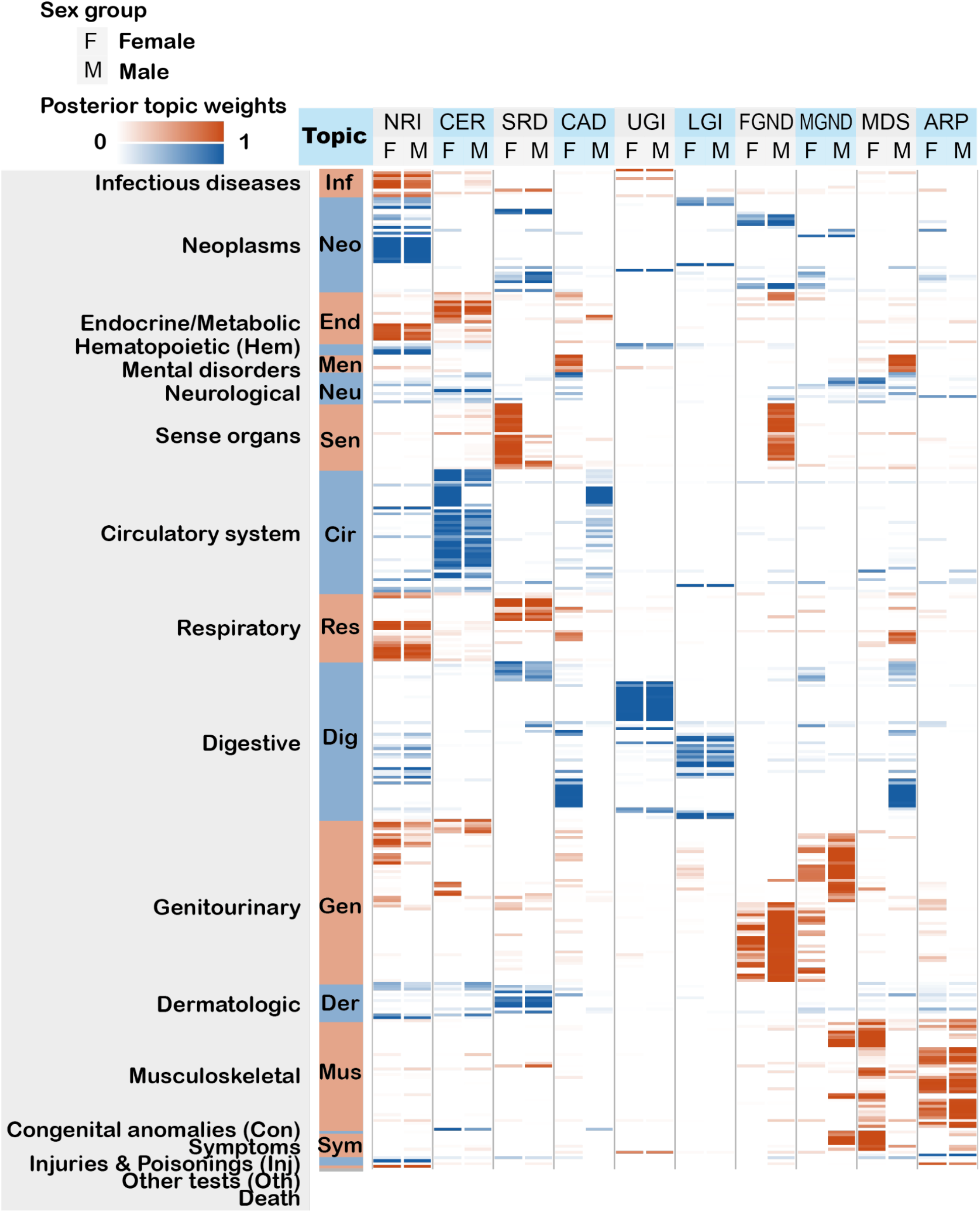
Posterior topic distributions of female and male populations. The figure is the same as Figure 3A but comparing the topics that are inferred from female and male populations separately.

**Supplementary Figure 13.**
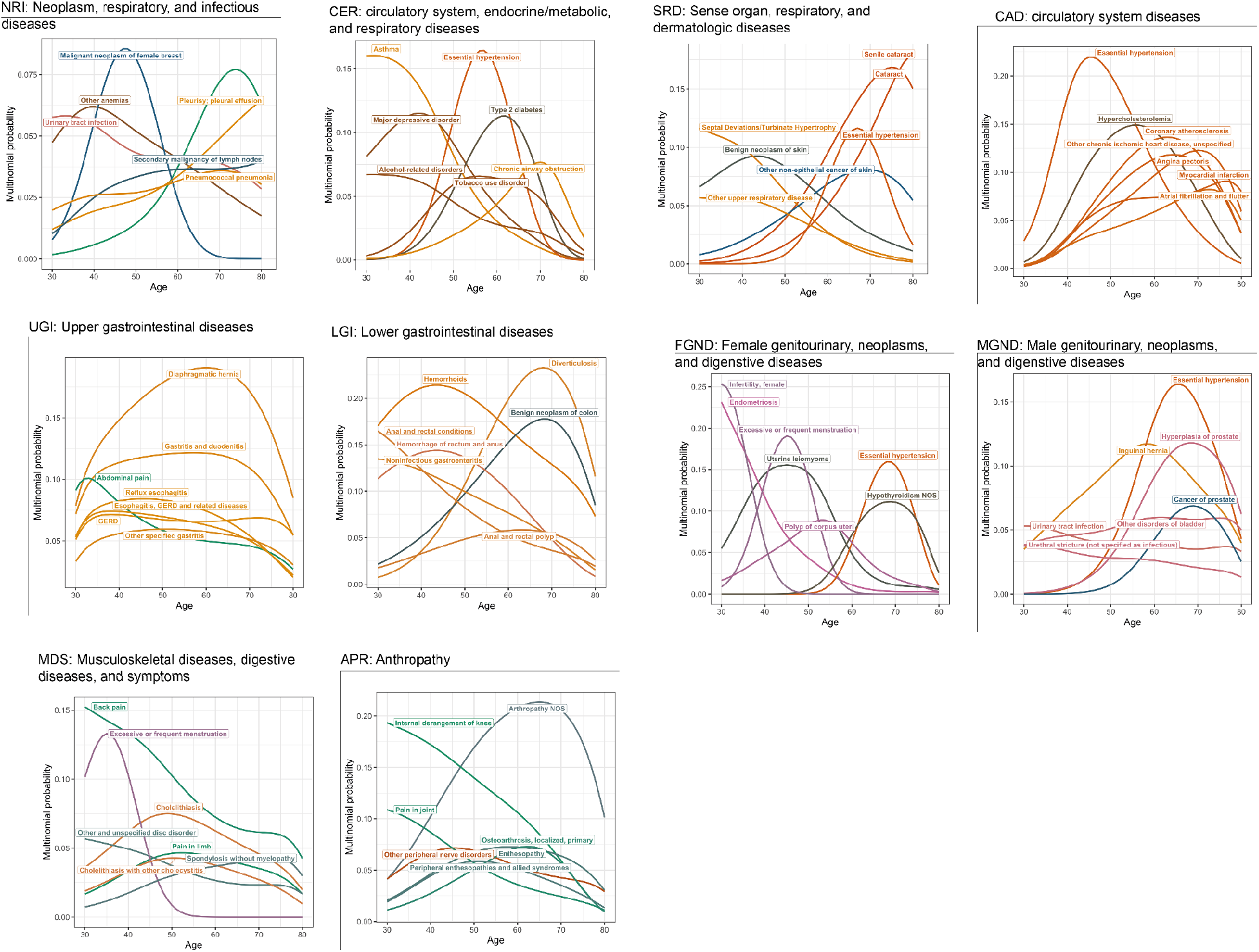
Top seven diseases in each comorbidity topic. Seven diseases that have highest loading within the topic are shown for each comorbidity topic. Colour of the curves reflect the ordering of Phecodes. We chose seven for best visual presentation. Numerical results are reported in Supplementary Table 5.

**Supplementary Figure 14.**
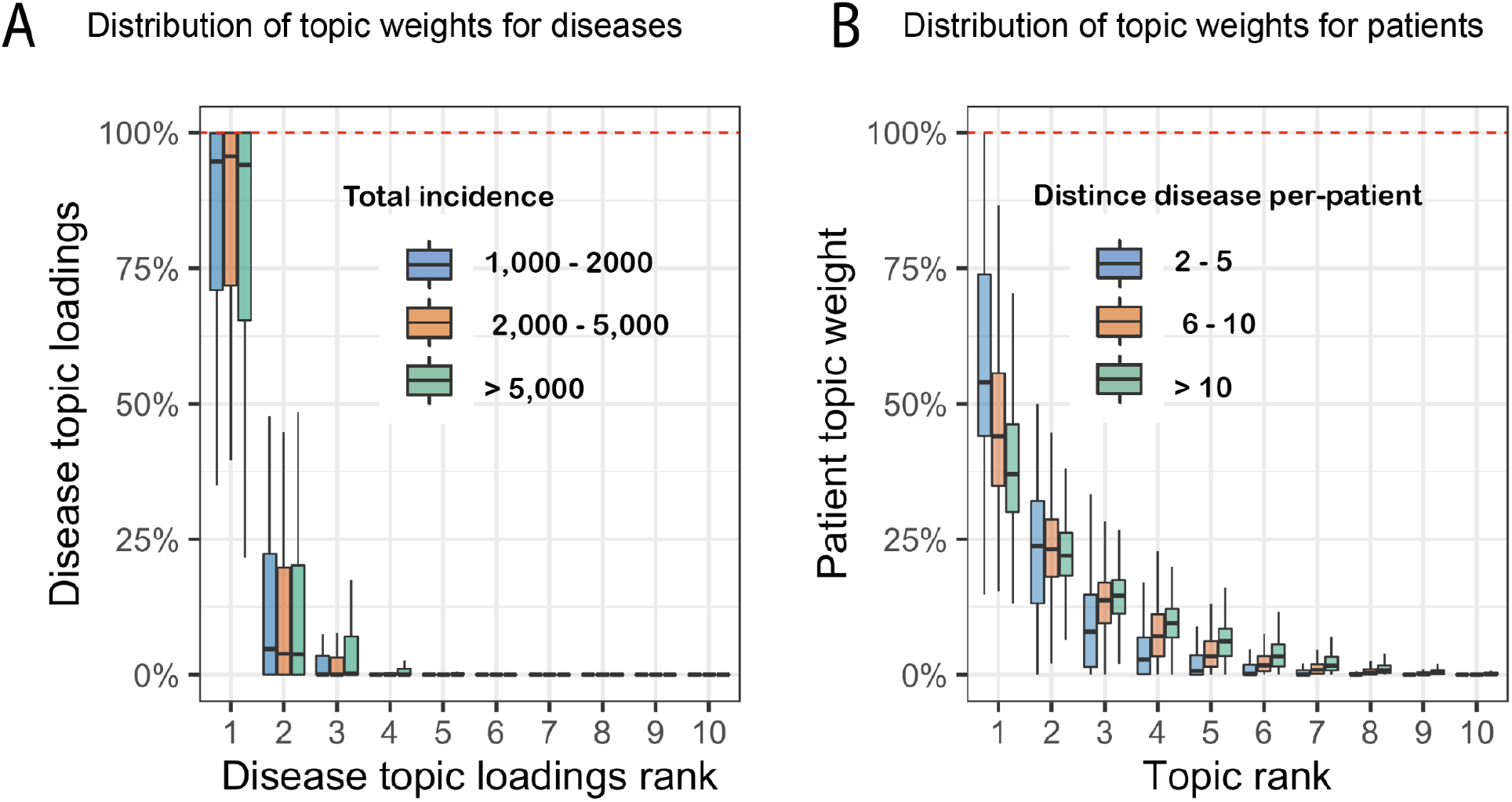
Additional topic sparsity analysis. (a) Sparsity of disease topic loadings. Box plot shows the distribution of topic loading for disease of different incidence numbers. (b) Sparsity of patient topic weights. Box plot shows the topic weight distribution in decreasing order for individuals with different numbers of diagnosis.

**Supplementary Figure 15.**
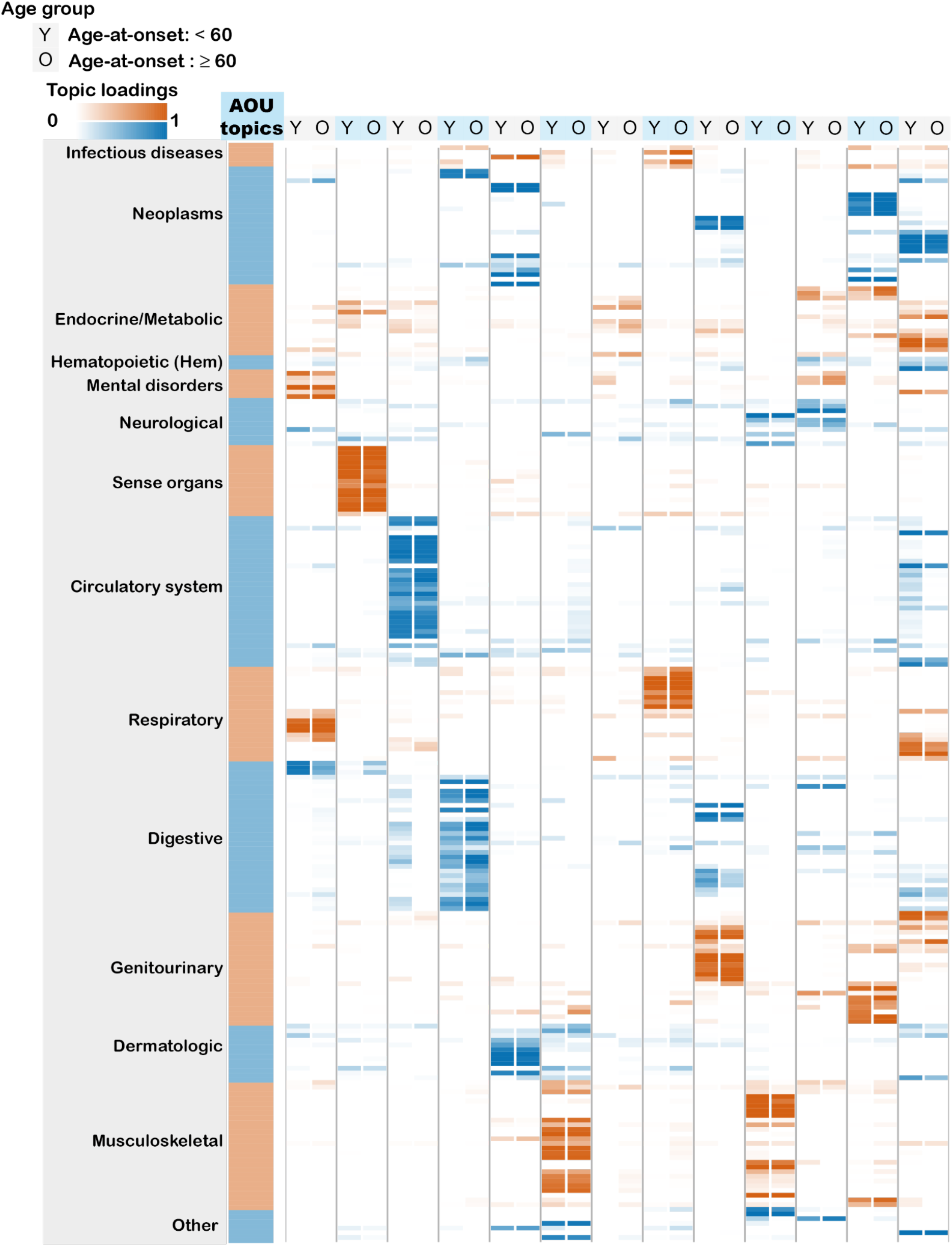
Age-dependent topic loadings of 13 inferred disease topics across 233 diseases in the All of Us. We report topic loadings averaged across younger ages (age at diagnosis < 60) and older ages (age at diagnosis > 60). Row labels denote disease categories ordered by Phecode systems, with alternating blue and red color for visualisation purposes; “Other” is a merge of five Phecode systems: “congenital anomalies”, “symptoms”, “injuries & poisoning”, “other tests”, and “death” (which is treated as an additional disease, see Methods). Topics are ordered by the corresponding Phecode system. This figure is an All of Us equivalent of Figure 3.

**Supplementary Figure 16.**
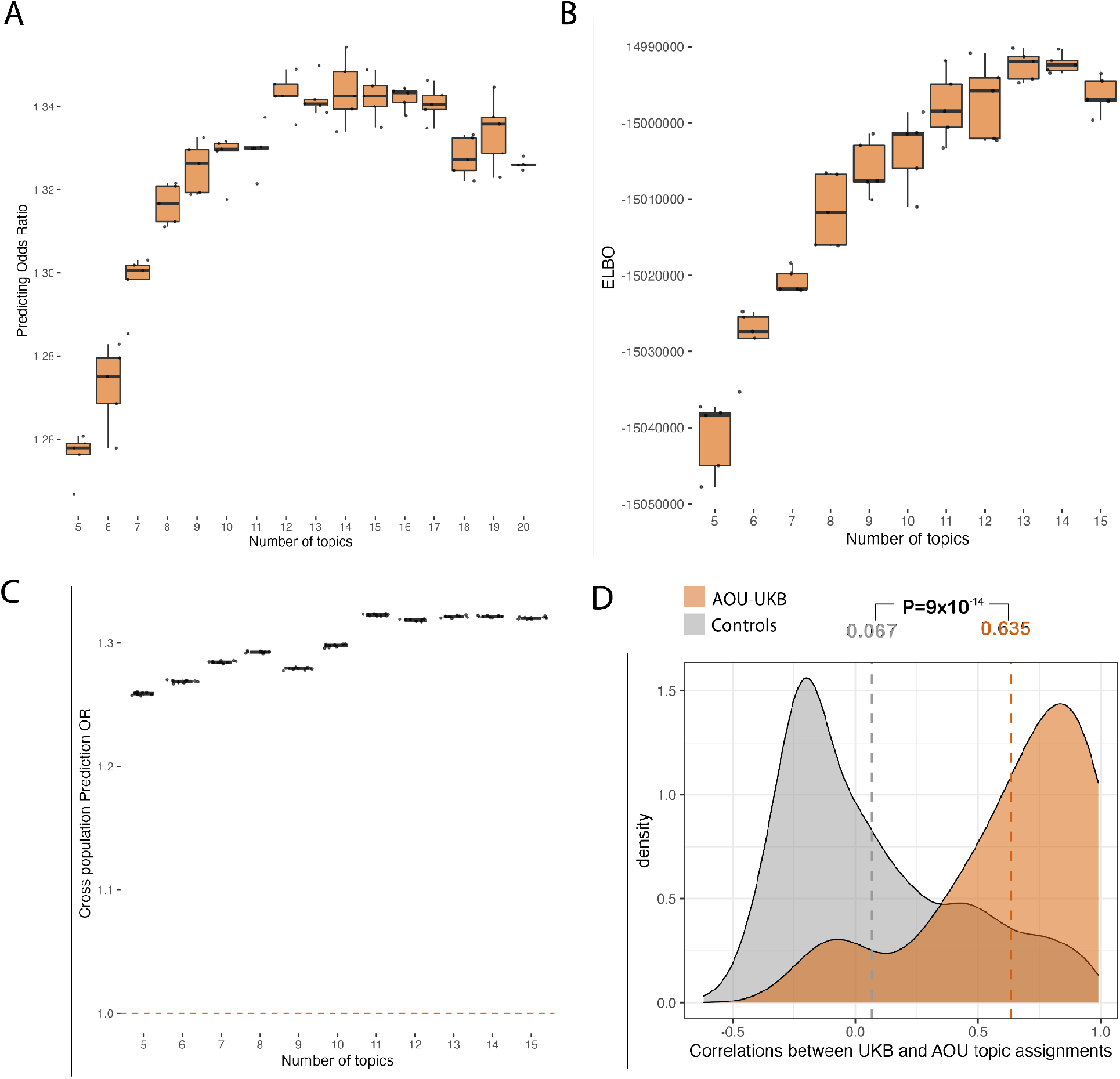
ATM infers disease topics from All of Us cohort which align with topics from UK Biobank. (A) Prediction odds ratio using ATM model with different topic numbers in All of Us. Each dot represents one of the five-fold cross validation within the All of Us individuals. (B) Evidence lower bound (ELBO) of different ATM model configurations on the entire All of Us dataset. Each dot represents one inference with random initialization. The models are run with different topic numbers and same configurations of topic loadings (spline model with one knot). (C) Prediction odds ratio on UK Biobank individuals using All of Us topic loadings. We divide the UKBiobank population into 10 jackknife blocks and each dot represents the prediction odds ratio on one leave-one-out jacknife sample. Topic weights are inferred using prior diseases of UKBB individuals, using loadings trained from All of Us. The odds ratios are between the odds that target diseases are within model-predicted top two-percentile disease set versus the odds that target diseases are within the prevalence-ordered top two-percentile disease set. Prediction odds ratio is 1.32 (s.e. = 0.0027) when using the optimal 13 All of Us topic to predict UK Biobank diagnoses. We chose the top-two percentile to match the UK Biobank analysis as All of Us has 233 of the 348 diseases analysed in the UK Biobank. (D) Correlations between UKB and AOU topic assignments for 41 diseases with subtypes between AOU and UKB (red shade) are significantly higher than expected (grey shade). The correlation between 41 AOU-UKB disease pairs are reported in Table 2 and Supplementary Figure 19. Grey shade is the distribution of non-diagonal correlations in Supplementary Figure 19. Grey and red vertical dashed line reports the mean of the grey and red shades; P-value is for the difference of the mean.

**Supplementary Figure 17.**
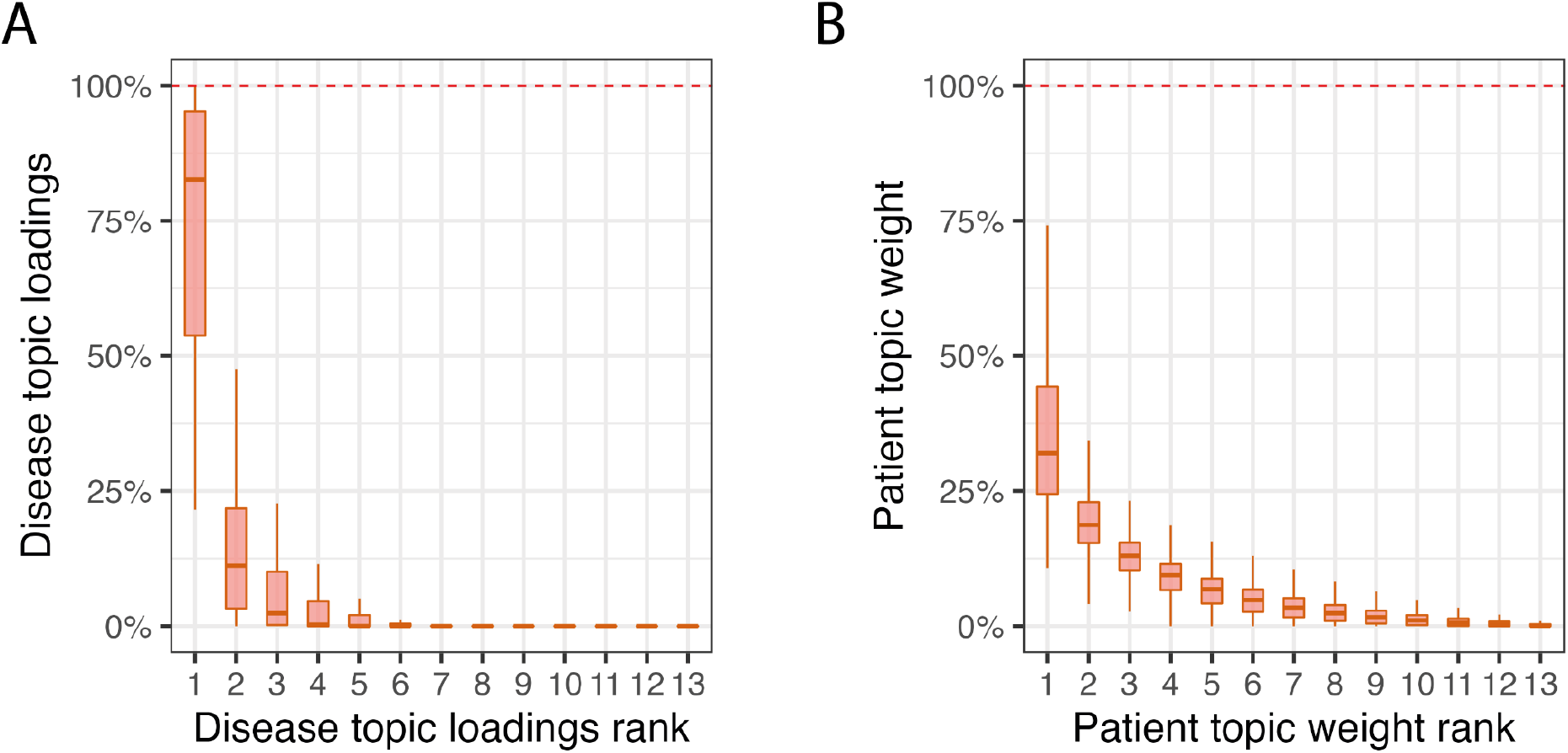
Distribution of topic loading across diseases and topic weights across patients for All of Us. (A) Box plot of disease topic loading as a function of rank; disease topic loadings are computed as a weighted average across all values of age at diagnosis. (A) Box plot of patient topic weight as a function of rank. This figure is an All of Us equivalent of Figure 4B-C.

**Supplementary Figure 18.**
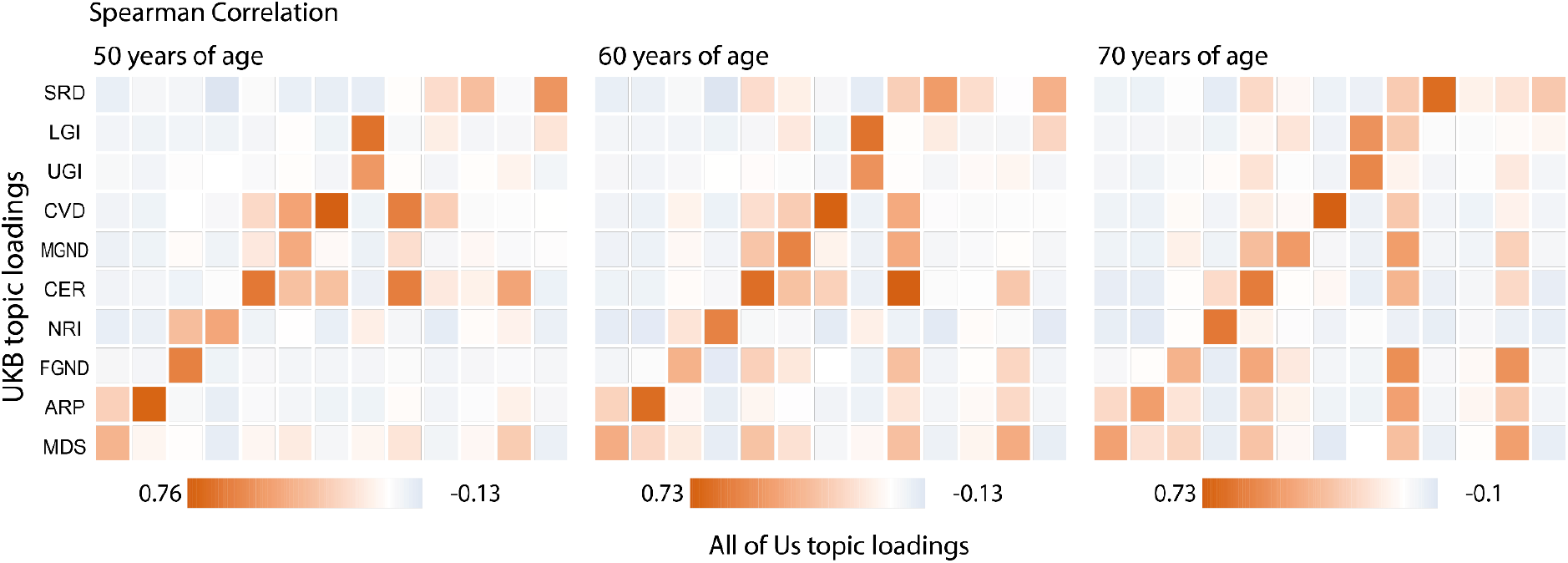
correlation between topic loadings from UK Bibank (y-axis) and All of Us (x-axis) for three age slices. The figures are the age-specific versions for Figure 5C.

**Supplementary Figure 19.**
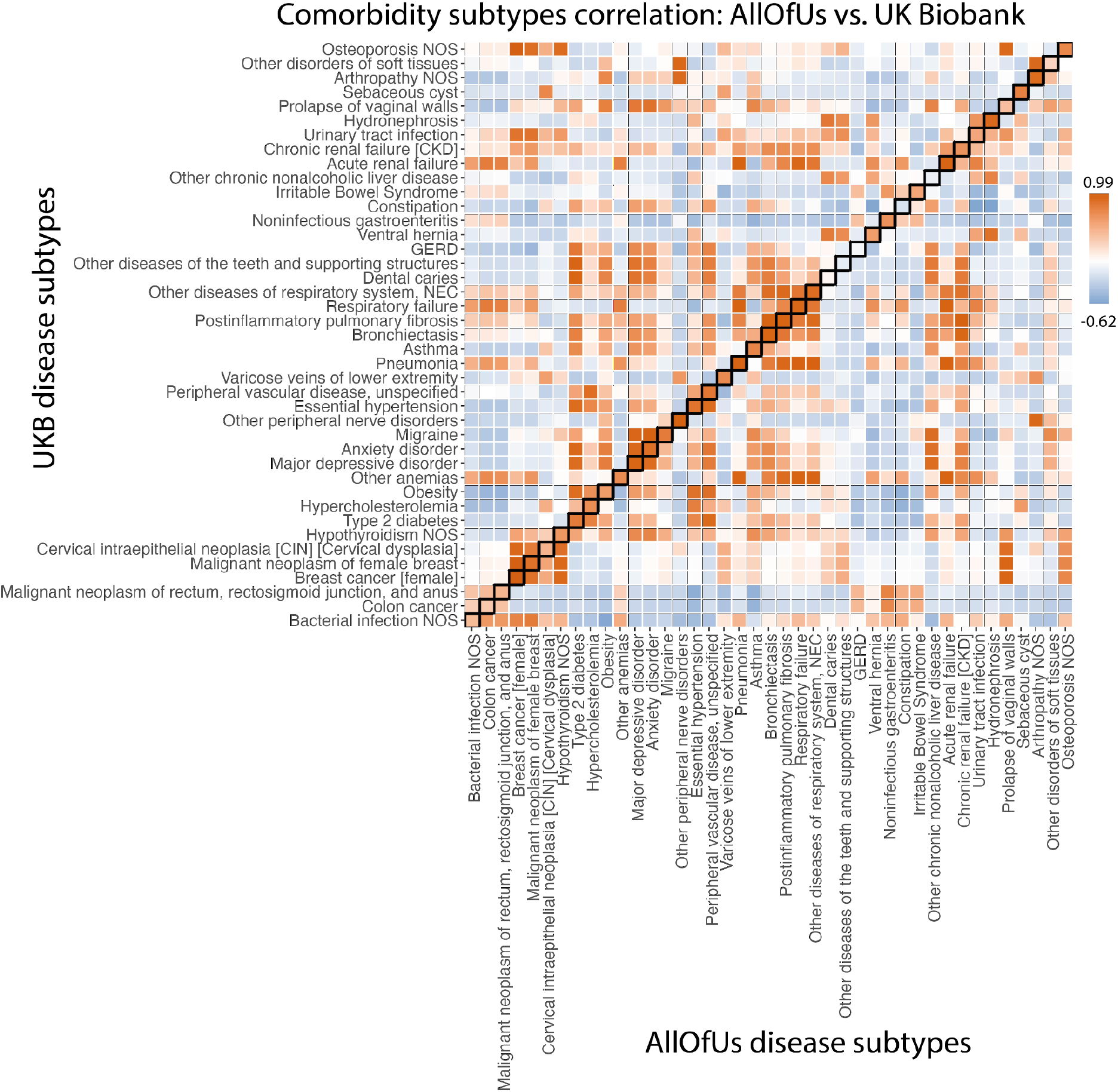
Subtype correlations between UK Biobank and All of Us for 41 diseases that are presented in both datasets and have subtypes in UK Biobank. Each box of the heatmap shows the correlation of average diagnosis-specific topic probability between a disease from All of Us and the other disease from UK Biobank. The diagnosis-specific topic probabilities from All of Us were mapped to UK Biobank based on proportional variance between the two topic spaces (Methods).

**Supplementary Figure 20.**
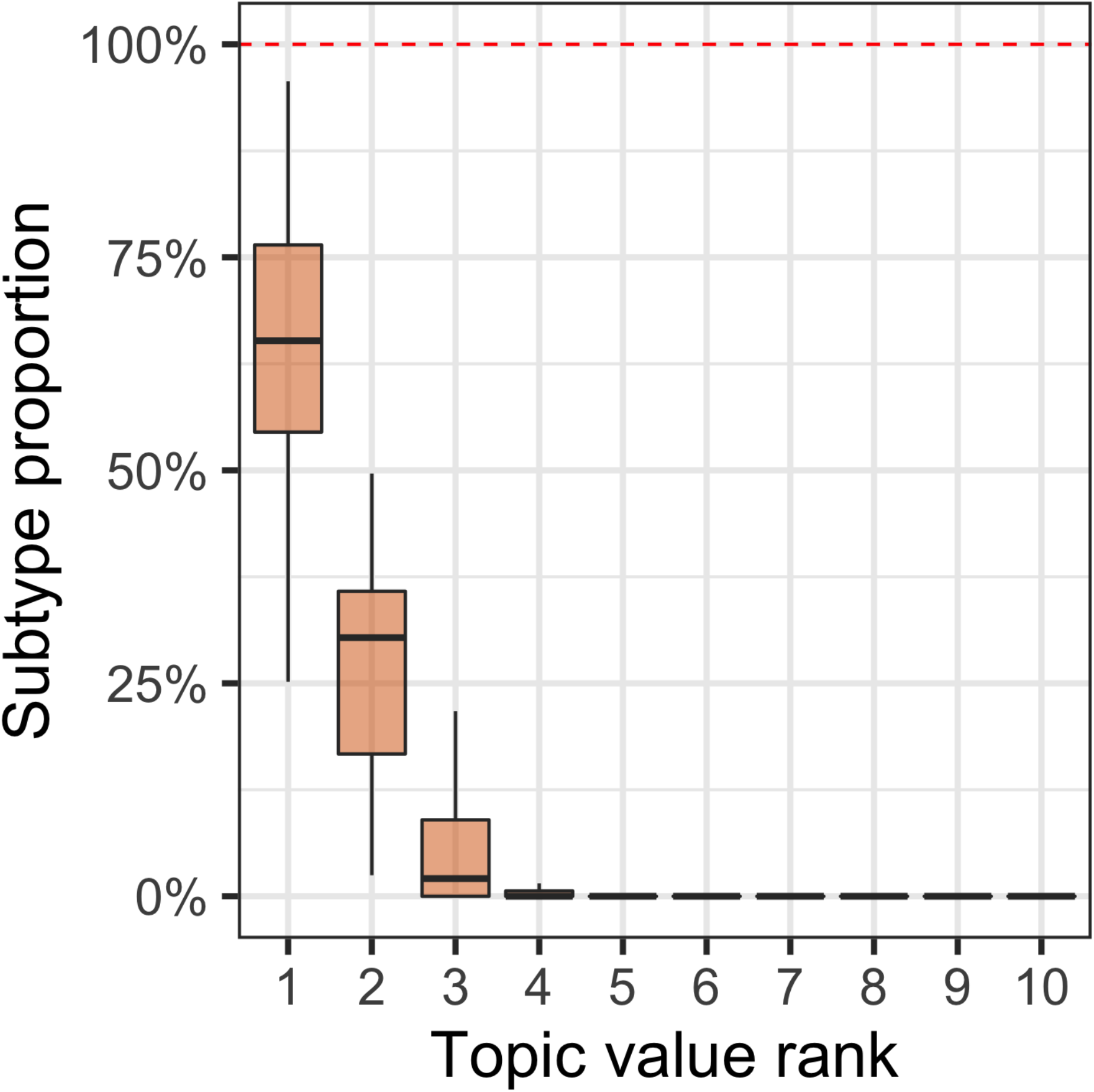
Topic distribution for the 52 diseases that have at least 500 cases assigned to distinct topics. For each disease of the 52 diseases, we computed the proportion of diagnoses assigned to each subtype. The box plot shows the distribution of the subtype proportion from the largest (leftmost boxes) to the smallest. For nearly all diseases, the cases are concentrated into three subtypes, with very few cases assigned to other topics.

**Supplementary Figure 21.**
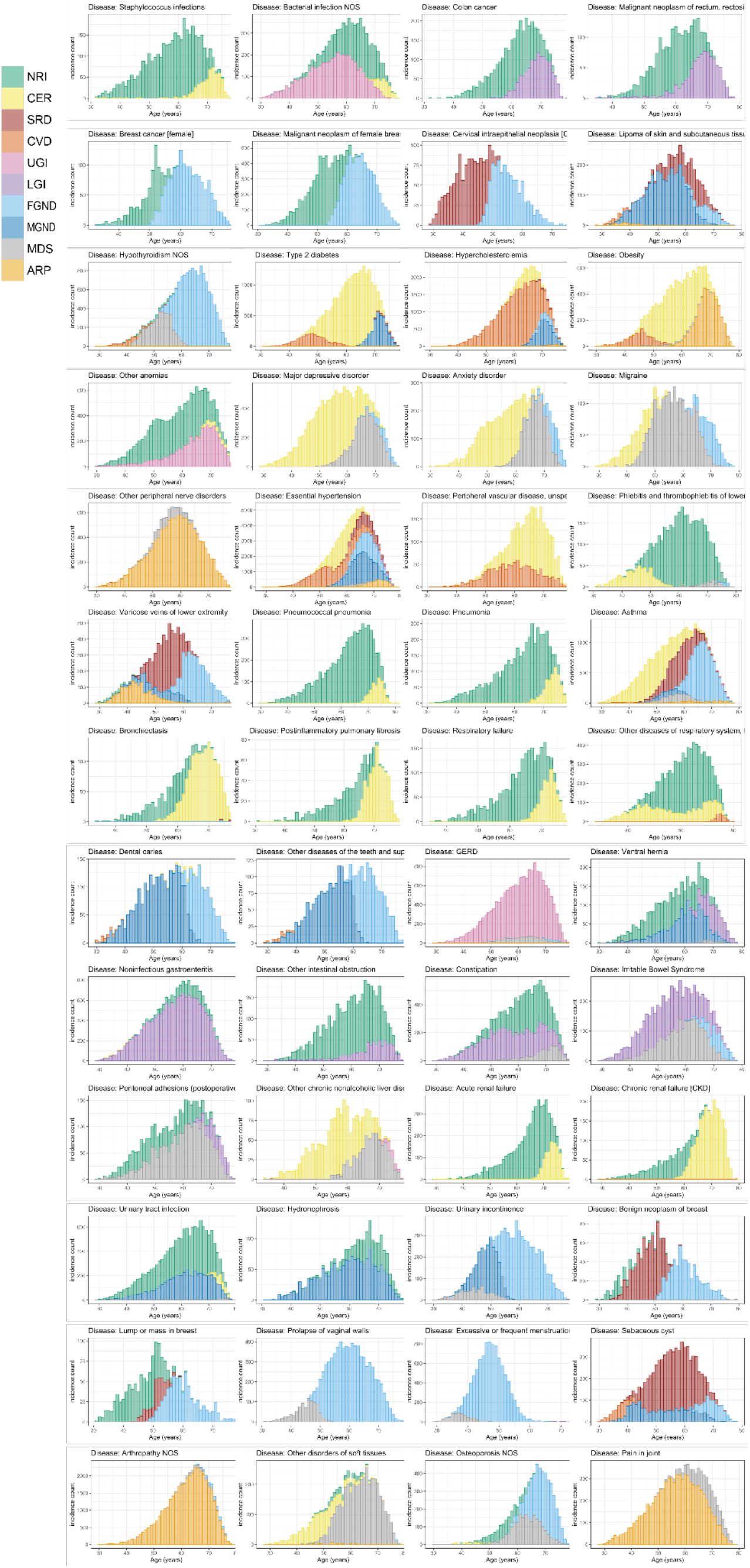
Comorbidity subtype distribution over age for 52 diseases. Diseases shown are ordered by 13 phecode systems: infectious diseases, neoplasms, endocrine/metabolic, hematopoietic, mental disorders, neurological, circulatory system, respiratory, digestive, neoplasms, genitourinary, dermatologic, and musculoskeletal. Numerical results are reported in Supplementary Table 10-11.

**Supplementary Figure 22.**
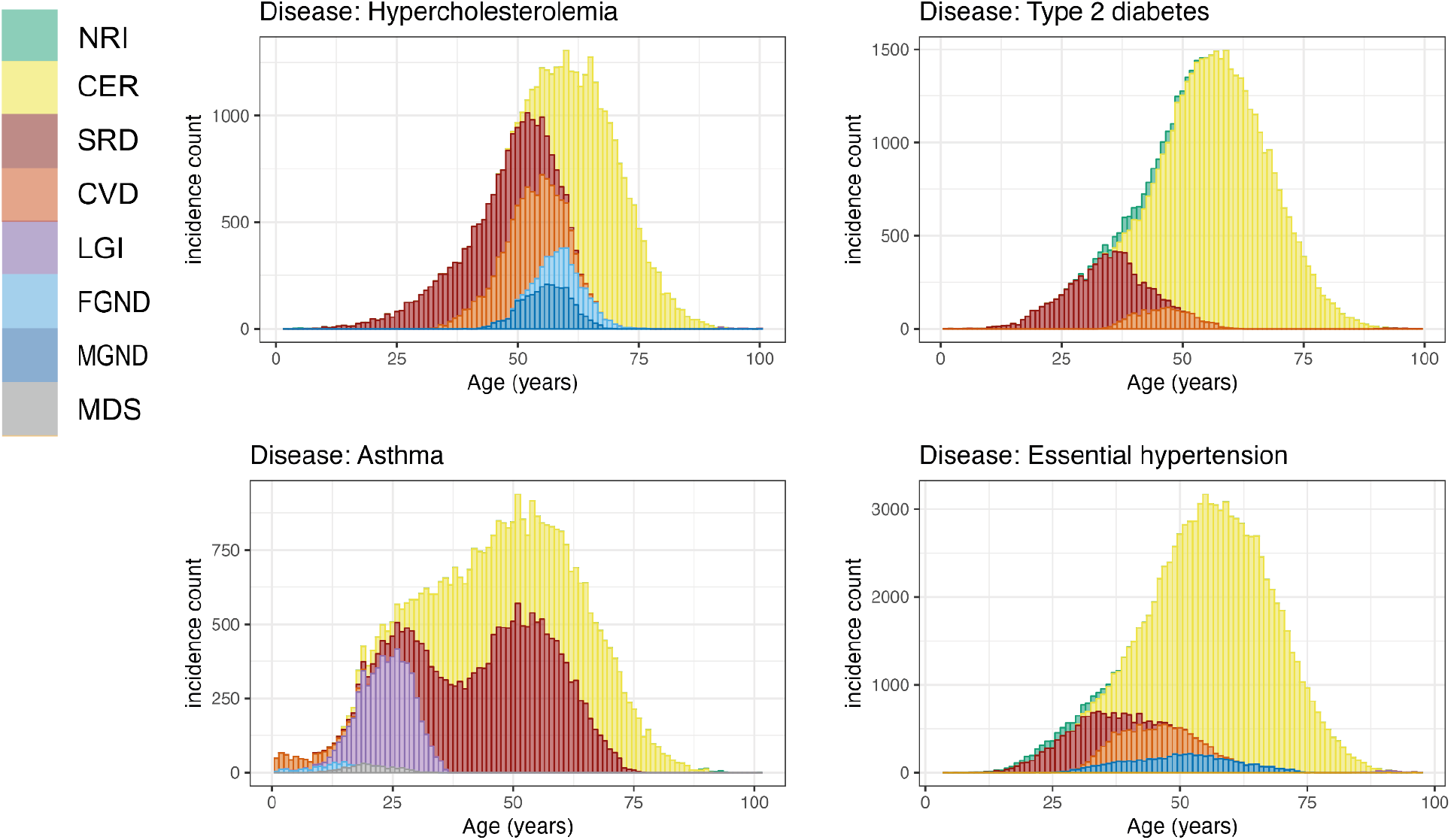
Stacked barplots of age-dependent subtypes in All of Us. Disease topics in All of Us are mapped to their most similar UK Biobank topics; colours are the same as Supplementary Figure 21. The figures are for 4 representative diseases in Figure 6A (type 2 diabetes, asthma, hypercholesterolemia, and essential hypertension); for each disease, we include all subtypes with at least one diagnosis.

**Supplementary Figure 23.**
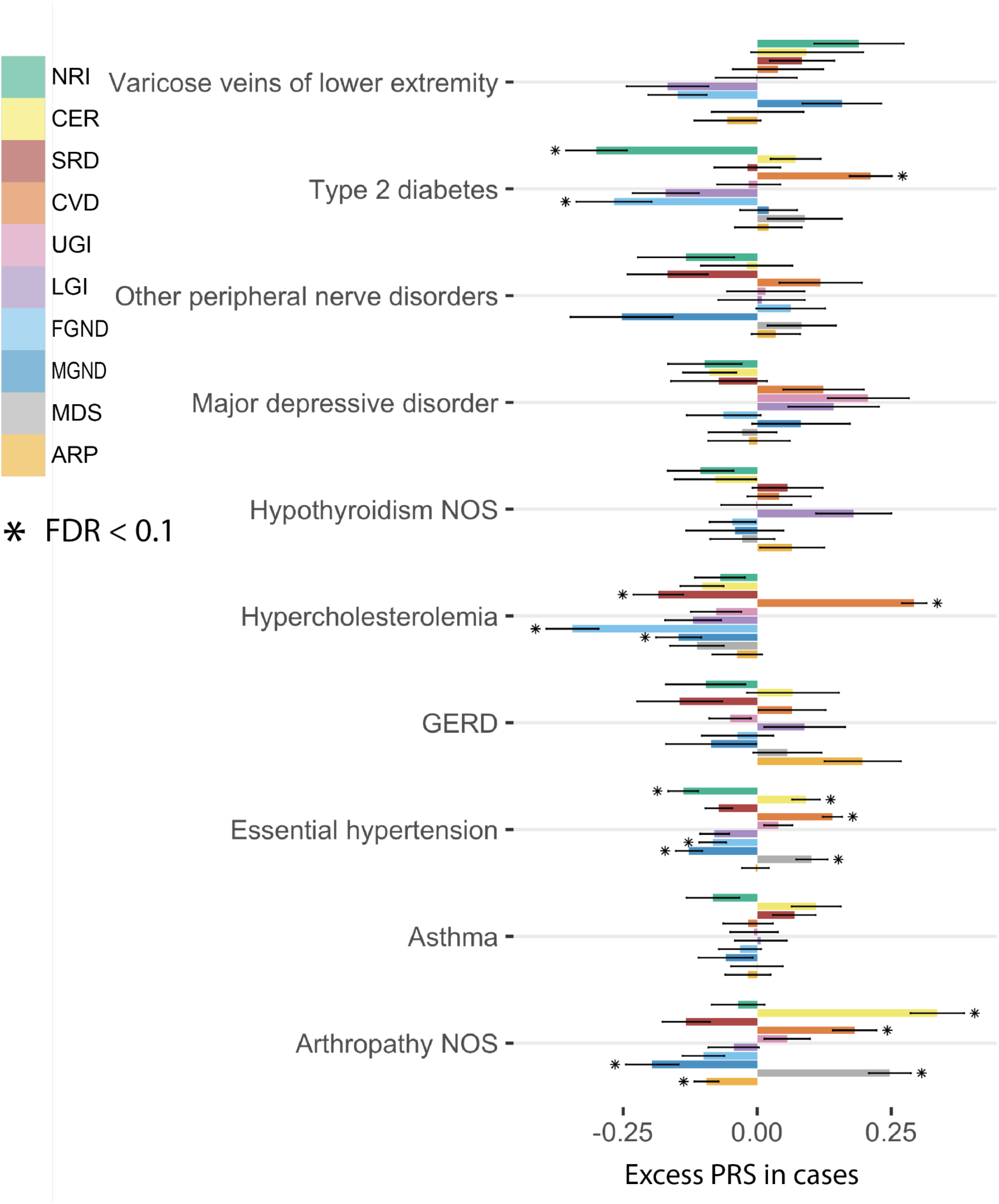
Excess PRS analysis for all topics across 10 diseases (selected by heritability z-score). The bar plot shows the estimated changes in s.d. of PRS per unit changes in the patient topic weight in disease cases. The PRS is estimated using all the cases in British Isle Ancestry. The stars show disease-topic pairs that are significant at FDR = 0.05. Numerical results are reported in Supplementary Table 8.

**Supplementary Figure 24.**
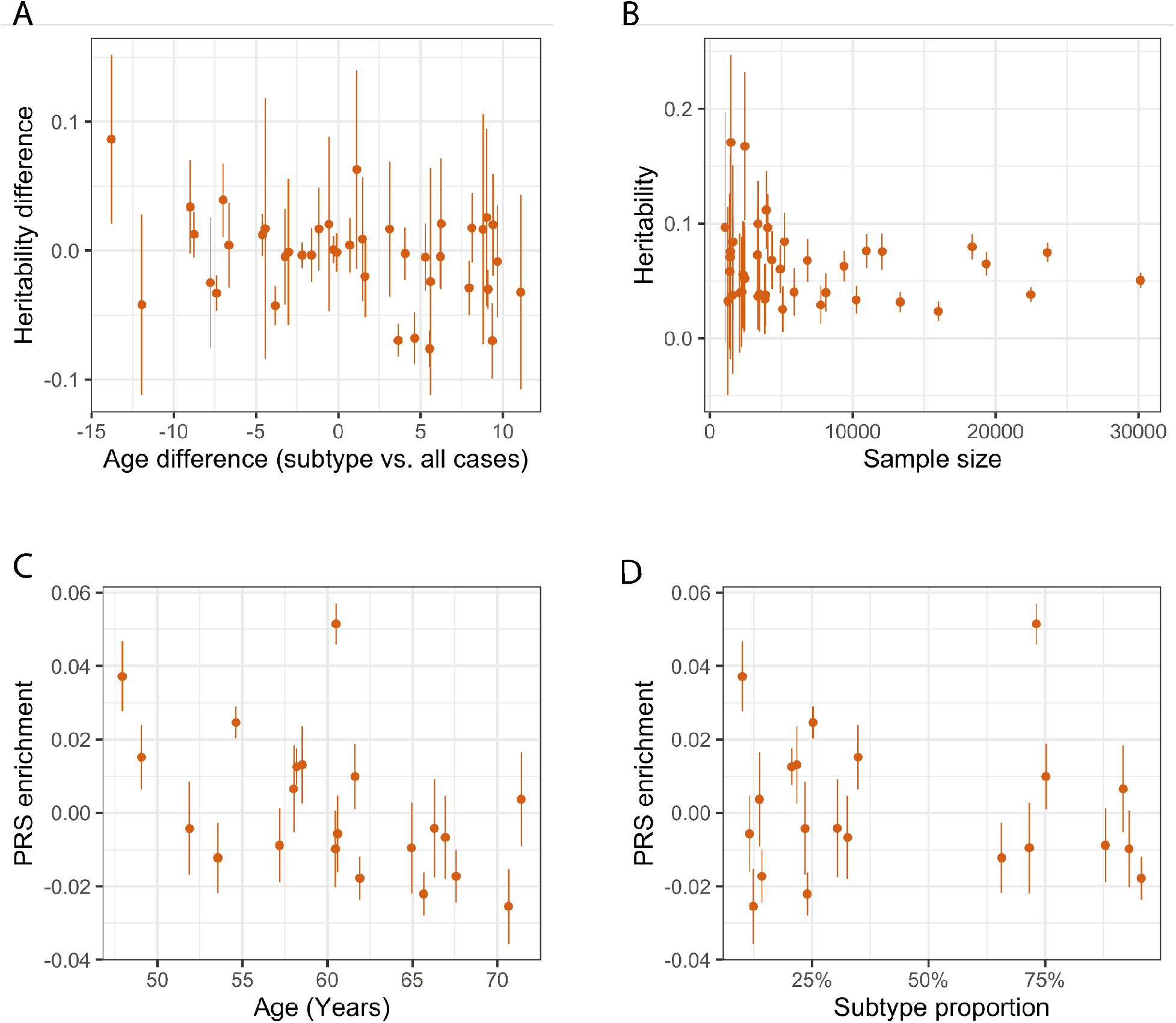
Heritability and PRS are not associated with age and subtype size. (a) We plot heritability deviation from all cases versus age deviation from all cases of 41 subtypes (from 14 diseases that have heritability z-score above 5). The heritability is estimated by first performing mixed-effect association analysis using BOLT-LMM on imputed SNPs from the British Isle Ancestry then using LDSC. (b) Heritability for the subtypes plotted with the sample size of the subtype. (c-d) Excess PRS from Figure 6B plotted against the age and the sample size (denoted by the ratio of samples between subtype and all cases) for the subtypes. The dots and the bars show the mean and 95% confidence interval across all subfigures.

**Supplementary Figure 25.**
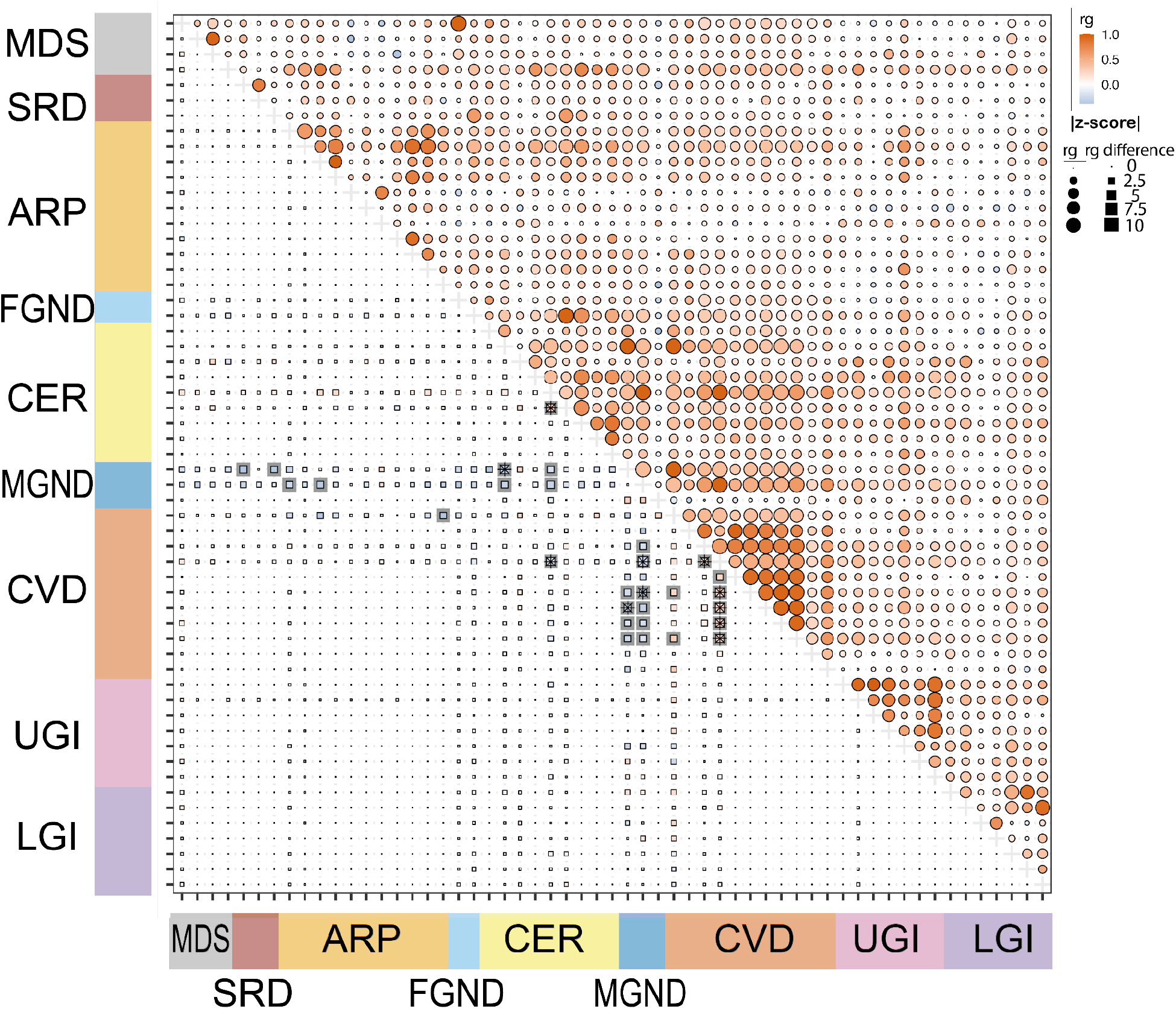
Excess genetic correlation (*r_g_*) between disease subtypes across disease subtypes. Lower left panel shows the *r_g_* of disease-subtype or subtype-subtype pairs subtracted by the *r_g_* of corresponding disease-disease pairs. Each row or column represents a disease or subtype. For disease-disease pairs, the excess *r_g_* is not defined in the lower left panel, since the difference is 0. The upper right panel shows *r_g_* of corresponding disease-disease pairs, where values could be duplicated as the same disease could have multiple subtypes. The 89 diseases or subtypes are chosen here by heritability z-score > 4 and *r_g_* z-score > 4 with at least one other disease or subtypes. We kept 57 rows and columns for better visualisation by removing 32 diseases that have subtypes included. A star means FDR < 0.1, while a shade means a nominal statistical significance at P = 0.05.

**Supplementary Figure 26.**
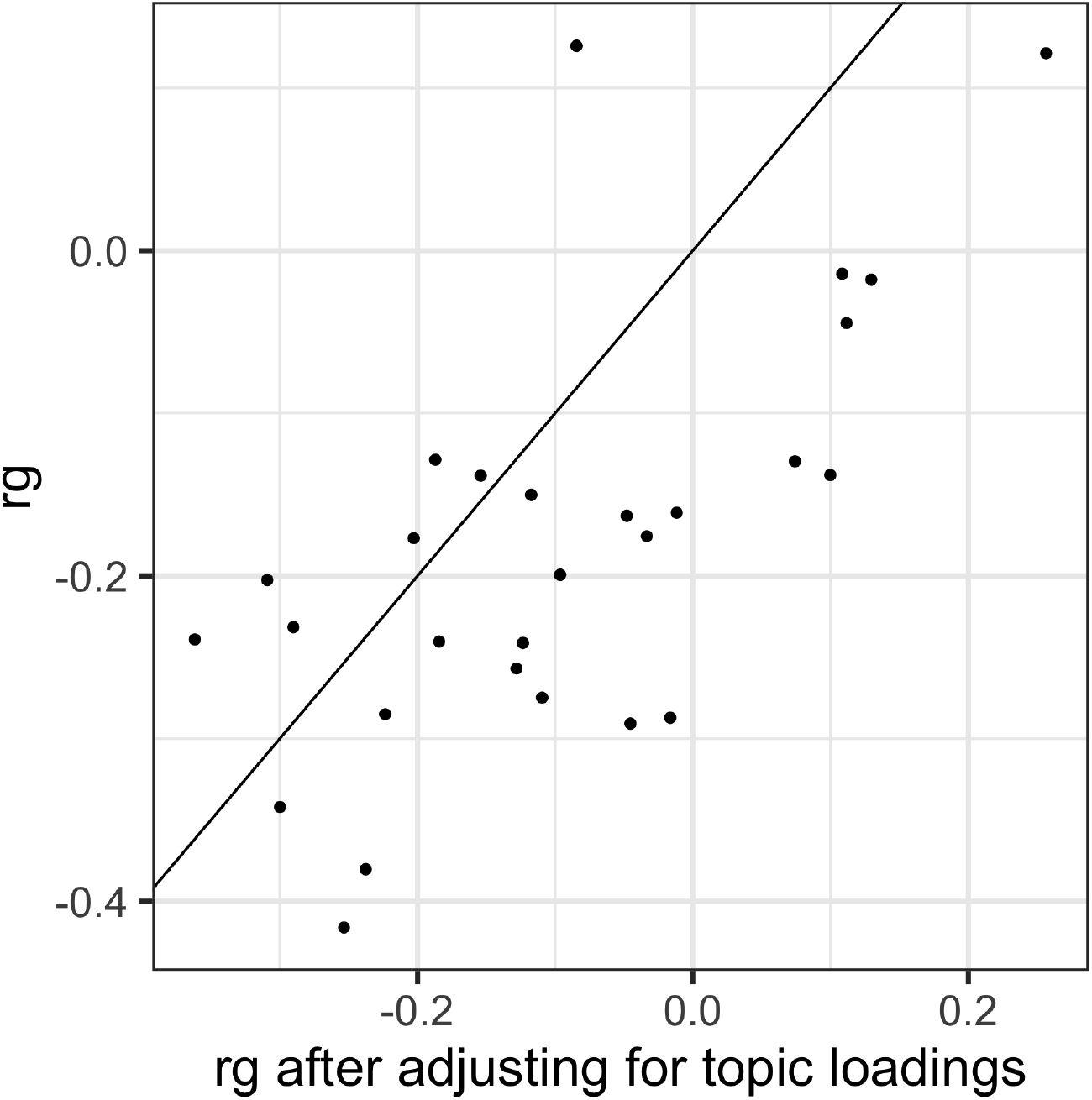
Comparison of excess genetic correlation (*r_g_*) between subtypes using summary statistics from case-control matched by topic weights (x-axis) and not matched by topic weights. The analysis is performed on subtypes of four diseases: type 2 diabetes, hypercholesterolemia, hypertension, and asthma. The excess ***r_g_*** (shown in panel (A) of Figure 7) of two case-control matching strategies across 28 subtype pairs are compared and the effect lies along the diagonal line. The excess genetic correlation attenuated slightly when topic weights are matched, while it can not explain all the excess ***r_g_***.

**Supplementary Figure 27.**
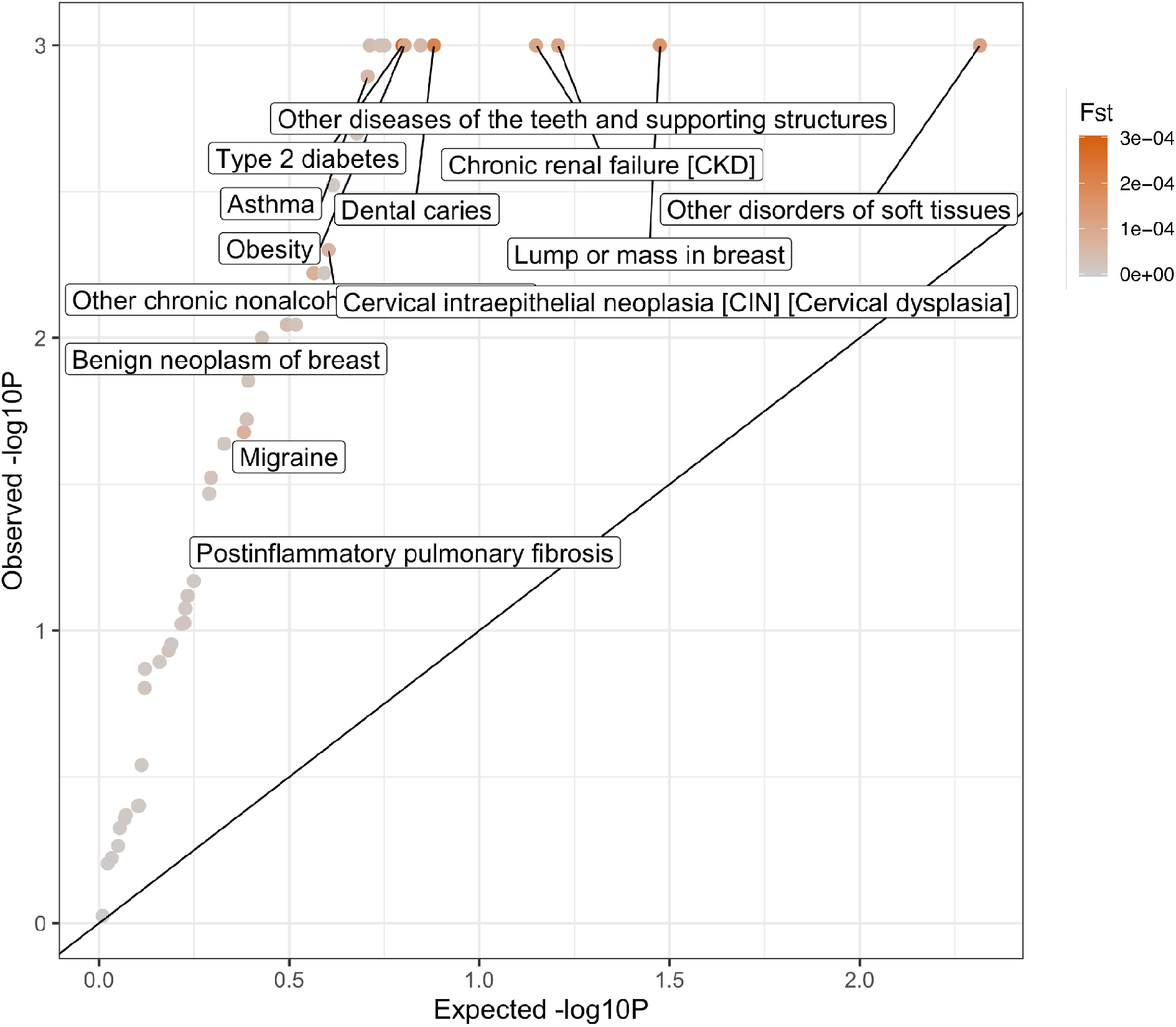
Excess *F_ST_* of disease subtypes compared with controls with matched topic weights. P-values are for testing case-*F_ST_* significantly higher than controls of similar topic weight distribution. The permutation controls are sampled for 1,000 times with the same topic weights distribution and sample size to the disease subtypes. We focus on 49 of the 52 diseases which have more than one subgroup of at least 500 cases. Subtypes are defined based on the max value of the diagnosis-specific topic probability. Three diseases (“hypertension”, “hypercholesterolemia”, and “arthropathy”) are excluded as there are not enough controls that match the topic weights of cases. The colour shows the value of *F_ST_* across subtypes.

**Supplementary Figure 28.**
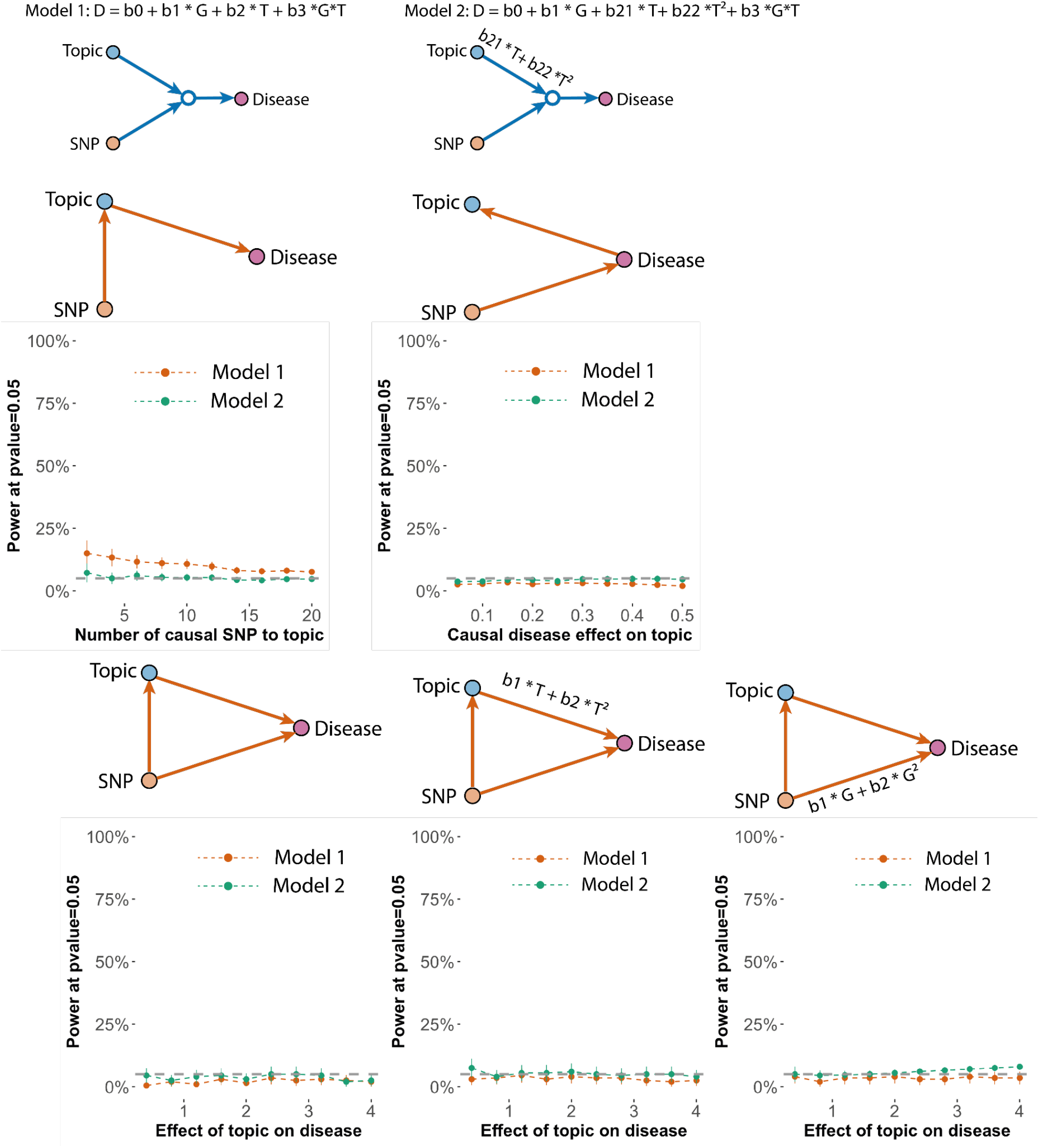
Simulation analysis verified the SNP x topic interaction tests are calibrated when no actual interaction exists. We show the false positive rate of two models under five simulated model structures where no actual interaction exists (Methods). The false positive rate is computed as the power to detect interaction effects using model 1 (red; linear model) and model 2 (green; with non-linear main effect term) under P-value=0.05. The five structures evaluated are (1) SNP causal to topic and topic causal to disease; (2) SNP causal to disease and disease causal to topic; (3) SNP is causal to both topic and disease; (4) and (5) SNP is causal to both topic and disease with nonlinear effects. Genotypes are simulated using the MAF from the 888 disease associated SNPs that were analysed in the SNPxTopic interaction tests.

**Supplementary Figure 29:**
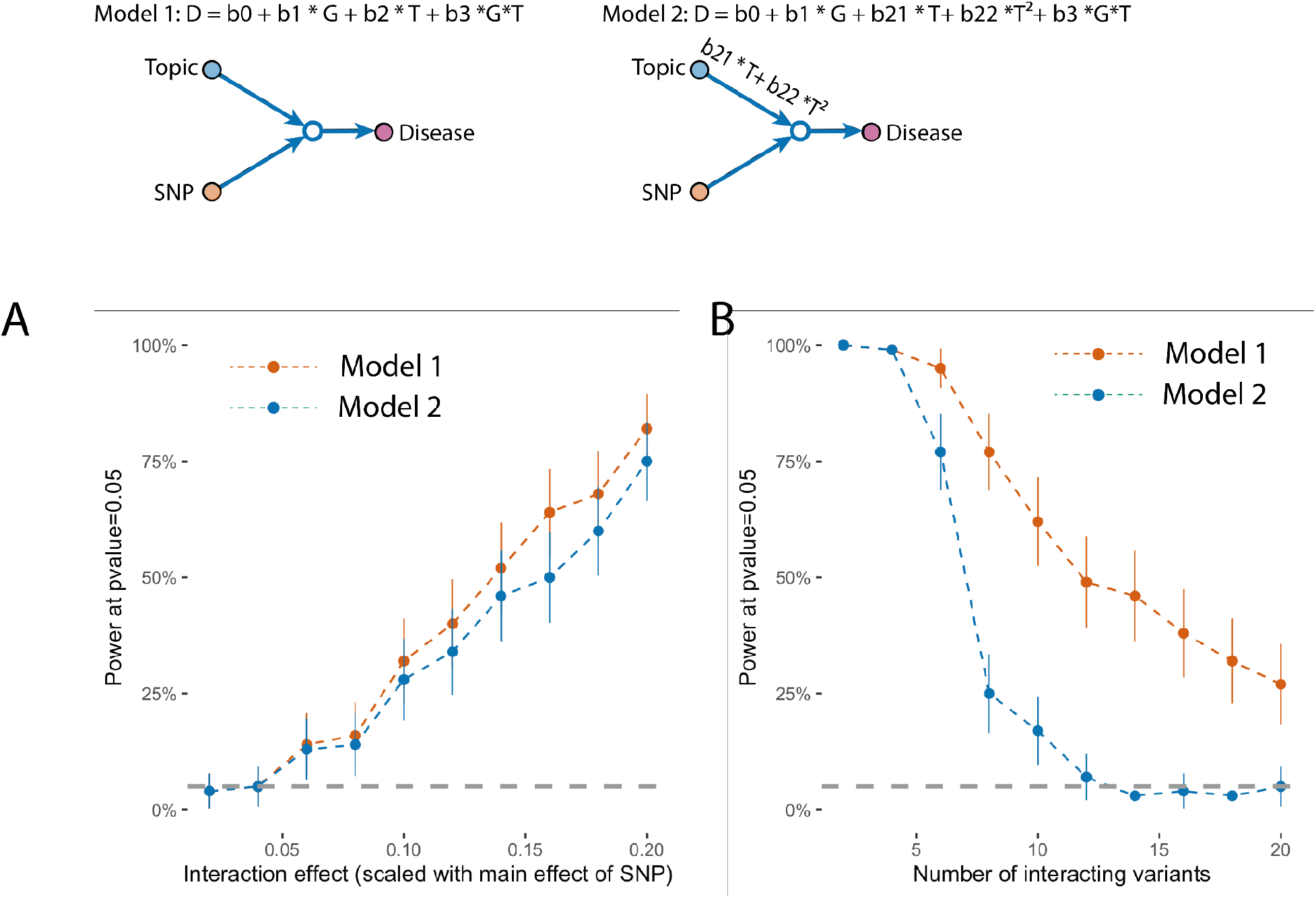
Power to detect true SNP x topic interaction effect under simulation. (a) We simulated data with 10,000 individuals, topic-to-disease main effect size equal to 2, interaction effect from 0.04 to 0.4, and SNP-to-disease main effect size proportional to the interaction effect (0.02 to 0.2); disease diagnoses are generated using gaussian liability with top 20 percentile as cases. We tested for the SNP x topic interaction using model 1 and model 2 and computed the power of discovering the true interaction. (b) We simulated data with 10,000 individuals and an interaction effect equal to 0.4. Instead of simulating a single SNP effect, we simulated 2 to 20 variants that all interact with topic weight. We then test the SNP x Topic interaction in model 1 and model 2 with one variant at a time, which is the same strategy as most GWAS interaction tests. We note the power of model 2 is lower than model 1, while we still choose model 2 as it is better calibrated (Supplementary Figure 28).

**Supplementary Figure 30.**
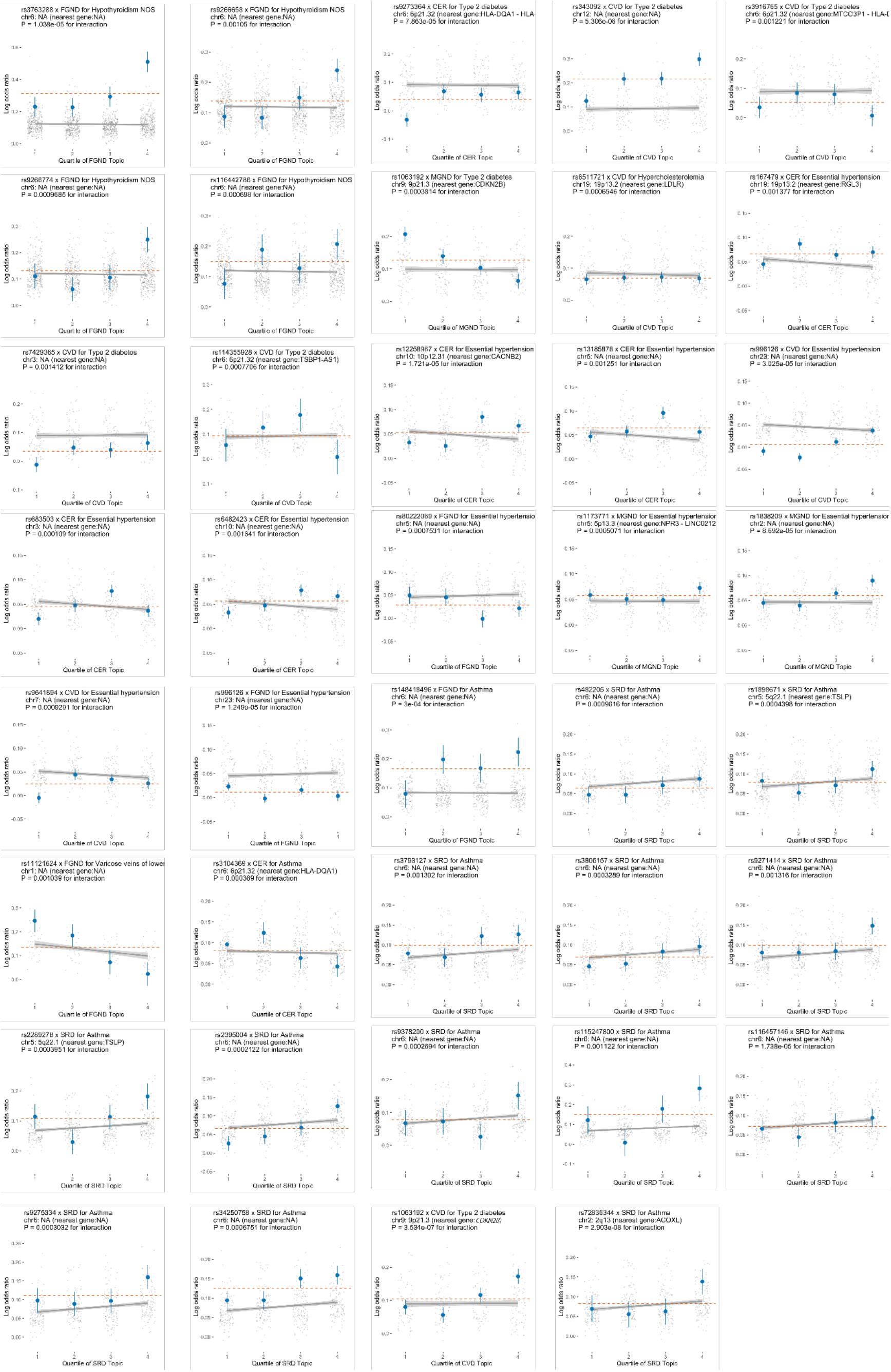
Additional 39 SNPs (mapped genes in the parentheses) that have different effect sizes in different quantiles of topic weights. Blue dots are the effect sizes of the target SNP within each topic weights quartile; grey dots are the effect sizes of background SNPs which are genome-wide significant for the traits but do not have evidence supporting interaction with topic weights (P>0.05). The grey line shows the regression line of the grey dots and its 95% confidence interval. The topic weights (of the topic being tested) are matched for both case and controls within each quartile. Numerical results are reported in Supplementary Table 18.

**Supplementary Figure 31:**
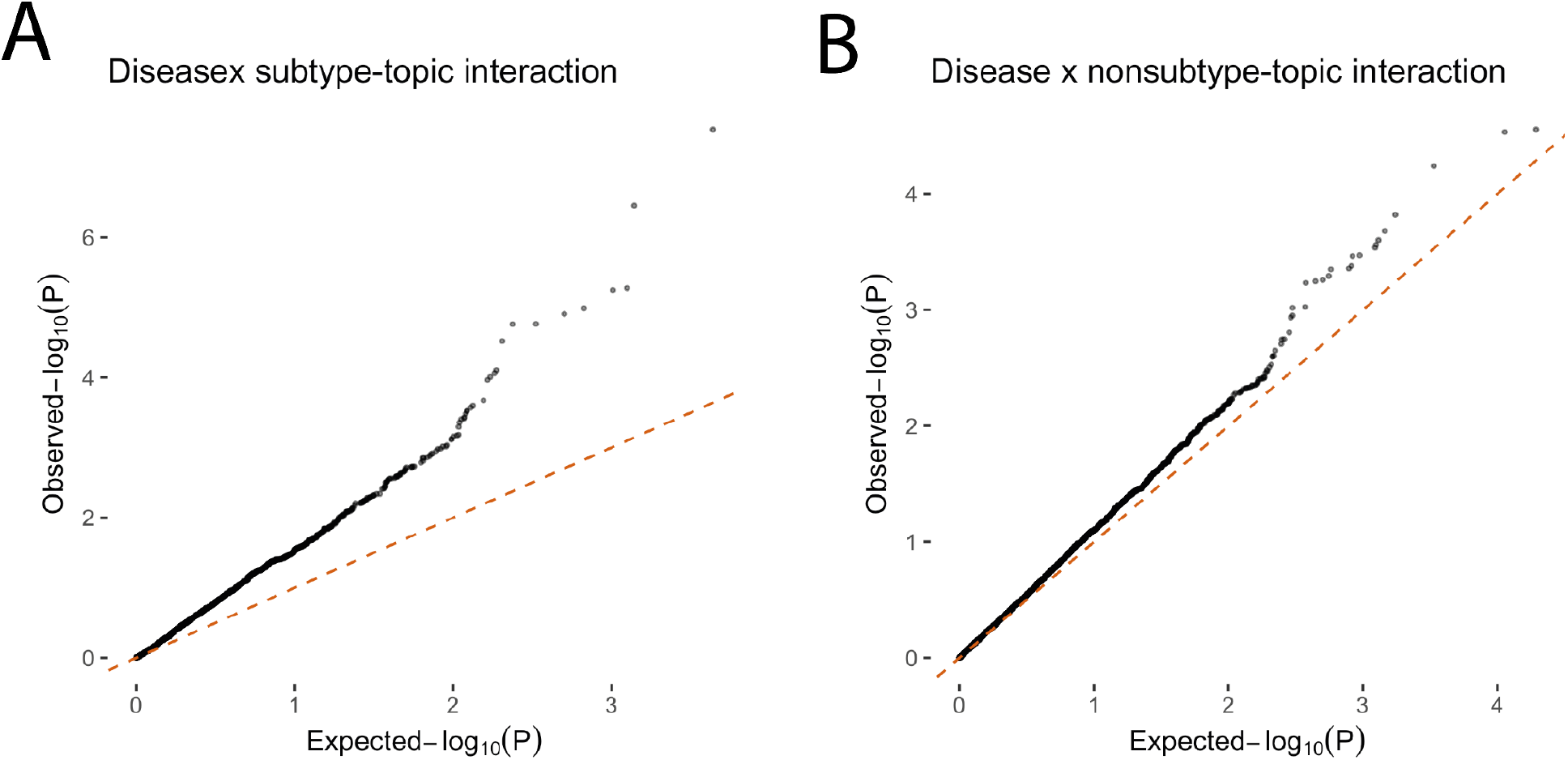
QQ plot of SNP x topic interaction for all GWAS SNPs (P <. *5* × *10*^-^*^8^*). (a) We show the interaction between SNP-topic where the topics define disease subtypes. We focus on the subset of subtypes whose disease have *ℎ^2^* z-score larger than 4 to ensure there is enough GWAS signal for testing. The P-values are for testing the interaction effects with nonlinear topic-to-disease main effects (Model 2 in Supplementary Figure 28). The median for observed p-value is 0.35. (b) As a control to show the calibration of the tests, we plot the QQ-plot over the same set of GWAS SNPs, but over the topic that are not identified as subtypes of the disease by ATM. The median for observed p-value in the Null test is 0.47. The observed small inflation of test statistics (0.47 < 0.5) is caused by the correlation between topics (i.e. a SNP that interacts with a subtype-topic is expected to have weak interaction with other non-subtype topics as the topic weights sum to one).

**Supplementary Figure 32.**
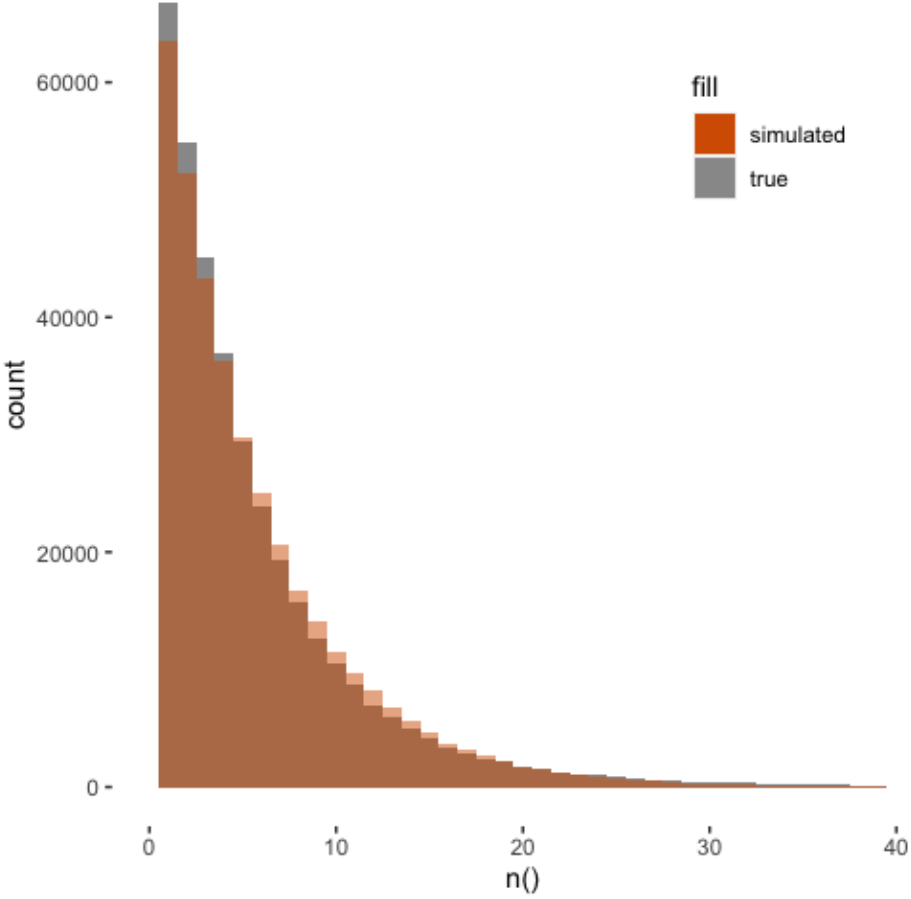
Comparison of simulated diagnosis per individual versus true UK Biobank data. Histogram of number of distinct diseases per patient from the UK Biobank HES dataset and from the simulated exponential distribution with mean = 6.1.

**Supplementary Figure 33.**
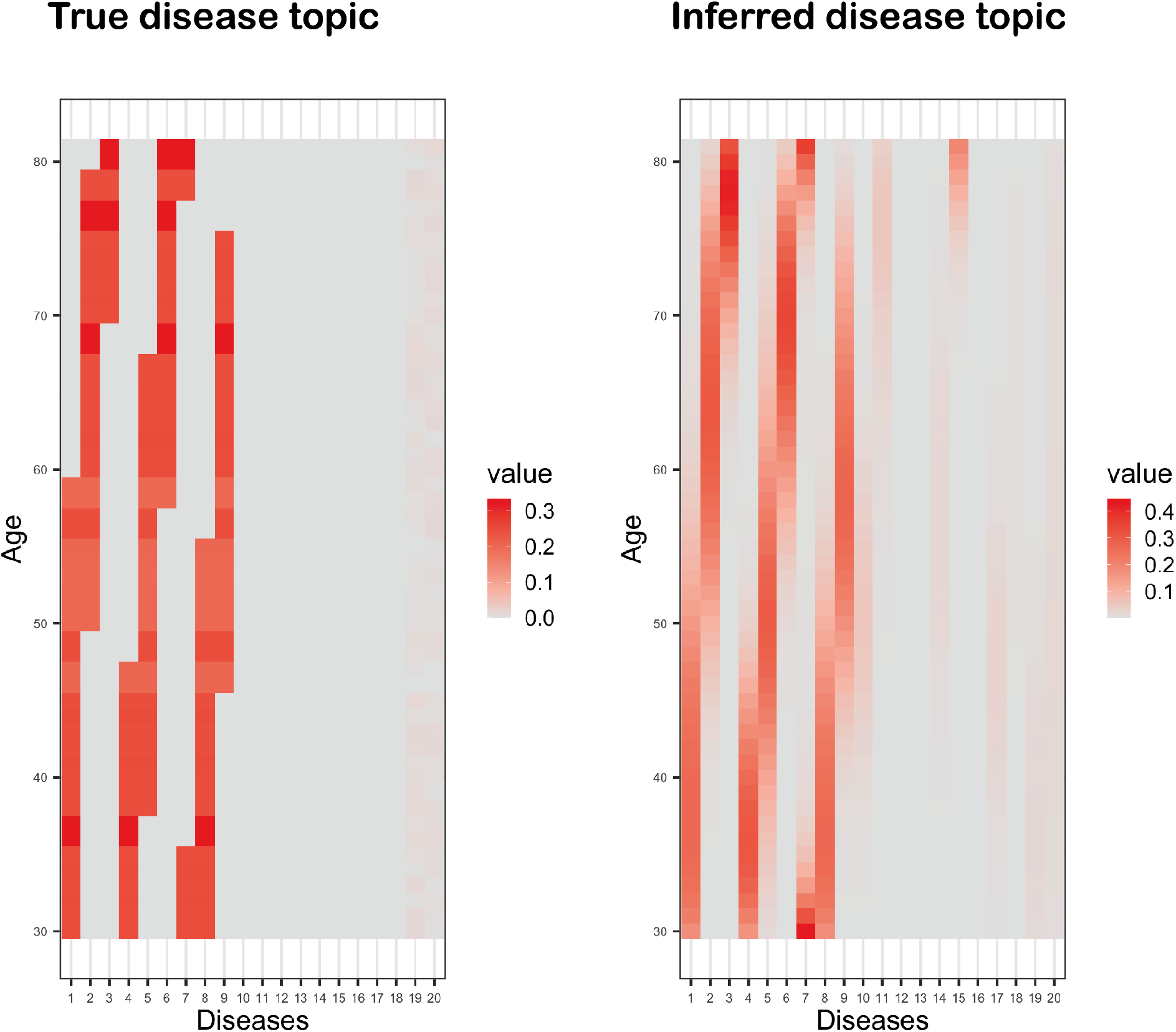
Examples of simulated vs. inferred topic loadings from ATM. Left panel shows the topic loadings used to simulated 10,000 individuals; right panel shows the inferred topic loadings using topic loadings parametrized as cubic polynomials.

