## Supplementary Note for "Age-dependent topic modelling of comorbidities in UK Biobank identifies disease subtypes with differential genetic risk"

Xilin Jiang

April 27, 2023

#### 1 Usage instruction for the supplementary notes

The purpose of this supplementary note is to provide a self-contained explanation of the mathematical basis underlying our topic based model. Therefore, the materials are not meant to provide new discoveries but to help readers to derive all of our inference methods without referring to external materials (though we do listed references to text wherever appropriate). As a consequence, we made no efforts to condense steps, and opt to expand with more details when we feel it is necessary.

#### 2 Generative process of a curve topic model

We constructed a Bayesian hierarchical model to infer latent risk profiles for common diseases. In summary, the model assumes there exist a few disease topics that underlie

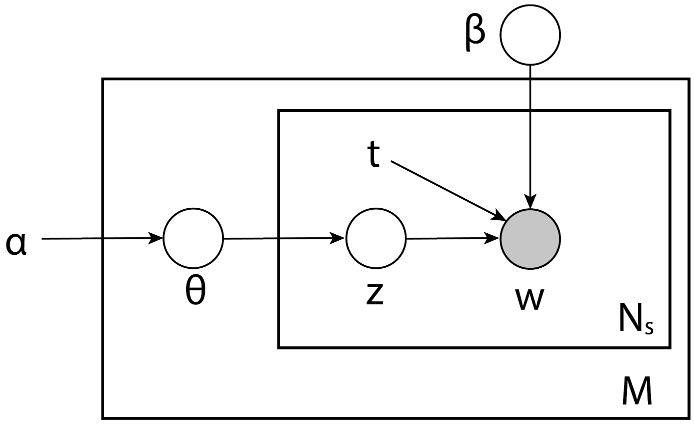

Supplementary Figure 1: Plate notation of ATM generative model. $M$ is the number of subjects, $N_s$ is the number of records within $s^{th}$ subject. All plates (circles) are variables in the generative process, where the plates with shade $w$ is the observed variable and plates without shade are unobserved variables to be inferred. $\theta$ is the topic weight for all individuals; z is diagnosis-specific topic probability; $t$ is the age at onset for each diagnosis; $\beta$ is the topic loadings which are functions of age $t$; $\alpha$ is the (non-informative) hyperparameter of the prior distribution of $\theta$. The generative process is described in the Methods and Supplementary Note.

many common diseases. Each topic is age-evolving and contain risk trajectories for all diseases considered. An individual's risk for each diseases is determined by the weights of all topics. The indices in this note are as follows:

$$s = 1, ..., M;$$

$$n = 1, ..., N_s;$$

$$i = 1, ..., K;$$

$$j = 1, ..., D;$$

where $M$ is the number of subjects, $N_s$ is the number of records within $s^{th}$ subject, $K$ is number of topics, and $D$ is the total number of diseases we are interested in. The generative process (Supplementary Figure 1) is as follows:

- $\theta \in \mathcal{R}^{M \times K}$ is the topic weight for all individuals, each row of which ($\in \mathcal{R}^{K}$) is assumed to be sampled from a Dirichlet distribution with parameter $\alpha$. $\alpha$ is set as a hyper parameter.

$$\theta_s \sim Dir(\alpha).$$

- $\mathbf{z} \in \{1,2,...,K\}^{\sum_s N_s}$ is the topic assignment for each diagnosis $\mathbf{w} \in \{1,2,...,D\}^{\sum_s N_s}$. Note the total number of diagnoses across all patients are $\sum_s N_s$. The topic assignment for each diagnosis is generated from a multinoulli distribution with parameter equal to $s^{th}$ individual topic weight.

$$z_{sn} \sim Multi(\theta_s).$$

- $\beta(t) \in \mathcal{F}(t)^{K\times D}$ is the topic which is $K \times D$ functions of age $t$. $\mathcal{F}(t)$ is the class of functions of $t$. At each plausible $t$, the following is satisfied:

$$\sum_j \beta_{ij}(t) = 1.$$

In practice we use softmax function to ensure above is true and add smoothness by constrain $\mathcal{F}(t)$ to be spline or polynomial functions:

$$\beta_{ij}(t) = \frac{\exp(\boldsymbol{p}_{ij}^T \phi(t))}{\sum_{j=1}^{D} \exp(\boldsymbol{p}_{ij}^T \phi(t))},$$

where $\boldsymbol{p}_{ij} = \{p_{ijd}\},\ d = 1,2,...,P$; $P$ is the degree of freedom than controls the smoothness; $\phi(t)$ is polynomial and spline basis for age $t$.

- $w \in \{1,2,...,D\}^{\sum_s N_s}$ are observed diagnoses. The $n^{th}$ diagnosis of $s^{th}$ individual $w_{sn}$ is sampled from the topic $\beta_{z_{sn}}(t)$ chosen by $z_{sn}$:

$$w_{sn} \sim Multi(\boldsymbol{\beta}_{z_{sn}}(t_{sn})),$$

here $t_{sn}$ is the age of the observed age-at-onset of the observed diagnosis $w_{sn}$.

The value of interest in this model are global topic parameter $\beta$, individual (patient) level topic value $\theta$, and topic value $z$ of each diagnosis. Based on the generative process above, we notice each patient is independent conditional on $\alpha$ and $\beta$. Therefore, we could adopt an EM strategy, where we first estimate $\theta$ and $z$, then estimate $\beta$ which maximise the evidence lower bound.

In the first step we could work on the likelihood function fore each patient to estimate posterior distributions of patient specific variables $\theta$ and $z$. The likelihood function for $s^{th}$ individual is as follows:

$$\begin{aligned}
&\ln p(\mathbf{w},\mathbf{z},\theta|\alpha,\beta) = \ln p(\theta|\alpha) + \sum_{n=1}^{N_s}\{\ln p(z_n|\theta) + \ln p(w_n|z_n,\beta)\},\\
&p(\theta|\alpha) = \frac{\Gamma(\sum_{i=1}^{K}\alpha_i)}{\prod_i^K \Gamma(\alpha_i)} \prod_{i=1}^{K} \theta_i^{\alpha_i - 1},\\
&p(z_n|\theta) = \theta_{1(i=z_n)},\\
&p(w_n|z_n,\beta(t_n)) = \beta_{1(i=z_n),1(j=w_n)}(t_n).
\end{aligned} \tag{1}$$

Due to the computational cost of simultaneously modelling hundreds of diseases in the biobank and the inference accuracy consideration (which we will explain in section 4), we adopted a collapsed variational methods for this step. The method is motivated by [1]. Detailed explanation on why we chose this rather sophisticated methods rather than the commonly used mean filed methods is discussed in section 4, for those interested.

In the second step, we treated the $\beta$ as parameter of the model and seek to maximise the evidence function $p(\mathbf{w}|\alpha, \beta)$ (obtained by integrate out $\theta$ and $z$ from the likelihood function). Directly working on the evidence function is implausible, therefore we work on the evidence lower bound, where we made use of the posterior distribution $q(\mathbf{z}, \theta)$ estimated in previous step.

$$\mathcal{L}(\mathbf{z}, \theta, \beta, \alpha) = \mathbf{E}_q\{\ln p(\mathbf{w}, \mathbf{z}, \theta|\alpha, \beta) - \ln q(\mathbf{z}, \theta)\}. \tag{2}$$

We will see this is still not easily achieved, therefore we applied a local variational method to find an approximate solution. For an easy introduction to local variational inference, see chapter 10.5 of [2]. Details of the inference will be explained in section 3.

### 3 Inference of model posterior distribution and parameters

The model inference will be performed by alternation an E-step and a M-step. The EM algorithm will guarantee good convergence properties. For both steps, variational methods will be used to approximate the distribution, though the techniques are very different. We have tested under realistic parameters, these approximated distribution are close to the true distribution.

#### 3.1 Collapsed variational inference to estimate patient-level posterior distribution $q(z, \theta)$

The variational inference aims to approximate posterior $p(\mathbf{z}, \theta|\mathbf{w}, \alpha, \beta)$ using variational distributions $q(\mathbf{z}, \theta)$ that has some constraint on, which makes them easier to estimate. The most widely used form of variational distribution is the factorised ones, where we assume target posterior distributions are independently distributed, i.e. $q(\mathbf{z}, \theta) = q(\mathbf{z})q(\theta)$. We will derive the inference using this assumption and compare it with the collapsed variational inference in section 4.

The latent variable model using Dirichlet distribution is typically designed to model text, where a document is equivalent as a patient in our model. A document will have thousands of words (equivalent of our diagnoses), which provides strong information to tolerate stronng assumptions on $q(\mathbf{z}, \theta)$. To estimate a model with less diagnoses per patient, flexible variational distributions are preferred for approximation accuracy. Here we adopted a collapsed variational method, which is is more accurate than the mean-field variational inference method. [1] The idea is to only assume a factorization over $q(\mathbf{z})$, but not between $\mathbf{z}$ and $\theta$. Therefore the assumptions and lower bound of evidence became (note we are considering likelihood function for only the $s^{th}$ patient from now on, as all of patients are independend conditional on $\alpha, \beta$):

$$\begin{aligned} q(\mathbf{z}, \theta) &= q(\theta|\mathbf{z}) \prod_n q(z_n), \\ \mathcal{L}(\mathbf{z}, \theta, \beta, \alpha) &= \mathbf{E}_q\{\ln p(\mathbf{w}, \mathbf{z}, \theta|\alpha, \beta) - \ln q(\mathbf{z}) - \ln q(\theta|\mathbf{z})\} \\ &= \mathbf{E}_{q(z)}\{\mathbf{E}_{q(\theta|z)}\{\ln p(\mathbf{w}, \mathbf{z}, \theta|\alpha, \beta) - \ln q(\theta|\mathbf{z})\} - \ln q(\mathbf{z})\} \end{aligned} \tag{3}$$

Maximise $\mathbf{E}_{q(\theta|z)}\{\ln p(\mathbf{w}, \mathbf{z}, \theta|\alpha, \beta) - \ln q(\theta|\mathbf{z})\}$ with respect to $q(\theta|\mathbf{z})$ will give us $q(\theta|\mathbf{z}) = p(\theta|\mathbf{w}, \mathbf{z}, \alpha, \beta)$. The maximisation is achieved similarly to the mean-field approximation where the evidence is decomposed into a lower bound and KL divergence, where lower bound is maximised when KL divergence is 0. Using methods provided in Teh et al, the lower bound could then be simplified to:

$$\mathcal{L}(\mathbf{z}, \theta, \beta, \alpha) = \mathbf{E}_{q(\mathbf{z})}\{\ln p(\mathbf{w}, \mathbf{z}|\alpha, \beta) - \ln q(\mathbf{z})\}$$

The optimisation of this lower bound is same to those provided in collapsed gibbs sampling, where we first marginalise over $\theta$.

$$\begin{aligned} p(\mathbf{w}, \mathbf{z}|\alpha, \beta) &= \int_\theta \frac{\Gamma(\sum_{i=1}^K \alpha_i)}{\prod_i^K \Gamma(\alpha_i)} \prod_{i=1}^K \theta_i^{\alpha_i + \sum_n z_{ni} - 1} \cdot \prod_{i=1}^K \prod_{j=1}^D \beta_{ij}^{\sum_n z_{ni} w_{nj}} \\ &= \frac{\Gamma(\sum_{i=1}^K \alpha_i)}{\prod_i^K \Gamma(\alpha_i)} \frac{\prod_i^K \Gamma(\alpha_i + \sum_n z_{ni})}{\Gamma(\sum_{i=1}^K \alpha_i + N_s)} \cdot \prod_{i=1}^K \prod_{j=1}^D \beta_{ij}^{\sum_n z_{ni} w_{nj}}. \end{aligned} \tag{4}$$

From this marginal complete data likelihood, we could derive the conditional distribution $p(z_{n'} = k|\mathbf{z}_{\neg \mathbf{n}'}, \mathbf{w}, \alpha, \beta)$ (as in collapsed gibbs sampling) to evaluate the dependency within $\mathbf{z}$. Here $\neg n'$ refer to indices of all words excluding $n'$.

$$\begin{aligned} p(z_{n'}|\mathbf{z}_{\neg \mathbf{n}'}, \mathbf{w}, \alpha, \beta) &= \frac{p(z_{n'}, \mathbf{z}_{\neg \mathbf{n}'}, \mathbf{w}|\alpha, \beta)}{p(\mathbf{z}_{\neg \mathbf{n}}, \mathbf{w}|\alpha, \beta)} \\ &= \frac{p(\mathbf{z}, \mathbf{w}|\alpha, \beta)}{p(\mathbf{z}_{\neg \mathbf{n}}, w_{\neg n}|\alpha, \beta) p(w_{n'}|\alpha, \beta)} \\ &\propto \frac{\prod_i^K \Gamma(\alpha_i + \sum_n z_{ni}) \prod_{i=1}^K \prod_{j=1}^D \beta_{ij}^{\sum_n z_{ni} w_{nj}}}{\prod_i^K \Gamma(\alpha_i + \sum_{\neg n'} z_{ni}) \prod_{i=1}^K \prod_{j=1}^D \beta_{ij}^{\sum_{\neg n'} z_{ni} w_{nj}}} \\ &\propto \prod_i^K (\alpha_i + \sum_{n \in \neg n'} z_{ni})^{z_{n'i}} \prod_{i=1}^K \prod_{j=1}^D \beta_{ij}^{z_{n'i} w_{n'j}}. \end{aligned} \tag{5}$$

For a large $N_s$, $(\alpha_i + \sum_{n\in\neg n'} z_{ni})$ will be approximately the same across $n'$, therefore $z_{n'}$ will be less dependent on $\mathbf{z}_{\neg n'}$.

$$\lim_{N_s\to\infty} p(z_{n'}|\mathbf{z}_{\neg\mathbf{n'}}, \mathbf{w}, \alpha, \beta) \propto \prod_{i}^{K} \Big[(\alpha_i + N_s\theta_i) \prod_{j=1}^{D} \beta_{ij}^{w_{n'j}}\Big]^{z_{n'i}},$$

where distribution of $\mathbf{z}$ factorises over $n$ within a single subjects. Therefore, the $q^*(\mathbf{z})$ in equation 17 which factorises over $n$ could approximate $p(\mathbf{z}|\mathbf{w}, \alpha, \beta)$ accurately. However, $N_s$ is likely to be small in the patient dataset, therefore the mean-field approximation in equation 17 would be less accurate as it does not include any dependency between $\mathbf{z}_n$ and $\mathbf{z}_{\neg\mathbf{n'}}$. We therefore adopt the strategies proposed by Teh et al to use variatioanal distribution to approximate the marginal distribuion in equation 4:

$$\begin{aligned}
\ln q^*(z_{n'}) &= \mathbf{E}_{q(\mathbf{z}_{\neg\mathbf{n'}})}\{\ln p(\mathbf{w}, \mathbf{z}|\alpha, \beta)\} \\
&= \mathbf{E}_{q(\mathbf{z}_{\neg\mathbf{n'}})}\{\sum_{i=1}^{K} \ln\Gamma(\alpha_i + \sum_{n} z_{ni}) + \sum_{i=1}^{K}\sum_{j=1}^{D} z_{n'i}w_{n'j}\ln\beta_{ij}\} + const(z_{n'}) \\
&= \sum_{i=1}^{K} z_{n'i}\Big(\mathbf{E}_{q(\mathbf{z}_{\neg\mathbf{n'}})}\{\ln(\alpha_i + \sum_{n\in\neg n'} z_{ni})\} + \sum_{j=1}^{D} w_{n'j}\ln\beta_{ij}\Big) + const(z_{n'})
\end{aligned} \tag{6}$$

We now have the form of multinomial distribution of $z_{n'}$. The key lies in how to estimate $\mathbf{E}_{q(\mathbf{z}_{\neg\mathbf{n'}})}\{\ln(\alpha_i + \sum_{n\in\neg n'} z_{ni})\}$. Teh et al [1] proposed a Gaussian approximation which could improve computation efficiency by magnitudes. We first expand $\ln(\alpha_i + \sum_{n\in\neg n'} z_{ni})$ by Taylor expansion:

$$\ln(\alpha_i + \sum_{n\in\neg n'} z_{ni}) = \ln(\alpha_i + n_0) + \frac{\sum_{n\in\neg n'} z_{ni} - n_0}{(\alpha_i + n_0)} - \frac{(\sum_{n\in\neg n'} z_{ni} - n_0)^2}{2(\alpha_i + n_0)^2},$$

where we included only first two terms. If setting $n_0 = \mathbf{E}_{q(\mathbf{z}_{\neg\mathbf{n'}})}\{\sum_{n\in\neg n'} z_{ni}\} = \sum_{n\in\neg n'} \mathbf{E}_q\{z_{ni}\}$, we get:

$$\mathbf{E}_{q(\mathbf{z}_{\neg\mathbf{n'}})}\{\ln(\alpha_i + \sum_{n\in\neg n'} z_{ni})\} = \ln(\alpha_i + n_0) - \frac{Var_q[\sum_{n\in\neg n'} z_{ni}]}{2(\alpha_i + n_0)^2}. \tag{7}$$

where, $Var_q[\sum_{n\in\neg n'} z_{ni}] = \sum_{n\in\neg n'}(1 - \mathbf{E}_q\{z_{ni}\})\mathbf{E}_q\{z_{ni}\}$ .Plugging this into equation 6 and notice the normalization of multinomial distribution, we get:

$$z_{n'i} \sim \mathbf{Cat}\Big(\frac{(\alpha_i + n_0)\exp\big(-\frac{Var_q[\sum_{n\in\neg n'} z_{ni}]}{2(\alpha_i+n_0)^2} + \sum_{j=1}^{D} w_{n'j}\ln\beta_{ij}\big)}{\sum_{i=1}^{K}(\alpha_i + n_0)\exp\big(-\frac{Var_q[\sum_{n\in\neg n'} z_{ni}]}{2(\alpha_i+n_0)^2} + \sum_{j=1}^{D} w_{n'j}\ln\beta_{ij}\big)}\Big). \tag{8}$$

For prediction tasks, the posterior $\theta$ could be evaluated using distribution of $\mathbf{z}$.

$$\theta \sim \mathbf{Dir}(\alpha + \sum_{n=1}^{N_s} \mathbf{E}_q\{z_n\})$$

The evidence lower bound over all subjects is as follows:

$$\begin{aligned}
\mathcal{L}(\mathbf{z}, \theta, \beta, \alpha) =& \mathbf{E}_{q(\mathbf{z})}\{\ln p(\mathbf{w}, \mathbf{z}|\alpha, \beta) - \ln q(\mathbf{z})\} \\
=& \sum_{s=1}^{M} \Big( \ln \Gamma(\sum_{i=1}^{K} \alpha_i) - \sum_{i}^{K} \ln \Gamma(\alpha_i) - \ln \Gamma(N_s + \sum_{i=1}^{K} \alpha_i) + \\
& \sum_{i=1}^{K} \mathbf{E}_{q(\mathbf{z})}\{\ln \Gamma(\alpha_i + \sum_{n} z_{ni})\} + \\
& \sum_{n=1}^{N_s} \sum_{i=1}^{K} \mathbf{E}\{z_{sni}\} \sum_{j=1}^{D} w_{snj} \ln \beta_{ij} \Big) - \\
& \sum_{s=1}^{M} \Big( \sum_{n=1}^{N_s} \sum_{i=1}^{K} \mathbf{E}\{z_{ni}\} \ln \mathbf{E}\{z_{ni}\} \Big),
\end{aligned} \tag{9}$$

where we need to approximate $\mathbf{E}_{q(\mathbf{z})}\{\ln \Gamma(\alpha_i + \sum_n z_{ni})\}$. Making use of the Stirling's approximation, we found $\ln \Gamma(z) = (z - \frac{1}{2}) \ln(z) - z + \frac{1}{12z} + \frac{1}{2} \ln(2\pi)$ could approximate $\ln \Gamma(z)$ accurately for $z > 1$. Therefore, by plugging in Stirling's approximation and reuse equation 7 we could approximate this expectation:

$$
\begin{aligned}
\mathbf{E}_{q(\mathbf{z})}\{\ln\Gamma(\alpha_i+\sum_n z_{ni})\} =& \mathbf{E}_{q(\mathbf{z})}\{(\alpha_i+\sum_n z_{ni})\ln(\alpha_i+\sum_n z_{ni}) \\
&-\frac{1}{2}\ln(\alpha_i+\sum_n z_{ni})-(\alpha_i+\sum_n z_{ni})+\frac{1}{12(\alpha_i+\sum_n z_{ni})}+\frac{1}{2}\ln(2\pi)\} \\
=& \mathbf{E}_{q(\mathbf{z})}\{(\alpha_i+n_0)\ln(\alpha_i+n_0)+\frac{(\sum_n z_{ni}-n_0])^2}{2(\alpha_i+n_0)} \\
&-\frac{1}{2}\ln(\alpha_i+n_0)+\frac{(\sum_n z_{ni}-n_0])^2}{4(\alpha_i+n_0)2} \\
&-(\alpha_i+\sum_n z_{ni}) \\
&+\frac{1}{12(\alpha_i+n_0)}+\frac{(\sum_n z_{ni}-n_0])^2}{12(\alpha_i+n_0)^3}+\frac{1}{2}\ln(2\pi)\} \\
=& (\alpha_i+n_0)\ln(\alpha_i+n_0)-\frac{1}{2}\ln(\alpha_i+n_0)-(\alpha_i+n_0)+\frac{1}{12(\alpha_i+n_0)} \\
&+Var_q[\sum_n z_{ni}](\frac{1}{2(\alpha_i+n_0)}+\frac{1}{4(\alpha_i+n_0)2}+\frac{1}{12(\alpha_i+n_0)^3}) \\
&+\frac{1}{2}\ln(2\pi),
\end{aligned}
\tag{10}
$$

Note here the first order terms in Taylor expansion are cancelled after taking the expectation and setting $n_0 = \sum_n \mathbf{E}_q\{z_{sni}\}$. The variance are computed by applying the independent assumption over $q(z_n)$: $Var_q[\sum_n z_{ni}] = \sum_n(1-\mathbf{E}_q\{z_{ni}\})\mathbf{E}_q\{z_{ni}\}$.

##### 3.2 Estimate topic profiles $\beta(t)$

In the conventional topic modeling, the topic values could be estimated by directly maximising the evidence lower bound with a constraint $\sum_{j=1}^{D}\beta_{ij} = 1$, which is described in section 4. Here we estimate the topic as functions of age by parameterising each $\beta_{ij}$ as a function of age. The only related term in likelihood function (equation 1) is:

$$
\ln p(w_n|z_n,\beta(t_n)) = \sum_{i=1}^{K} z_{sni}\sum_{j=1}^{D} w_{snj}\ln\pi(\beta_{ij}(t_n)),
$$

where we use softmax function to ensure topics are each a multinomial distribution:

$$
\pi(\beta_{ij}(t_n)) = \frac{\exp(\boldsymbol{\beta}_{ij}^T\phi(t_n))}{\sum_{j=1}^{D}\exp(\boldsymbol{\beta}_{ij}^T\phi(t_n))}.
$$

We used spline/polynomial functions to model age. The goal is to estimate spline/polynomial coefficients $\boldsymbol{\beta}_{ij} = \{\beta_{ijd}\},\ d = 1, 2, ..., P$, where $P$ is the degree of freedom that controls the smoothness. $\phi(t_n)$ is polynomial or spline basis. Notice here the scale of $\boldsymbol{\beta}_{ij} = \{\beta_{ijd}\}$ does not matter, as we could subtract same intercept from the exponential in both numerator and denominator to change in the scale. However, in practice we put a prior $\mathcal{N}(\boldsymbol{\beta}_{ij}|0, \sigma_0^2\mathbf{I})$ on $\beta_{ij}$ to regularise the search space of the gradient descent optimization described below. Here we choose a non-informative prior with large variance, $\sigma_0^2 = 100$.

To maximise the evidence lower bound, we notice that $\ln(\cdot)$ is a concave function and by Taylor expansion:

$$\ln(\sum_{j=1}^{D} \exp(\boldsymbol{\beta}_{ij}^T \phi(t_{sn}))) \leq \ln \zeta + \zeta^{-1}(\sum_{j=1}^{D} \exp(\boldsymbol{\beta}_{ij}^T \phi(t_{sn})) - \zeta).$$

Therefore, by introducing a variational variable $\zeta$, we find following lower bound of the ELBO function $\mathcal{L}$ with respect to $\boldsymbol{\beta}_{ij}$:

$$\begin{aligned}
\mathcal{L}_{[\beta]} &= \sum_{s=1}^{M} \sum_{n=1}^{N_s} \mathbf{E}_q\{\ln p(w_n|z_n, \beta(t_{sn}))\} \\
&= \sum_{s=1}^{M} \sum_{n=1}^{N_s} \sum_{i=1}^{K} \sum_{j=1}^{D} \Big(\boldsymbol{\beta}_{ij}^T \phi(t_{sn}) - \ln(\sum_{j'=1}^{D} \exp\{\boldsymbol{\beta}_{ij'}^T \phi(t_{sn})\})\Big) \mathbf{E}\{z_{sni}\} w_{snj} \geq \\
&\sum_{s=1}^{M} \sum_{n=1}^{N_s} \sum_{i=1}^{K} \sum_{j=1}^{D} \big(\boldsymbol{\beta}_{ij}^T \phi(t_{sn}) - \zeta_{sni}^{-1} \sum_{j'=1}^{D} \exp\{\boldsymbol{\beta}_{ij'}^T \phi(t_{sn})\} - \ln \zeta_{sni} + 1\big) \mathbf{E}\{z_{sni}\} w_{snj}.
\end{aligned} \tag{11}$$

We could then apply a method called local variational inference to maximise the right hand side of equation 11. We do this by updating $\boldsymbol{\beta}$ and $\zeta$ in turn. Take derivative with respect to $\zeta_{sni}$, we obtained following update:

$$\zeta_{sni} = \sum_{j=1}^{D} \exp\{\boldsymbol{\beta}_{ij}^T \phi(t_{sn})\} \tag{12}$$

In order to update the lower bound with respect to $\boldsymbol{\beta}$, we separate the terms containing $\boldsymbol{\beta}_{ij}$:

$$\mathcal{L}_{[\beta_{ij}]} = \sum_{s=1}^{M} \sum_{n=1}^{N_s} \mathbf{E}\{z_{sni}\} w_{snj} \boldsymbol{\beta}_{ij}^T \phi(t_{sn}) - \sum_{s=1}^{M} \sum_{n=1}^{N_s} \mathbf{E}\{z_{sni}\} \zeta_{sni}^{-1} \exp\{\boldsymbol{\beta}_{ij}^T \phi(t_{sn})\}.$$

There is no analytical solution for $\boldsymbol{\beta}_{ij}$, but the lower bound is convex so we could maximize the lower bound using following gradient information:

$$\nabla_{\boldsymbol{\beta}_{ij}} \mathcal{L}_{[\beta]} = \sum_{s=1}^{M} \sum_{n=1}^{N_s} \Big(\mathbf{E}\{z_{sni}\} w_{snj} - \mathbf{E}\{z_{sni}\} \zeta_{sni}^{-1} \exp\{\boldsymbol{\beta}_{ij}^T \phi(t_{sn})\}\Big) \phi(t_{sn}) \tag{13}$$

The gradient information of $\mathcal{L}_{[\beta_{ij}]}$ allows efficient numeric estimation of $\boldsymbol{\beta}_{ij}$. However, evaluating $\mathcal{L}_{[\beta_{ij}]}$ and $\nabla_{\boldsymbol{\beta}_{ij}}\mathcal{L}_{[\beta]}$ is computational expensive due to $\exp\{\boldsymbol{\beta}_{ij}^T\phi(t_{sn})\}$, which require looping through $s, n$ (the entire records set over all subjects!). The gradient descent methods for estimating $\boldsymbol{\beta}_{ij}$ requires evaluating$\mathcal{L}_{[\beta_{ij}]}$ and $\nabla_{\boldsymbol{\beta}_{ij}}\mathcal{L}_{[\beta]}$ at each gradient step, which prohibit scaling up the model to large data set. To solve this problem in practice we discretise $t_{sn}$ into years which allows us to pre-compute the sum of $\mathbf{E}\{z_{sni}\}\zeta_{sni}^{-1}$ over all incidences that happened at each age year. For each new $\boldsymbol{\beta}_{ij}$, we could then sum over all years, which reuses the sums computed. This trick significant reduced the computation cost of evaluating $\mathcal{L}_{[\beta_{ij}]}$ and $\nabla_{\boldsymbol{\beta}_{ij}}\mathcal{L}_{[\beta]}$ which makes the estimation of age topics over the entire UK Biobank HES possible. In conclusion, we could update $\boldsymbol{\beta}$ using following psuedo-code:

**Algorithm 1:** Maximize local variationl lower bound

```
initialization;
for i ← 1 to K do
    for j ← 1 to D do
        Update ζ_sni = Σ_{j=1}^{D} exp{β_ij^T φ(t_sn)} ;
        Update β_ij to maximize L_[β_ij] ;
    end
end
```

Note here we need to update $\zeta_{sni}$ for each $j$, while in practice we only update $\zeta_{sni}$ once for each optimization of $\boldsymbol{\beta}$ to allow parallel computation over $j$.

The above computation provide a point estimate for $\boldsymbol{\beta}$, which we adopted when applying our methods to empirical data. We also provide mathematical derivation for posterior distributions of $\boldsymbol{\beta}$, though we did not present results on empirical data and the full Bayesian method is **not** implemented in ATM software, due to computational cost. We use a Gaussian prior for $\beta_{ijd}$ and a full variational inference of $\beta$ is on following lower bound:

$$\begin{aligned}
\mathcal{L}_{[\beta]} &= \sum_{s=1}^{M}\sum_{n=1}^{N_s}\mathbf{E}_q\{\ln p(w_n|z_n,\beta(t_{sn}))\} + \sum_{i=1}^{K}\sum_{j=1}^{D}\sum_{d=1}^{P}\Big(\mathbf{E}_q\{\ln p(\boldsymbol{\beta}_{ijd})\} - \mathbf{E}_q\{\ln q(\boldsymbol{\beta}_{ijd})\}\Big) \\
&= \sum_{s=1}^{M}\sum_{n=1}^{N_s}\sum_{i=1}^{K}\sum_{j=1}^{D}\big(\mathbf{E}_q\{\boldsymbol{\beta}_{ij}\}^T\phi(t_{sn}) - \mathbf{E}_q\{\ln(\sum_{j=1}^{D}\exp(\boldsymbol{\beta}_{ij}^T\phi(t_{sn})))\}\big)\mathbf{E}\{z_{sni}\}w_{snj} - \\
&\frac{1}{2\sigma^2}\sum_{i=1}^{K}\sum_{j=1}^{D}\sum_{d=1}^{P}\mathbf{E}_q\{\boldsymbol{\beta}_{ijd}^2\} - \sum_{i=1}^{K}\sum_{j=1}^{D}\sum_{d=1}^{P}\mathbf{E}_q\{\ln q(\boldsymbol{\beta}_{ijd})\} \geq \\
&\sum_{s=1}^{M}\sum_{n=1}^{N_s}\sum_{i=1}^{K}\sum_{j=1}^{D}\big(\mathbf{E}_q\{\boldsymbol{\beta}_{ij}\}^T\phi(t_{sn}) - \zeta_{sni}^{-1}\sum_{j=1}^{D}\mathbf{E}_q\{\exp(\boldsymbol{\beta}_{ij}^T\phi(t_{sn}))\} - \ln\zeta_{sni} + 1\big)\mathbf{E}\{z_{sni}\}w_{snj} - \\
&\frac{1}{2\sigma^2}\sum_{i=1}^{K}\sum_{j=1}^{D}\sum_{d=1}^{P}\mathbf{E}_q\{\boldsymbol{\beta}_{ijd}^2\} - \sum_{i=1}^{K}\sum_{j=1}^{D}\sum_{d=1}^{P}\mathbf{E}_q\{\ln q(\boldsymbol{\beta}_{ijd})\}.
\end{aligned} \tag{14}$$

Following [3], we assumed an independent variational Gaussian distribution for each $\beta_{ijd}$:

$$\beta_{ijd} \sim \mathcal{N}(\lambda_{ijd}, \nu_{ijd}^2),$$

and observe the moment-generating function of Guanssian distribution is:

$$\mathbf{E}_q\{\exp(\boldsymbol{\beta}_{ijd}\phi_d(t_{sn}))\} = \exp\Big(\phi_d(t_{sn})\lambda_{ijd} + \frac{\phi_d^2(t_{sn})\nu_{ijd}^2}{2}\Big),$$

We obtain a tractable lower bound with respect to the variational parameters $\{\zeta_{sni}, \lambda_{ijd}, \nu_{ijd}^2\}$:

$$\begin{aligned}
\mathcal{L}_{\zeta,\lambda,\nu^2} &= \sum_{s=1}^{M}\sum_{n=1}^{N_s}\sum_{i=1}^{K}\sum_{j=1}^{D}\big(\lambda_{ij}^T\phi(t_{sn}) - \zeta_{sni}^{-1}\sum_{j=1}^{D}\exp\{\sum_{d=1}^{P}\Big(\phi_d(t_{sn})\lambda_{ijd} + \frac{\phi_d^2(t_{sn})\nu_{ijd}^2}{2}\Big)\} - \\
&\quad \ln\zeta_{sni} + 1\big)\cdot\mathbf{E}\{z_{sni}\}w_{snj} - \sum_{i=1}^{K}\sum_{j=1}^{D}\sum_{d=1}^{P}\big(\frac{1}{2\sigma^2}\nu_{ijd}^2 + \frac{1}{2}\ln\nu_{ijd}^2\big).
\end{aligned} \tag{15}$$

### 4 Comparison of collapsed variational inference and mean field variational inference

A vast number of inference methods have been developed for models based on original Latent Dirichlet Allocation. The most prominet of which are collapsed gibbs sampling and mean field variational inference. For inference of model with exchangeable variables using extremely large and noisy data set, it is desirable to have a deterministic method such as variational inference. Collapsed variational inference makes less assumptions for approximation, therefore the inferred distributions are strictly closer to the true posterior distributions than the mean-field variational Bayesian methods. We will explain why accuracy would be importance for the data we are considering in section 4.2.

#### 4.1 Mean field variational inference to estimate patient-level posterior distribution $q(z, \theta)$

Please note this section is just a replication of [4] using our notation, which is provided to make the note self-contained. We assume variational distributions for latent variables $\theta$ and $\mathbf{z}$ are independent of each other, then we could get the variational lower bound for the log likelihood of a single subject:

$$\begin{aligned} &q(\mathbf{z}, \theta) = q(\mathbf{z})q(\theta), \\ &\mathcal{L}(\mathbf{z}, \theta, \beta, \alpha) = \mathbf{E}_q\{\ln p(\mathbf{w}, \mathbf{z}, \theta|\alpha, \beta) - \ln q(\mathbf{z}, \theta)\}. \end{aligned} \tag{16}$$

It is straightforward to estimate $q(\mathbf{z})$ and $q(\theta)$ that maximise the lower bound $\mathcal{L}(\mathbf{z}, \theta, \beta, \alpha)$:

$$\begin{aligned} \ln q^*(\theta) &= \ln p(\theta|\alpha) + \mathbf{E}_{q(\mathbf{z})}\{\sum_{n=1}^{N_s} \ln p(z_n|\theta)\} + const \\ &= \sum_{i=1}^{K}(\alpha_i + \sum_{n=1}^{N_s} \mathbf{E}\{z_{ni}\} - 1) \ln \theta_i + const, \\ \ln q^*(\mathbf{z}) &= \mathbf{E}_{q(\theta)}\{\sum_{n=1}^{N_s} \ln p(z_n|\theta)\} + \sum_{n=1}^{N_s} \ln p(w_n|z_n, \beta) + const \\ &= \sum_{n=1}^{N_s}\sum_{i=1}^{K} z_{ni}\Big(\mathbf{E}\{\ln \theta_i\} + \sum_{j=1}^{D} w_{nj} \ln \beta_{ij}\Big) + const. \end{aligned} \tag{17}$$

We see that $q(\theta)$ factorises over $i$ and $q(\mathbf{z})$ factorises over $n, i$. Therefore, we get the variational distribution for $\mathbf{z}$ and $\theta$:

$$\theta_i \sim \mathbf{Dir}(\alpha_i + \sum_{n=1}^{N_s} \mathbf{E}\{z_{ni}\})$$

$$z_{ni} \sim \mathbf{Cat}\Big(\frac{\exp\big(\mathbf{E}\{\ln \theta_i\} + \sum_{j=1}^{D} w_{nj} \ln \beta_{ij}\big)}{\sum_{i=1}^{K} \exp\big(\mathbf{E}\{\ln \theta_i\} + \sum_{j=1}^{D} w_{nj} \ln \beta_{ij}\big)}\Big)$$

We then has the $(m+1)^{th}$ E-step as follows:

$$\begin{aligned}
\mathbf{E}^{m+1}\{\ln\theta_i\} &= \Psi(\alpha_i + \sum_{n=1}^{N_s}\mathbf{E}^m\{z_{ni}\}) - \Psi(\sum_{i=1}^{K}\big(\alpha_i + \sum_{n=1}^{N_s}\mathbf{E}^m\{z_{ni}\}\big)), \\
\mathbf{E}^{m+1}\{z_{ni}\} &= \frac{\exp\big(\mathbf{E}^{m+1}\{\ln\theta_i\} + \sum_{j=1}^{D} w_{nj}\ln\beta_{ij}^m\big)}{\sum_{i=1}^{K}\exp\big(\mathbf{E}^{m+1}\{\ln\theta_i\} + \sum_{j=1}^{D} w_{nj}\ln\beta_{ij}^m\big)},
\end{aligned} \tag{18}$$

where $\mathbf{E}^m$ and $\beta^m$ refers to the estimation of previous step ($m^{th}$ step); $\Psi$ is the digamma function.

To perform the M-step, we maximize the lower bound $\mathcal{L}$ in equation 2 for the entire population.

$$\begin{aligned}
\mathcal{L}(\mathbf{z},\theta,\beta,\alpha) = &\sum_{s=1}^{M}\Big(\ln\Gamma(\sum_{i=1}^{K}\alpha_i) - \sum_{i}^{K}\ln\Gamma(\alpha_i) + \sum_{i=1}^{K}(\alpha_i - 1)\mathbf{E}\{\ln\theta_{si}\} + \\
&\sum_{n=1}^{N_s}\sum_{i=1}^{K}(\mathbf{E}\{z_{sni}\}\mathbf{E}\{\ln\theta_{si}\}) + \\
&\sum_{n=1}^{N_s}\sum_{i=1}^{K}\mathbf{E}\{z_{sni}\}\sum_{j=1}^{D} w_{snj}\ln\beta_{ij}\Big) - \\
&\sum_{s=1}^{M}\Big(\ln\Gamma(\sum_{i=1}^{K}\big(\alpha_i + \sum_{n=1}^{N_s}\mathbf{E}\{z_{ni}\}\big)) - \sum_{i=1}^{K}\ln\Gamma(\alpha_i + \sum_{n=1}^{N_s}\mathbf{E}\{z_{ni}\}) + \\
&\sum_{i=1}^{K}(\alpha_i + \sum_{n=1}^{N_s}\mathbf{E}\{z_{ni}\} - 1)\mathbf{E}\{\ln\theta_{si}\} + \\
&\sum_{n=1}^{N_s}\sum_{i=1}^{K}\mathbf{E}\{z_{ni}\}\ln\mathbf{E}\{z_{ni}\}\Big)
\end{aligned} \tag{19}$$

For $\beta$, we take terms in $\mathcal{L}$ and add Lagrange multipliers:

$$\mathcal{L}_{[\beta]} = \sum_{i=1}^{K}\sum_{j=1}^{D}\ln\beta_{ij}\sum_{s=1}^{M}\sum_{n=1}^{N_s}\mathbf{E}\{z_{sni}\}w_{snj} + \sum_{i=1}^{K}\lambda_i(\sum_{j=1}^{D}\beta_{ij} - 1).$$

Set the derivative of $\mathcal{L}_{[\beta]}$ with respect $\beta$ to zero, we could get the $(n+1)^{th}$ update for *beta*:

$$\beta_{ij}^{n+1} = \frac{\sum_{s=1}^{M}\sum_{n=1}^{N_s}\mathbf{E}^{n+1}\{z_{sni}\}w_{snj}}{\sum_{j=1}^{D}\sum_{s=1}^{M}\sum_{n=1}^{N_s}\mathbf{E}^{n+1}\{z_{sni}\}w_{snj}}$$

The terms in lower bound that contains $\alpha$ are:

$$\mathcal{L}_{[\alpha_i]} = \sum_{s=1}^{M}\Big(\ln\Gamma(\sum_{i=1}^{K}\alpha_i) - \ln\Gamma(\alpha_i) + \sum_{i=1}^{K}(\alpha_i - 1)\mathbf{E}^{n+1}\{\ln\theta_{si}\}\Big).$$

Take the derivatives with respect to $\alpha$:

$$\frac{\partial \mathcal{L}_{[\alpha]}}{\partial \alpha_i} = M \cdot \Big( \Psi(\sum_{i=1}^{K} \alpha_i) - \Psi(\alpha_i) \Big) + \sum_{s=1}^{M} \mathbf{E}^{n+1}\{\ln \theta_{si}\}.$$

And the Hessian:

$$\nabla^2_{\alpha} \mathcal{L}_{[\alpha]} = M \cdot \mathrm{diag}(-\Psi^1(\alpha_i)\Big) + M \cdot \Psi^1(\sum_{i=1}^{K} \alpha_i),$$

where $\Psi^1$ is the Trigamma function. We use the Newton-Raphson method the find the maximal of $\alpha$ as described in [4]. In practice, we used $\alpha = 1$ to put an uninformative prior robust optimization.

#### 4.2 Patient with a few diseases versus documents with many words

In section 3.1, we briefly explained why we chose to use collapsed variational inference over a simpler mean-filed variational inference method. We will focus on the difference between equation 17 and equation 5. For the mean-field variational distribution:

$$\ln q^*(\mathbf{z}) = \sum_{n=1}^{N_s} \sum_{i=1}^{K} z_{ni} \Big( \mathbf{E}\{\ln \theta_i\} + \sum_{j=1}^{D} w_{nj} \ln \beta_{ij} \Big) + const,$$

which factorised over each of the $N$ diagnosis. Therefore, the inferred distribution for each $z_n$ is conditional i.i.d.

$$q(z_{n'} | \mathbf{z}_{\neg \mathbf{n'}}, \mathbf{w}, \alpha, \beta, \theta) = q(z_{n'} | \mathbf{w}, \alpha, \beta, \theta),$$

Here $\neg n'$ refer to indices of all diagnoses excluding $n'$. However, for collapsed VB, conditional distribution depends on other diagnoses of the same patient:

$$q(z_{n'} | \mathbf{z}_{\neg \mathbf{n'}}, \mathbf{w}, \alpha, \beta) \propto \prod_{i}^{K} (\alpha_i + \sum_{n \in \neg n'} z_{ni})^{z_{n'i}} \prod_{i=1}^{K} \prod_{j=1}^{D} \beta_{ij}^{z_{n'i} w_{n'j}}$$

The impact of the the dependency on the accuracy of posterior approximation depends on the data structure. Most of topic modelling modes were designed for text modelling, where each document have a large word number $N_s$. In this case, $(\alpha_i + \sum_{n \in \neg n'} z_{ni})$ will be approximately the same across $n'$:

$$\lim_{N_s \to \infty} p(z_{n'} | \mathbf{z}_{\neg \mathbf{n'}}, \mathbf{w}, \alpha, \beta) \propto \prod_{i}^{K} \Big[ (\alpha_i + N_s \theta_i) \prod_{j=1}^{D} \beta_{ij}^{w_{n'j}} \Big]^{z_{n'i}},$$

where $\theta_i$ is the topic value for the $s^{th}$ document. We see $\mathbf{z}_{\neg\mathbf{n}'}$ no longer exist and $q^*(\mathbf{z})$ in equation 17 could approximate $p(\mathbf{z}|\mathbf{w}, \alpha, \beta)$ accurately. However, each patient on average have 6.1 distinct diagnoses in UK Biobank HES, making $N_s$ small for the mean-field approximation. Note, we do not need to assume independence of $z_n$, it is a consequence of assume independence between $q(\theta)$ and $q(\mathbf{z})$, which is called induced factorisation in some cases. (section 10.2.5 in [2]) In this cases collapsed VB models the dependency between $\mathbf{z}_n$ and $\mathbf{z}_{\neg\mathbf{n}'}$ and have better accuracy at approximating posterior distribution.
